# Pluripotent stem cell-based drug discovery uncovers sildenafil as a treatment for mitochondrial disease

**DOI:** 10.1101/2025.05.15.25325571

**Authors:** Annika Zink, Dao-Fu Dai, Annika Wittich, Marie-Thérèse Henke, Giulia Pedrotti, Sonja Heiduschka, Guillem S. Aguilar, Tancredi Massimo Pentimalli, Christian Brueser, Sofia Notopoulou, Aleksandra Zhaivoron, Abdul Rahim Umar, Laura Petersilie, Caleb Jerred, Jesper Bergmans, Fabian Schumacher, Jan Keller-Findeisen, Agnieszka Rybak-Wolf, Daniel Stach, Jeanette Reinshagen, Andrea Zaliani, Undine Haferkamp, Liliya Euro, Alessia Di Donfrancesco, Chiara Santanatoglia, Enrica Cappellozza, Marta Suarez Cubero, Mario Pavez-Giani, Oleh Bakumenko, David Meierhofer, Alan Foley, Susanne Morales-Gonzalez, Isabella Tolle, Diran Herebian, Daniele Bonesso, Ildiko Szabo, Giulia Cecchetto, Sakurako Wong, Monica Moresco, Alessandra Maresca, Ilaria Decimo, Merel J.W. Adjobo-Hermans, Burkhard Kleuser, Lukas Cyganek, Chris Mühlhausen, Lars Schlotawa, Valeria Tiranti, Andrea Rossi, Ertan Mayatepek, Chiara La Morgia, Valerio Carelli, Thomas Klopstock, Felix Distelmaier, Ghanim Ullah, Stefan Jakobs, Nikolaus Rajewsky, Christine R. Rose, Spyros Petrakis, Frank Edenhofer, Werner Koopmann, Dario Brunetti, Pawel Lisowski, Anu Suomalainen, Antonio del Sol, Emanuela Bottani, Ole Pless, Markus Schuelke, Alessandro Prigione

**Affiliations:** Department of General Pediatrics, Neonatology and Pediatric Cardiology, Medical Faculty, University Hospital Düsseldorf, Heinrich-Heine-University, Düsseldorf, Germany; Department of Pathology, Johns Hopkins University School of Medicine, Baltimore, USA; Fraunhofer Institute for Translational Medicine and Pharmacology ITMP, Discovery Research, ScreeningPort, Hamburg, Germany; Charité Universitätsmedizin Berlin, corporate member of Freie Universität Berlin and Humboldt-Universität zu Berlin, Department of Neuropediatrics, Berlin, Germany; Charité Universitätsmedizin Berlin, corporate member of Freie Universität Berlin and Humboldt-Universität zu Berlin, NeuroCure Clinical Research Center, Berlin, Germany; Department of Diagnostics and Public Health, University of Verona, Italy; Faculty of Mathematics and Natural Sciences, Heinrich Heine University, Düsseldorf, Germany; Computational Biology Group, Luxembourg Centre for Systems Biomedicine, University of Luxembourg, Esch-sur-Alzette, Luxembourg; Berlin Institute for Medical Systems Biology (BIMSB), Max Delbrück Center for Molecular Medicine in the Helmholtz Association (MDC), Berlin, Germany; Charité – Universitätsmedizin Berlin, Germany; Fraunhofer Institute for Translational Medicine and Pharmacology ITMP, Translational Neuroinflammation and Automated Microscopy, Göttingen, Germany; Institute of Applied Biosciences (INAB), Centre For Research and Technology Hellas (CERTH), Thessaloniki, Greece; University of Helsinki, Stem Cells and Metabolism Program, Faculty of Medicine, Helsinki, Finland; Department of Physics, University of South Florida, Tampa, USA; Institute of Neurobiology, Heinrich Heine University, Düsseldorf, Germany; Radboud Center for Mitochondrial Medicine, Radboud University Medical Center, Nijmegen, The Netherlands; Department of Pediatrics, Amalia Children’s Hospital, Radboud University Medical Center, Nijmegen, The Netherlands; Institute of Pharmacy, Freie Universität Berlin, Berlin, Germany; Fondazione IRCCS Istituto Neurologico Carlo Besta, Milan, Italy; Department of Molecular Biology & CMBI, Genomics, Stem Cell & Regenerative Medicine Group, University of Innsbruck, Innsbruck, Austria; Stem Cell Unit, Clinic for Cardiology and Pneumology, University Medical Center Göttingen, Göttingen, Germany; Experimental Cardiology Institute, Medical Clinic I/Cardiology and Angiology, Justus Liebig University of Giessen, Giessen; Quantitative RNA Biology, Max Planck Institute for Molecular Genetics, Berlin, Germany; Department of Biology, University of Padova; IRCCS Istituto delle Scienze Neurologiche di Bologna, Programma di Neurogenetica, Bologna, Italy; Department of Medical BioSciences, Radboud Center for Mitochondrial Medicine, Radboud University Medical Center, Nijmegen, The Netherlands; Department of Pediatrics and Adolescent Medicine, University Medical Center, Georg-August-University, Göttingen, Germany; Genome Engineering and Model Development lab (GEMD), IUF-Leibniz Research Institute for Environmental Medicine, Düsseldorf, Germany; Department of Biomedical and Neuromotor Sciences (DIBINEM), University of Bologna, Bologna, Italy; Friedrich-Baur-Institute, Department of Neurology, LMU University Hospital, Ludwig-Maximilians-Universität München, 80336 Munich, Germany; Munich Cluster for Systems Neurology (SyNergy), 81377 Munich, Germany; German Center for Neurodegenerative Diseases (DZNE), Munich, Germany; Department of NanoBiophotonics, Max Planck Institute for Multidisciplinary Sciences, Göttingen, Germany; Department of Neurology, University Medical Center Göttingen, Göttingen, Germany; Human and Animal Physiology, Wageningen University, Wageningen, The Netherlands; Department of Clinical Sciences and Community Health, Dipartimento di Eccellenza 2023-2027, University of Milan, Milan, Italy; Neuropsychiatry and Laboratory of Molecular Psychiatry, Department of Psychiatry and Neurosciences, Charité Universitätsmedizin Berlin, Germany; Department of Molecular Biology, Institute of Genetics and Animal Biotechnology, Polish Academy of Sciences, Jastrzebiec n/Warsaw, Poland; HiLife, University of Helsinki, Helsinki, Finland; HUS Diagnostics, Helsinki University Hospital, Helsinki, Finland; IKERBASQUE, Basque Foundation for Science, Bilbao, Spain; CIC bioGUNE-BRTA (Basque Research and Technology Alliance), Bizkaia Technology Park, Derio, Spain; German Center for Child and Adolescent Health (DZKJ), Section CNS Development and neurological Disease, partner site Berlin, Germany

## Abstract

Mitochondrial disease encompasses inherited disorders affecting mitochondrial function. A severe and untreatable form of mitochondrial disease is Leigh syndrome (LS) causing psychomotor regression and metabolic crises. To accelerate drug discovery for LS, we screened a library of 5,632 repurposable compounds in induced pluripotent stem cell (iPSC)-derived neural progenitor cells (NPCs) from LS patients. We identified phosphodiesterase 5 inhibitors (PDE5i) as leads, and prioritized sildenafil due to its safety profile. Sildenafil restored pathways regulating nervous system development, enhanced neurite outgrowth in LS neurons, and mitigated abnormal calcium responses in LS brain organoids under metabolic stress. In a mouse model of LS, sildenafil extended the lifespan and ameliorated metabolic and encephalopathy phenotypes. Chronic off-label compassionate treatment with sildenafil in six LS patients showed improvements in motor function and resistance to metabolic crises. These findings highlight the potential of iPSC-driven drug discovery and position sildenafil as a promising candidate for mitochondrial diseases.

## Introduction

Mitochondrial disease refers to a spectrum of rare genetic conditions caused by dysfunction in the cell’s energy-producing organelles^1^.

A severe form of mitochondrial disease is Leigh syndrome (LS, OMIM #256000), which is characterized by basal ganglia and brainstem necrosis leading to neurodevelopmental regression and muscle weakness. Early death in these patients is typically due to acute deterioration following metabolic crises triggered by events that place excessive demands on the body’s energy production, such as infection or surgery^2,3^. LS can result from pathogenic gene variants in more than 100 genes in the nuclear DNA or mitochondrial DNA (mtDNA) that are involved in oxidative phosphorylation (OXPHOS)^4^. Commonly affected genes include the complex V gene *MT-ATP6* (mitochondrially encoded ATP synthase subunit 6)^5^, the complex IV assembly factor gene *SURF1* (SURFEIT1)^6^, and complex I genes such as *NDUFS4* (NADH dehydrogenase [ubiquinone] iron-sulfur protein 4)^7^.

Currently, there are no treatments for LS^8^. One of the significant challenges in identifying disease-modifying therapies for LS is the limited availability of appropriate model systems^9^. The most widely used model of LS is the homozygous *Ndufs4* knockout (KO) mouse, which develops progressive encephalopathy, growth retardation, motor regression, cardiomyopathy, and premature death at approximately 50 days of age^10,11^. This mouse is however the only rodent model that recapitulates the brainstem pathology of LS patients. For example, *Surf1* KO mice failed to replicate LS phenotypes and instead showed increased longevity^12^, implicating different capacity to compensate for specific pathways between mice and humans^13^. In addition, the limitations of CRISPR/Cas9-based genome editing technologies in targeting mtDNA have hindered the establishment of models for mtDNA defects such as *MT-ATP6* mutations^14^. Potential treatment strategies have been proposed using the *Ndufs4* KO mouse model, including antioxidants^15–17^, rapamycin^18^, interferon-gamma-targeting therapies^19^, or hypoxia^20,21^. However, only few of these strategies have been tested in clinical application in LS patients, and no successful results have been obtained.

One approach for establishing models of LS suitable for drug discovery studies is the use of patient-derived induced pluripotent stem cells (iPSCs). iPSCs allow the generation of disease-relevant two-dimensional (2D) or three-dimensional (3D) cellular models containing patient-specific nuclear or mitochondrial defects^22^. In LS, such models have been instrumental in uncovering pathological mechanisms. Identified phenotypes include neuronal outgrowth defects^23^, altered calcium homeostasis^24,25^, and glutamate toxicity in 2D neuronal cultures^26^, as well as impaired cortical development and neuromorphogenesis in 3D brain organoids^23,27,28^. Despite these advances, large-scale high-throughput drug screens in iPSC models of mitochondrial disease have yet to be performed.

We have previously demonstrated that iPSC-derived neural progenitor cells (NPCs) are an effective drug discover platform for mitochondrial diseases^25^. We found that LS NPCs with *MT-ATP6* defects exhibit abnormal hyperpolarization of mitochondrial membrane potential (MMP), a feature that can be exploited for high-throughput screens using high-content microscopy^29^. Here, we leveraged on this MMP phenotype to screen a library of 5,632 repurposable drug candidates in NPCs from LS patients harboring *MT-ATP6* mutations. We identified phosphodiesterase type 5 inhibitors (PDE5i) as lead compounds capable of ameliorating the MMP defect. Among these, we prioritized sildenafil given its known safety profile in infants and adults^30^. Sildenafil rescued the neurodevelopmental disease signature, promoted neuronal outgrowth, and normalized calcium homeostasis in 2D and 3D human neuronal models of LS carrying different pathogenic gene variants. *In vivo*, sildenafil extended the lifespan of *Ndufs4* KO mice and improved their metabolic expenditure. Mechanistically, the action of sildenafil appeared to be mediated through modulation of cGMP-dependent protein kinase 1 (PRKG1). Lastly, individualized off-label compassionate treatments with sildenafil in six patients with LS carrying different *MT-ATP6* mutations resulted in significant clinical improvement of muscle strength and endurance and resistance to metabolic crises. The data collectively suggest a potential use of sildenafil as a repurposable drug for individuals with the mitochondrial disease Leigh syndrome.

## Results

### Small molecule repurposing screen in LS NPCs identifies PDE5 inhibitors

We previously detected abnormally hyperpolarized MMP in LS NPCs carrying the m.9185T>C mutation in the *MT-ATP6*, which encodes the “a” subunit of complex V involved in the release of the proton gradient established across the inner mitochondrial membrane^25^. As MMP levels can be used as a phenotypic readout for compound screening^29^, we set out to determine whether such a phenotype could be robustly observed across LS NPC lines carrying different *MT-ATP6* mutations. We generated NPCs from seven LS patient-derived iPSCs carrying different *MT-ATP6* mutations (three m.9185T>C, one m.8993T>C, two m.8993T>G, and one m.9176T>G) **(Figure S1a)**^25,31–34^. For controls, we used NPCs from eight iPSC lines derived from healthy individuals, including two patient mothers who carried *MT-ATP6* mutations at low level of heteroplasmy (less than 2 % in iPSCs and NPCs) **(Figure S1e)**. LS NPCs showed disrupted ATP6 protein expression **(Figure S1b)**, but expressed NPC markers similarly to control NPCs, demonstrating physiological neural commitment **(Figure S1c-d)**. They also retained the mtDNA mutations at the same level of heteroplasmy as the parental iPSCs **(Figure S1e)**. Importantly, in agreement with our previous observations^25^, all LS NPCs displayed mitochondrial membrane hyperpolarization when compared to control NPCs **(Figure 1a)**.

**Figure 1.**
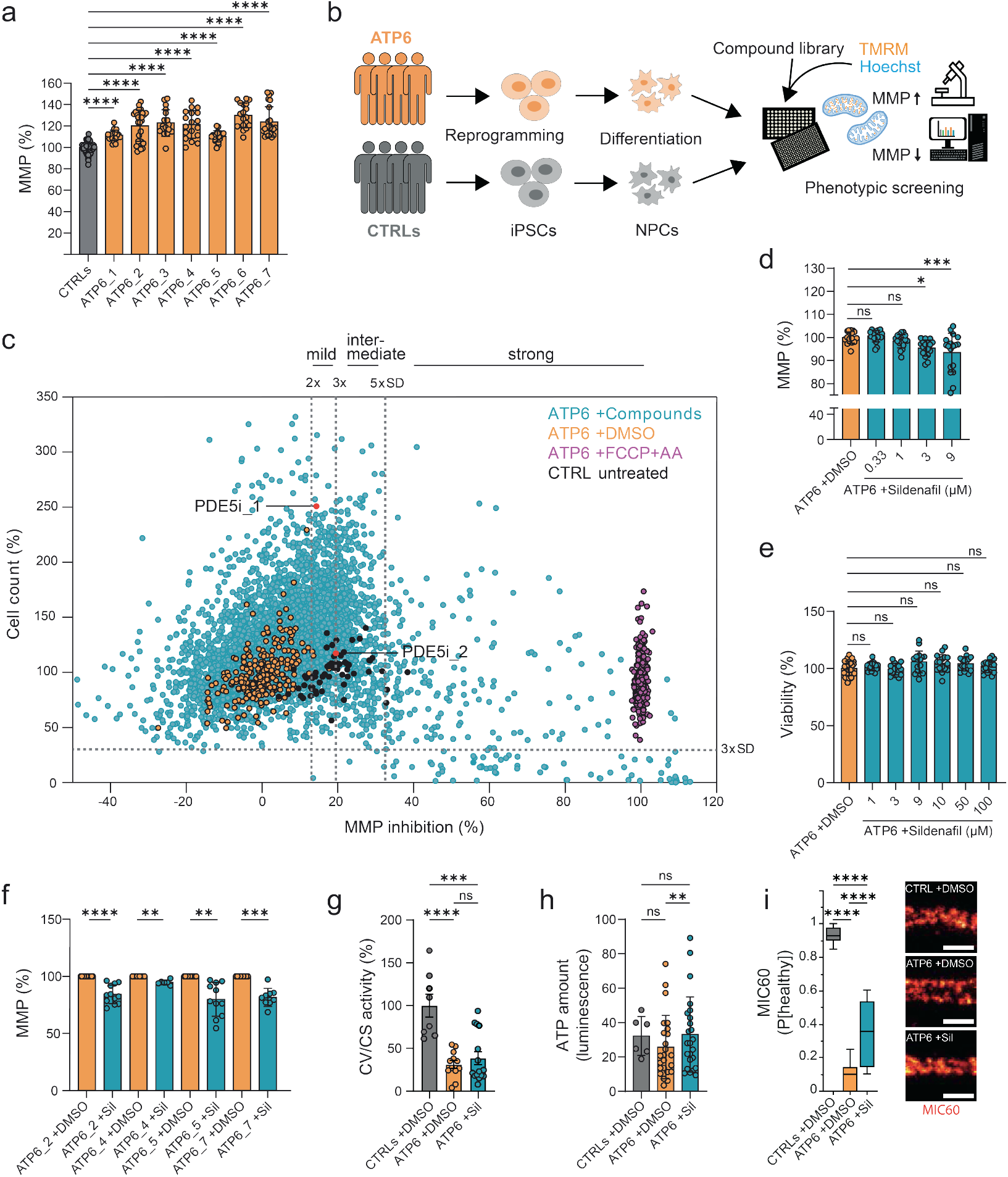
Compound screen in LS NPCs leads to the identification of the PDE5i sildenafil. **a**. High-content analysis (HCA)-based quantification of mitochondrial membrane potential (MMP) in neural progenitor cells (NPCs) from LS patients (ATP6_1, ATP6_2, ATP6_3, ATP6_4, ATP6_5, ATP6_6, ATP6_7) and healthy controls (CTRL_1). Dots represent mean values per well out of n=3-5 independent experiments. ****p<0.0001; ordinary one-way ANOVA with Dunnett’s multiple comparison. **b**. Schematic of drug screen approach. **c**. MMP screening of 5,632 repurposable compounds. Scatter dot plot illustrating the percentage of MMP depolarization (TMRM signal) against the percentage of cell count (Hoechst signal) after treatment with 5 *μ*M for 16 h of each compound in LS NPCs normalized to controls. Dots represent individual wells of LS NPCs (ATP6_2) treated with either compounds (blue), DMSO (orange), or 5 *μ*M FCCP+AA (purple). Untreated control NPCs (CTRL_1) are shown in black. PDE5 inhibitor (PDE5i) 1 (T-0156) and PDE5i 2 (vardenafil hydrochloride) are highlighted in red. **d**. Dose-dependent effect of PDE5i sildenafil on the MMP of LS NPCs (ATP6_2) treated for 16 h. Dots represent mean values per well out of n=4 independent experiments normalized to DMSO only; *p<0.05, ***p<0.001; ns (not significant); ordinary one-way ANOVA with Dunnett’s multiple comparison. **e**. Cell viability in LS NPCs (ATP6_2) treated for 16 h with increasing concentrations of sildenafil. Dots represent individual wells out of 16 replicates per sildenafil concentration and 31 replicates for DMSO. ns; ordinary one-way ANOVA with Dunnett’s multiple comparison. **f**. Effect of sildenafil on the MMP of LS NPCs. Dots represent individual cytofluorimetry measurements with TMRM using 10 *μ*M sildenafil for 16 h out of n=3 independent experiments. **p<0.01, ***p<0.001, ****p<0.0001; two-tailed paired t-test. **g**. Complex V (CV) activity normalized to citrate synthase (CS) activity. Dots represent a pool of 10-15 million cells in n=3 independent experiments of CTRL NPCs (CTRL_1, CTRL_3) and ATP6 NPCs (ATP6_2, ATP_4, ATP6_5, ATP6_7) using 10 *μ*M sildenafil for 16 h. ***p<0.001, ****p<0.0001, ns; unpaired two-tailed t-test. **h**. Luminescence-based ATP content quantification. Dots represent biological replicates out of n=3 independent experiments in control NPCs (CTRL_1, CTRL_3) and LS NPCs (ATP6_2, ATP6_4, ATP6_5, ATP6_7) grown in glucose-free galactose medium using 10 *μ*M sildenafil for 16 h. *p<0.05, **p<0.01, ns; paired t-test. **i**. Left, machine learning analysis of the MIC60 labeling pattern of stimulated emission depletion microscopy (STED) images in control NPCs (CTRL_1) and LS NPCs (ATP6_2) using 10 *μ*M sildenafil for 16 h. Quantification shows the probability of the pattern to be similar to that of untreated control NPCs (p[healthy]). Right, representative STED images. ****p<0.0001; Kruskal-Wallis with Dunn’s multiple comparison test.

We then applied a well-curated library of 5,632 repurposable molecules^35^ to assess their impact on the MMP of LS mutant NPCs. We used live-cell high-content analysis (HCA) based on the intensity of the MMP-dependent fluorescent dye tetramethylrhodamine methyl ester (TMRM)^29^ and Hoechst 33342 for counter staining **(Figure 1a, Figure S2a-b)**. All compounds were resuspended in dimethylsulfide (DMSO) at a maximal concentration of 0.05 % DMSO, which did not affect the viability of LS NPCs **(Figure S2c)**. In the screening plates, we included one control NPC line for baseline MMP value, after having confirmed that different control NPC lines displayed comparable MMP levels **(Figure S2d)**.

To establish the Z’ factor^36^ of our assay, we exploited the MMP depolarizing effect of the mitochondrial uncoupler carbonyl cyanide-p-trifluoromethoxyphenylhydrazone (FCCP) in combination with the mitochondrial complex III inhibitor antimycin A (AA)^29^ FCCP+AA applied at a concentration of 5 *μ*M each proved effective in depolarizing the MMP without loss of cell viability **(Figure S2e-f)**, resulting in an excellent Z’ factor of 0.68 0.85 across all screening plates **(Figure S2g)**. Next, we performed MMP assays with a representative compound plate at different concentrations. We identified 5 *μ*M as the optimal screening concentration, achieving a hit rate of ~5 % with overall low toxicity **(Figure S2h)**. Screening of duplicates confirmed high data reproducibility across plates on different days **(Figure S2i)**.

We classified the 5,632 compounds based on their depolarizing effect on the hyperpolarized MMP of LS NPCs (MMP inhibition) relative to the standard deviation (SD) of DMSO-treated cells (SD=6.51 %). We identified 767 mild uncoupler molecules that depolarized the MMP by more than two times the SD (13.0 19.5 % inhibition), 520 intermediate uncoupler molecules that depolarized the MMP by more than three times the SD (19.5 32.5 % inhibition), and 187 strong uncoupler molecules that depolarized the MMP by more than five times the SD **(Figure 1c)**. We classified compounds as non-toxic using the cell count of DMSO-treated cells (cut-off: 29.86 %). We focused on mild uncouplers, as they were sufficient to depolarize the MMP of LS NPCs to values similar of those of control NPCs **(Figure 1c)**. Among those, we identified two phosphodiesterase type 5 inhibitors (PDE5i): T-0156 and vardenafil hydrochloride. This was of special interest to us, because our previous small-scale proof-of-concept screen of 140 repurposable drugs in LS NPCs had highlighted the beneficial effect of the PDE5i avanafil^25^. In follow up experiments, we confirmed that both T-0156 and vardenafil hydrochloride led to dose-dependent mild depolarization of MMP in LS NPCs without loss of viability **(Figure S3a-d)**.

Given these current and previous results for various PDE5i molecules, we searched the literature for applications of PDE5i in pediatric diseases. Sildenafil emerged as a strong candidate, given its safety profile in children treated for pulmonary hypertension or lymphatic malformations^37–40^. As seen with the other PDE5i **(Figure S3a-d)**, sildenafil led to a dose-dependent depolarization of MMP in LS NPCs without inducing toxicity **(Figure 1d-e, Figure S3e-f)**. Sildenafil treatment normalized the hyperpolarized MMP of LS NPCs with an IC50 of approximately 3 *μ*M after 16 hours of exposure **(Figure S3g)** and did not affect cell proliferation **(Figure S3h)**. The sildenafil-induced MMP improvement occurred similarly in several LS NPC lines carrying distinct *MT-ATP6* mutations **(Figure 1f)**. Sildenafil-treated control NPCs showed no changes in MMP quantified by either HCA or cytofluorimetry **(Figure S3i)**.

We next assessed the bioenergetic effects of sildenafil in LS NPCs. Sildenafil treatment in LS NPCs made complex V activity more similar to that of control NPCs **(Figure 1g)**. The effects of sildenafil became more evident when NPCs were grown in glucose-free galactose medium, which resulted in an increased intracellular ATP concentration in LS NPCs **(Figure 1h)**. Since *MT-ATP6* defects can disrupt mitochondrial cristae morphology^41^, we analyzed the distribution of mitochondrial crista junctions using the cristae junction regulator MIC60 as a proxy with stimulated emission depletion (STED) microscopy^42^. We inspected the MIC60 labeling pattern of 4,000 STED images with a machine learning approach by training a neural network classifier to distinguish healthy control NPCs (P[healthy]-score = 1) from LS NPCs (P[healthy]-score = 0). Using images not used for training, we detected alterations of the localization of MIC60 fluorescence signals in LS NPCs that were partially reversed by sildenafil treatment **(Figure 1i, Figure S3j-k)**. Manual quantification of approximately 300 STED images yielded similar results **(Figure S3l)** and highlighted a peripheral distribution pattern of MIC60 in LS NPCs that was normalized by sildenafil **(Figure S3m-n)**. Collectively, these data suggest sildenafil as a potential treatment strategy for LS capable of improving mitochondrial phenotypes.

### The PDE5i sildenafil ameliorates the LS disease signature by modulating neuronal development pathways

To gain insight into the mechanisms of action of sildenafil in LS, we selected four LS NPC lines (one for each of the *MT-ATP6* mutations: m.9185T>C, m.8993T>C, m.8993T>G, and m.9176T>G) and four healthy control NPC lines to perform multi-omics analysis. We used 10 *μ*M sildenafil treatment for 16 hours, which was sufficient to elicit changes in mitochondrial function **(Figure 1f-i)**. We compared samples treated with sildenafil dissolved in 0.1 % DMSO to samples treated with only 0.1 % DMSO. The omics design would allow us to identify: i) the disease signature, by comparing DMSO-treated LS NPCs to DMSO-treated control NPCs, ii) the sildenafil signature, by comparing sildenafil-treated LS NPCs to DMSO-treated LS NPCs **(Figure 2a)**.

**Figure 2.**
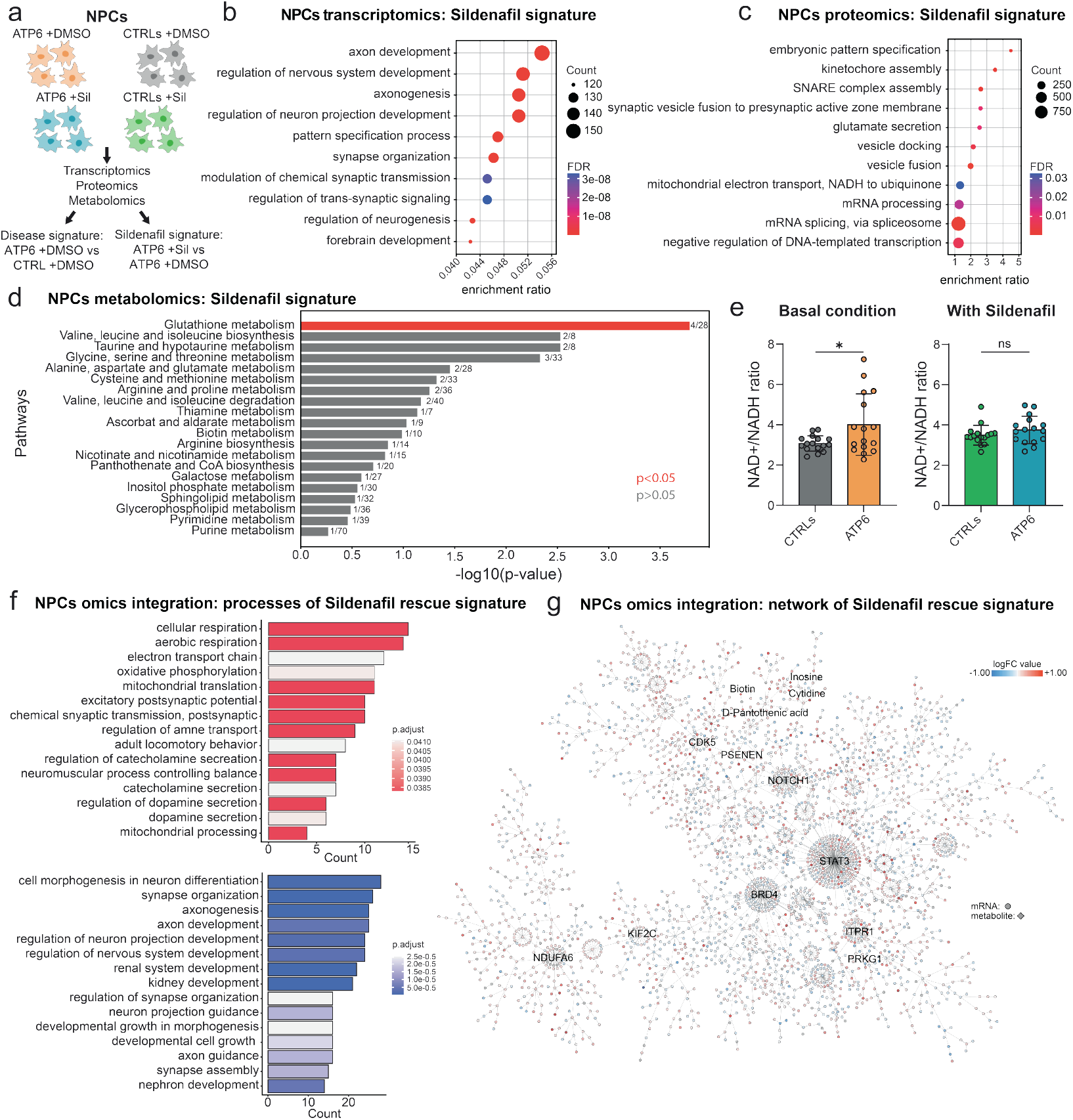
Neurodevelopmental impact of sildenafil unveiled by multi-omics. **a**. Schematic of omics setup in control NPCs (CTRL_1, CTRL_2, CTRL_3, CTRL_4) and LS NPCs (ATP6_2, ATP_4, ATP6_5, ATP6_7) to dissect the disease signature and the sildenafil signature (10 *μ*M for 16 h) compared to DMSO only (n=3 independent biological replicates per sample). **b**. Dot plot highlighting top ten enriched Gene Ontology (GO) biological processes in LS NPCs upon sildenafil treatment based on transcriptomics. **c**. Dot plot highlighting top ten enriched GO biological processes in LS NPCs upon sildenafil treatment based on proteomics. **d**. Improved metabolic pathways in LS NPCs treated with sildenafil based on metabolomics. One-way ANOVA with false discovery rate (FDR) adjustment for multiple comparison (p value 0.05). **e**. Quantification of NAD+/NADH ratio in NPCs under basal condition and upon sildenafil treatment. *p<0.05, ns; unpaired two-tailed t-test. **f**. Enrichment analysis for 724 reversed molecules by sildenafil based on multi-omics integration. GO biological processes for reversed molecules with logarithmic fold change (logFC)>0 after sildenafil treatment. **g**. 3D multi-omics network of the sildenafil signature in LS NPCs. Size of dots defines their centrality in the network; color of dots indicates their logFC value.

Global transcriptomics revealed a disease signature affecting biological processes related to nervous system development and axon development **(Figure S4a)**, and cellular components related to mitochondrial inner membrane and neuronal cell body **(Figure S4b)**. The sildenafil signature in LS NPCs affected similar biological processes, including axon development and nervous system development **(Figure 2b)**, and cellular components, such as mitochondrial inner membrane and neuronal cell body **(Figure S4c)**. These data indicated that dysregulation of key genes and pathways resulting from the *MT-ATP6* mutations could be counteracted with sildenafil. For example, the neurodevelopment-associated gene developing homeobox 1 (*DBX1*)^43^ was downregulated in LS NPCs compared to control NPCs **(Figure S4d)** and upregulated in treated LS NPCs compared to untreated **(Figure S4e, Table S1)**. Changes induced by sildenafil appeared specific for LS NPCs, as no significant transcriptional effects were observed in sildenafil-treated control NPCs **(Figure S4k)**.

Proteomic analysis revealed a disease signature associated with dysregulation of biological processes of mitochondrial gene expression and cell projection **(Figure S4f)** and cellular components of the inner mitochondrial membrane and of axons **(Figure S4g)**. The sildenafil signature was associated with pathways involved in development, synaptic activity, and electron transport chain (ETC) including NADH-dependent activities **(Figure 2c, Figure S4h)**. Thus, similar to the transcriptomic findings, proteomic data indicated that sildenafil restored pathways impaired by *MT-ATP6* mutations. For example, the protein BAG4 that protects against ATP depletion-induced cell death^44^ was downregulated in LS NPCs compared to control NPCs **(Table S2)** and upregulated in LS NPCs treated with sildenafil **(Figure S4j)**.

Metabolomics identified key metabolites altered by the disease, including creatine and cysteine **(Figure S5a)**, which were normalized by sildenafil treatment **(Figure S5b, Table S3)**. The sildenafil signature mainly affected the glutathione metabolism pathway **(Figure 2d, Figure S5c-d)**. Consistent with the metabolomics and proteomics signature of sildenafil modulating NADH-dependent pathways **(Figure S4h)**, the NAD+/NADH ratio was significantly altered in LS NPCs compared to control NPCS and normalized upon sildenafil treatment **(Figure 2e)**.

We next performed multi-omics integration as previously described^45^. The integrated disease signature confirmed the enrichment of dysregulated mRNAs, proteins, and metabolites that are known to have an impact on development, WNT and Notch signaling, and axon guidance **(Figure S5e)**. The integrated sildenafil signature affected similar pathways, including development, Notch signaling, and axon guidance **(Figure S5f)**. The integrated disease signature dataset and the integrated treatment signature dataset shared 796 molecules, with 724 exhibiting opposite regulation upon sildenafil administration. Hence, sildenafil reversed 33.5 % of dysregulated genes in the cellular model (724 reversed out of 2160 dysregulated molecules). Biological processes reversed by sildenafil included two groups, one related to mitochondrial respiration and one related to morphogenesis and axon development **(Figure 2f)**. The multi-omics map of the sildenafil rescue signature in LS NPCs (including only transcriptomics and metabolomics, as proteomics did not show sufficient significant changes) highlighted a network including metabolites like biotin and pantothenic acid and genes associated with ETC function (e.g. *NDUFA6*), signaling and development (e.g. *NOTCH1, STAT3*), and the cGMP-dependent protein kinase 1 (*PRKG1*), a known master regulator and downstream target of PDE5^46^ **(Figure 2g)**.

### LS brain organoids show neurodevelopmental defects and highlight the impact of sildenafil on progenitor populations

As both the LS disease signature and sildenafil signature in NPCs included genes related to nervous system development **(Figure 2, Figure S4-5)**, we investigated this process using cortical brain organoids. In agreement with previous studies in LS brain organoids^23,27,28^, *MT-ATP6* mutations impaired neurogenic zone formation **(Figure 3a)** and altered the ratio of early neurons to neural progenitors **(Figure 3b)**. Two different protocols for generating cortical brain organoids showed defective growth rates in the presence of *MT-ATP6* mutations **(Figure S6a)**. One protocol^47^ showed size defects in LS brain organoids after 50 days in culture **(Figure S6b)**, whereas another protocol that required additional growth factors and the use of AggreWells plates^48^ allowed an initial homogenous organoid shape, resulting in earlier growth defects in LS brain organoids that became less pronounced over time **(Figure S6b)**. These results collectively suggest that *MT-ATP6* mutations might affect neural progenitor development.

**Figure 3.**
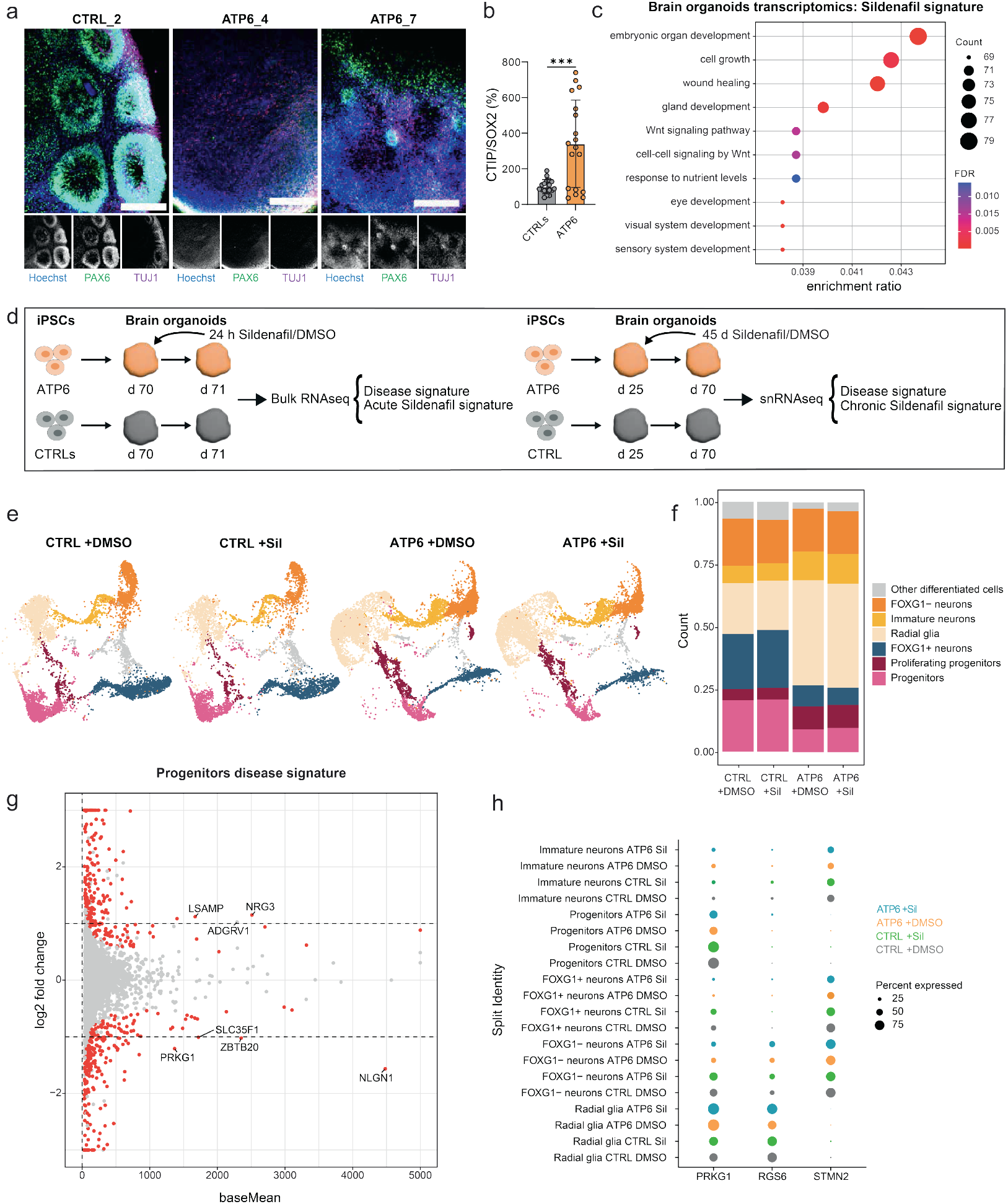
Sildenafil treatment in LS brain organoids affects progenitor populations. **a**. Representative images of cortical brain organoids from controls (CTRL_2) and LS (ATP6_4, ATP6_7) at day 35 stained with neural progenitor marker PAX6 and neuronal marker TUJ1. Scale bar: 200 *μ*m. **b**. Differentiation ratio based on gene expression of *CTIP2* over *SOX2* in day 70 brain organoids from controls (CTRL_1 and CTRL_2) and LS (ATP6_4, ATP6_7). 2-3 brain organoids per line per experiment, n=3 independent experiments. ***p<0.001, ns; unpaired two-tailed t-test. **c**. Dot plot highlighting top ten enriched GO biological processes by sildenafil (10 *μ*M for 24 h) in day 70 LS brain organoids (ATP6_7) based on transcriptomics. 5 brain organoids per line per experiment, n=3 independent experiments. **d**. Schematic depicting the experimental setup to assess the acute sildenafil signature in day 70 brain organoids with bulk transcriptomics from controls (CTRL_1, CTRL_2, ATP6_4, ATP6_7) and LS (ATP6_4, ATP6_7) and the chronic sildenafil signature with single-nucleus RNA sequencing (snRNAseq) from controls (CTRL_1) and LS (ATP6_7). **e-f**. Uniform manifold approximation and projection (UMAP) plot and bar plot showing distribution of day 70 brain organoids from controls (CTRL_1) and LS (ATP6_7) treated with either sildenafil (10 *μ*M for 45 days) or only DMSO. 15 brain organoids per line per experiment, n=3 independent experiments. **g**. Volcano plot of pseudo-bulk analysis of snRNAseq dataset depicting the disease signature in the progenitor population. **h**. Pseudo-bulk analysis of snRNAseq dataset showing the expression pattern of *PRKG1, RGS6*, and *STMN2* across distinct populations of day 70 brain organoids.

We treated LS brain organoids with sildenafil for either 24 hours (acute paradigm) or 45 days (chronic paradigm) **(Figure 3d)**. Global transcriptomics showed that *MT-ATP6* mutations in brain organoids elicited responses similar to those seen in NPCs, including effects on biological processes related to synapses and neuronal projections **(Figure S6e)**, as well as effects on cellular components related to neuronal cell bodies and synapses **(Figure S6f)**. The acute sildenafil signature in LS brain organoids affected biological processes related to embryonic development and WNT signaling **(Figure 3c)** and corrected key gene defects. For example, the *WDR45B* gene whose mutations are associated with neurodevelopmental disorders^49^ was downregulated in LS brain organoids compared to control brain organoids **(Table S4)** and upregulated in sildenafil-treated LS brain organoids **(Figure S6d)**.

To gain insight into the cell populations most affected by *MT-ATP6* mutations and by chronic sildenafil treatment, we performed single-nucleus RNA sequencing (snRNAseq) **(Figure 3d)**. Unsupervised clustering highlighted 9 clusters within cortical brain organoids at day 70 **(Figure S7a-b, Table S5)**. Based on their gene expression patterns, we assigned clusters 0 and 3 to radial glia cells, clusters 4 and 8 to progenitors, cluster 6 to proliferating progenitors, cluster 5 to immature neurons, cluster 2 to FOXG1-positive neurons, cluster 1 to FOXG1-negative neurons, and cluster 7 to other cell types **(Figure S7a-b, Table S5)**. This annotation showed that *MT-ATP6* mutations impaired neuronal commitment, with alterations in the populations of radial glia cells, progenitors, and FOXG1-positive neurons **(Figure 3e-f)**. The disease signature highlighted the downregulation in the progenitor population of neuroligin 1 (*NLGN1*), a master regulator for synapse development^50^, and the PDE5 target *PRKG1* **(Figure 3g)**, and the downregulation in the neuronal population of neuronal outgrowth-associated genes such as *STMN2*^51^ **(Figure S7d, Table S6)**. Differential gene expression revealed that the effect of sildenafil was mostly evident in radial glia cells, progenitors, and FOXG1-positive neurons **(Figure S7c, Table S6)**. The sildenafil signature uncovered the upregulation of neuronal outgrowth-associated gene as *RGS6*^52^ across several populations **(Figure S7c-d, Table S6)**. While *STMN2* was mainly present in FOXG1-positive neurons and immature neurons, *RGS6* was found mainly in radial glia cells, and *PRKG1* in radial glia cells and progenitors **(Figure 3h, Figure S7d)**. We next inspected the glycolytic signature, which has been suggested as an indicator of brain organoid stress^53^. This signature was not altered in our samples, suggesting that neither *MT-ATP6* mutations nor sildenafil treatment posed additional stress on brain organoids **(Figure S7e)**. Taken together, LS mutations disrupted brain organoid development by impairing early neuronal organization of radial glia and progenitor populations, and sildenafil treatment specifically affected those populations.

### Sildenafil improves calcium homeostasis and neurite outgrowth in human LS models

We next examined the functional consequences of sildenafil treatment. A crucial aspect of the pathogenesis of LS is the sudden deterioration of patients following episodes of metabolic decompensation^3^. We therefore aimed to induce acute metabolic stress in LS brain organoids and determine the resulting intracellular calcium increase. Indeed, we and others have previously observed dysregulated calcium responses in iPSC-derived neural cells carrying *MT-ATP6* mutations^24,25^.

We dissected brain organoids on days 70-74 to prepare cortical brain organoid slices (cBOS)^54^ that we grew until day 129 and then treated with sildenafil for 24 hours before applying acute metabolic stress (2 minutes of glucose deprivation and inhibition of glycolysis and OXPHOS) **(Figure 4a)**. The calcium response to metabolic stress was more pronounced and premature in LS cBOS compared to control cBOS, suggesting an increased susceptibility to metabolic imbalance in the presence of *MT-ATP6* mutations **(Figure 4b-c)**. Pre-treatment with sildenafil in LS cBOS was sufficient to reduce their calcium load after metabolic stress and their peak calcium amplitude **(Figure 4b-c)**. The findings suggest that sildenafil may prevent excessive decompensation in LS neuronal cells under acute metabolic stress.

**Figure 4.**
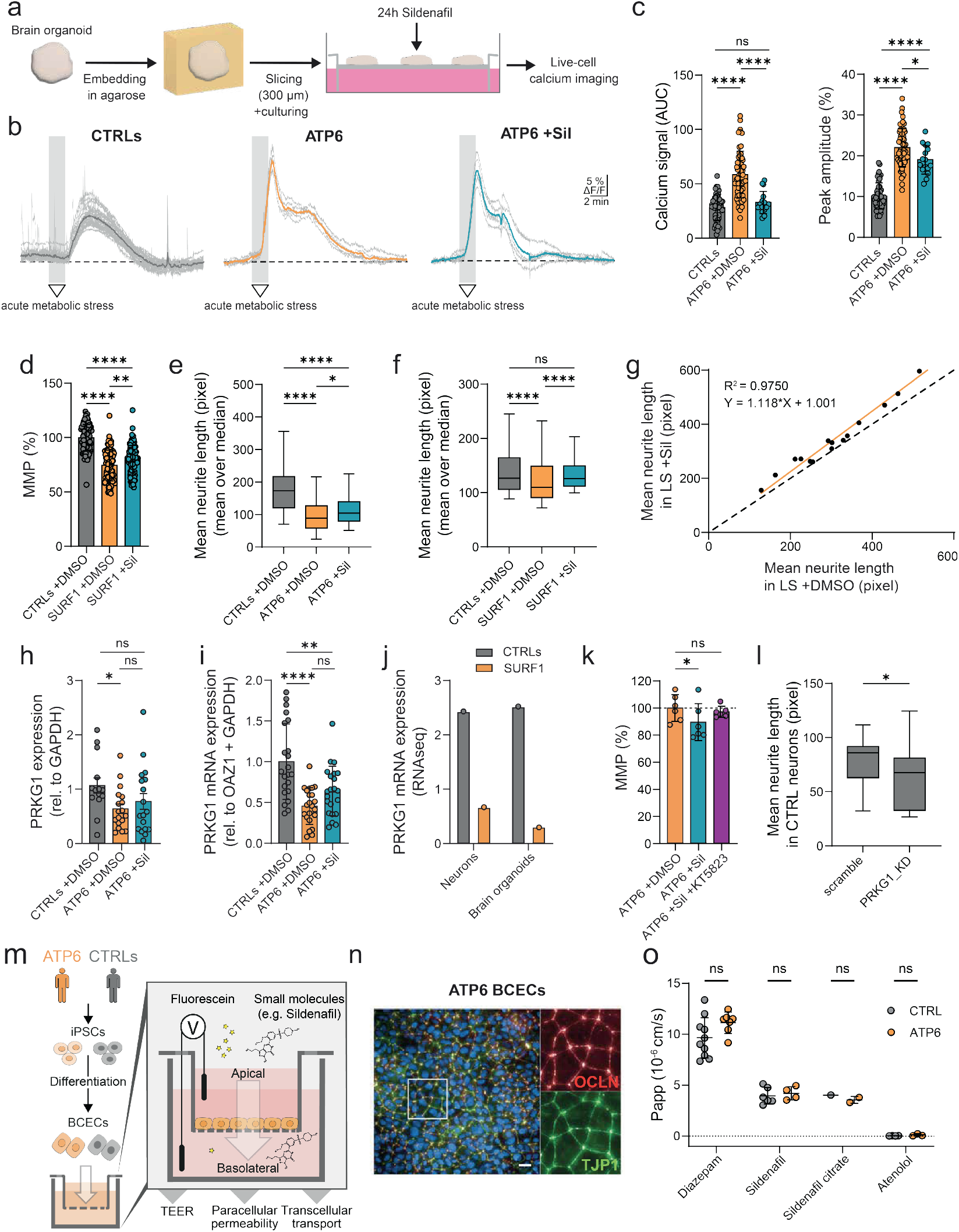
Sildenafil ameliorates calcium homeostasis and neurite growth and can cross the human BBB. **a**. Schematic of cortical brain organoid slices (cBOS) for live-cell calcium imaging. **b**. Exemplary traces of intracellular calcium in control cBOS (CTRL_2) and LS cBOS (ATP6_7) in response to acute metabolic stress (2 min of glucose-free medium + 2 mM 2-deoxyglucose and 5 mM sodium azide). Individual cells recorded in a single experiment (grey lines) plus corresponding average traces (colored). **c**. Area under the curve (AUC) and peak of calcium signals evoked by metabolic stress in control cBOS (CTRL_1, n=10; CTRL_2, n=40) and LS cBOS (ATP6_7, n=58; ATP6_7 +Sil, n=16). Dots represent individual cells within cBOS. *p<0.05, ****p<0.0001, ns (not significant); two-tailed Mann-Whitney U test. **d**. HCA-based MMP quantification in SURF1 mutant NPCs (SURF1_1) and respective isogenic control NPCs (CTRL_1). Dots represent mean values per image in n=3 independent experiments per line treated for 16 h with 10 *μ*M sildenafil or DMSO. **p<0.01, ****p<0.0001, ns; ordinary one-way ANOVA with Holm-Šídák’s multiple comparisons test. **e-f**. Quantification of neuronal outgrowth of day 16 dopaminergic-enriched neurons from MT-ATP6 mutants (ATP6_2, n=2 and ATP6_4, n=1) and controls (CTRL_1, n=2, CTRL_2, n=2 and CTRL_3, n=1), and from SURF1 mutants (SURF1_1, n=3) and isogenic controls (CTRL_1, n=3) treated for 8 days with 10 *μ*M sildenafil or DMSO. Dots represent the mean neurite length per image calculated as the mean over the median per each experiment. *p<0.05, ****p<0.0001, ns; Kruskal–Wallis test with Dunn’s multiple comparison test. **g**. Linear regression (sildenafil over DMSO) of the mean neurite length in LS neurons (ATP6_2, n=3; ATP6_4, n=1; ATP6_7, n=3; SURF1_1 n=3; NDUFS4_1, n=2). Black dotted line indicates the hypothetical linear regression in which no effect of the treatment would be seen (y=x or slope=1). Dots represent individual experiments. **h**. *PRKG1* expression quantified from n=5 independent experiments in control NPCs (CTRL_1, CTRL_2, CTRL_3, CTRL_4, CTRL_5) and LS NPCs (ATP6_2, ATP_4, ATP6_5, ATP6_7) treated for 16 h with 10 *μ*M sildenafil or DMSO. *p<0.05, ns (not significant); unpaired two-tailed t-test. **i**. qPCR-based gene expression of PRKG1 in day 70 cortical brain organoids from controls (CTRL_1, CTRL_2) and LS (ATP6_4, ATP6_7) treated for 24 h with 10 *μ*M sildenafil or DMSO. Data were normalized to housekeeping genes *OAZ1* and *GAPDH* and presented in relation to pooled controls. 5 brain organoids per sample out of n=3 independent experiments. **p<0.01****, p<0.0001, ns (not significant); two-tailed Mann-Whitney U test. **j**. *PRKG1* gene expression in SURF1 mutant neurons and brain organoids (SURF1_1) compared to isogenic controls (CTRL_1) based on published bulk RNAseq^23^. **k**. HCA-based MMP quantification in LS NPCs (ATP6_2) treated for 16 h with DMSO, 10 *μ*M sildenafil, or 10 *μ*M sildenafil plus pre-treatment for 1 h with 1 *μ*M of the PRKG1 inhibitor KT5823. *p<0.05, ns (not significant); one-way ANOVA. **l**. Quantification of neurite outgrowth in d16 dopaminergic-enriched neurons from controls (CTRL_1) exposed to two sequential small interfering RNA (siRNA) against PRKG1 (PRKG1_KD) or scrambled siRNA for 4 days. Dots represent mean values per well with 4-9 wells per experiment out n=3 independent experiments. *p<0.05; unpaired two-tailed t-test. **m**. Schematic of the iPSC-derived blood brain barrier (BBB) model to generate brain capillary endothelial cells (BCECs). **n**. Representative immunostaining of LS BCECs (ATP6_2) showing expression and co-localization of occludin (OCLN) and tight junction protein 1 (TJP1) required to form a barrier with high transendothelial electrical resistance (TEER). Scale bar: 100 *μ*m. **o**. Permeability coefficient (Papp) measured by LC-MS/MS in the apical basolateral media of control BCECs (CTRL_8) and LS BCECs (ATP6_2) exposed to 10 *μ*M sildenafil or sildenafil citrate for 1 h in n=1-5 independent experiments. Diazepam and atenolol (10 *μ*M each) served as internal positive and negative control, respectively. ns; two-way ANOVA.

Next, we carried out computational modeling to determine the implications of the increased cytosolic calcium increase in LS cBOS in response to metabolic stress and the modulatory effect of sildenafil **(Figure S8a)**. To model *MT-ATP6* defects, we computationally reduced the activity of ATP synthase of control cBOS and found that this resulted in an elevated calcium response, slight changes in NADH production, hyperpolarization of MMP, and reduction in cellular and mitochondrial ATP content **(Figure S8b)**. Consistently, the computational increase in ATP synthase activity of LS cBOS resulted in decreased calcium response, reduced NADH, mitochondrial hyperpolarization, and increased ATP concentration **(Figure S8c)**. This model suggested that the effect of sildenafil on calcium homeostasis may subsequently lead to changes in MMP, NADH levels, and ATP production. Indeed, we found that sildenafil modulated MMP **(Figure 1f)**, NAD+/NADH levels **(Figure 2e)**, and ATP production **(Figure 1g)**. The model raised the possibility that MMP normalization by sildenafil may not be due to a direct mitochondrial uncoupling effect. If this was the case, sildenafil might also prove beneficial in other forms of LS in which the MMP is depolarized rather than hyperpolarized. To test this hypothesis, we measured MMP in NPCs derived from LS iPSCs carrying a mutation in the nuclear gene *SURF1* and the corresponding isogenic control^23^ **(Figure S8j)**. The MMP was depolarized in SURF1-mutant NPCs compared to isogenic control NPCs **(Figure 4d)**. Nevertheless, sildenafil treatment led to MMP normalization in SURF1-mutant NPCs by increasing mitochondrial membrane polarization **(Figure 4d)**.

We then investigated the effect of sildenafil on neuronal outgrowth, as this process was highlighted by omics analyses of both NPCs **(Figure 2b, Figure S4a)** and brain organoids **(Figure S6f)**. We generated dopaminergic-enriched neuronal cultures **(Figure S8e)**, since LS particularly affects basal ganglia and dopaminergic neurons^55^. We quantified neurite outgrowth using HCA^56^. In agreement with previous findings^23,28^, MT-ATP6 mutant neurons and SURF1 mutant neurons exhibited reduced neurite length compared to control neurons **(Figure 4e-f, Figure S8f)**. Sildenafil treatment promoted effective neurite outgrowth in LS neurons **(Figure 4e-f, Figure S8f)**. This improvement was specific to LS neurons (carrying mutations in the genes *MT-ATP6, SURF1*, or *NDUFS4*) **(Figure 4g, Figure S8j)** and was not observed in control neurons treated with sildenafil **(Figure S8g)**.

Lastly, we focused on PRKG1, a PDE5 target that was downregulated in the progenitor population of LS brain organoids **(Figure 3g-h)** and was part of the sildenafil rescue signature of LS NPCs **(Figure 2g)**. PRKG1 is known to exert effects on both calcium homeostasis and neurite outgrowth,^57,58–61^ potentially explaining the mechanism of action that we observed in LS neural cells on these two pathways. PRKG1 levels were lower in MT-ATP6 mutant NPCs **(Figure 4h, Figure S8d)**, neurons **(Figure S8h)** and brain organoids **(Figure 4i)**, and SURF1 mutant neurons and brain organoids **(Figure 4j)**. Moreover, the PRKG1 inhibitor KT5823 blunted the effect of sildenafil on MMP normalization in LS NPCs **(Figure 4k)**. Knock-down of *PRKG1* using small interfering RNA (siRNA) in control iPSC-derived neurons recapitulated the neurite outgrowth defects seen in LS neurons **(Figure 4l)**. Hence, the effects of sildenafil in LS neural cells may be mediated by PRKG1 modulation **(Figure S8i)**.

### Sildenafil can cross the human blood-brain barrier and extends the lifespan of LS mice

In order for systemically administered sildenafil to exert potential beneficial effects in LS patients, the drug must cross the blood-brain-barrier (BBB). To probe the permeability for sildenafil in LS, we applied a human iPSC-based BBB model^62^ **(Figure 4m)**. LS brain capillary endothelial cells (BCECs) showed correct morphology and marker expression similarly to control BCECs **(Figure 4n, Figure S9a)**, as well as acceptable monolayer integrity and transendothelial electrical resistance (TEER) **(Figure S9b-c)**. The permeability of sildenafil and sildenafil citrate (the active pharmacological substance applied orally in clinical application) in LS BCECs was similar to that in control BCECs and was higher than the negative control atenolol (beta-blocker known to not permeate into the brain), but lower than the positive control diazepam (an anxiolytic benzodiazepine) **(Figure 4o)**. These results are consistent with permeability values of sildenafil reported in other clinical settings^63^. Thus, sildenafil may be able to effectively cross the BBB of patients with LS.

To address the therapeutic potential of sildenafil *in vivo*, we next employed the germline *Ndufs4* KO mouse, which is the most widely used animal model of LS showing motor regression, encephalopathy, cardiac conduction defect, and premature death^10,11^. Sildenafil was added to the drinking water of the animals after weaning starting on day 25 of life and continued throughout their life. This treatment significantly extended the lifespan of *Nduks4* KO mice **(Figure 5a)**. Sildenafil treatment attenuated the neurological decline of these mice by alleviating muscle weakness and ataxia, and by partially correcting the defective energy expenditure **(Figure 5b)**. Upon placing LS mice in metabolic chambers **(Figure S9f)**, we observed that sildenafil improved oxygen consumption and carbon dioxide production of LS mice **(Figure 5c)**, which may indicate enhanced metabolic fitness. Cardiac bradyarrhythmia and dysfunction were also ameliorated by sildenafil in *Ndufs4* KO mice, including restoration of heart rate and diastolic function **(Figure S9g)**.

**Figure 5.**
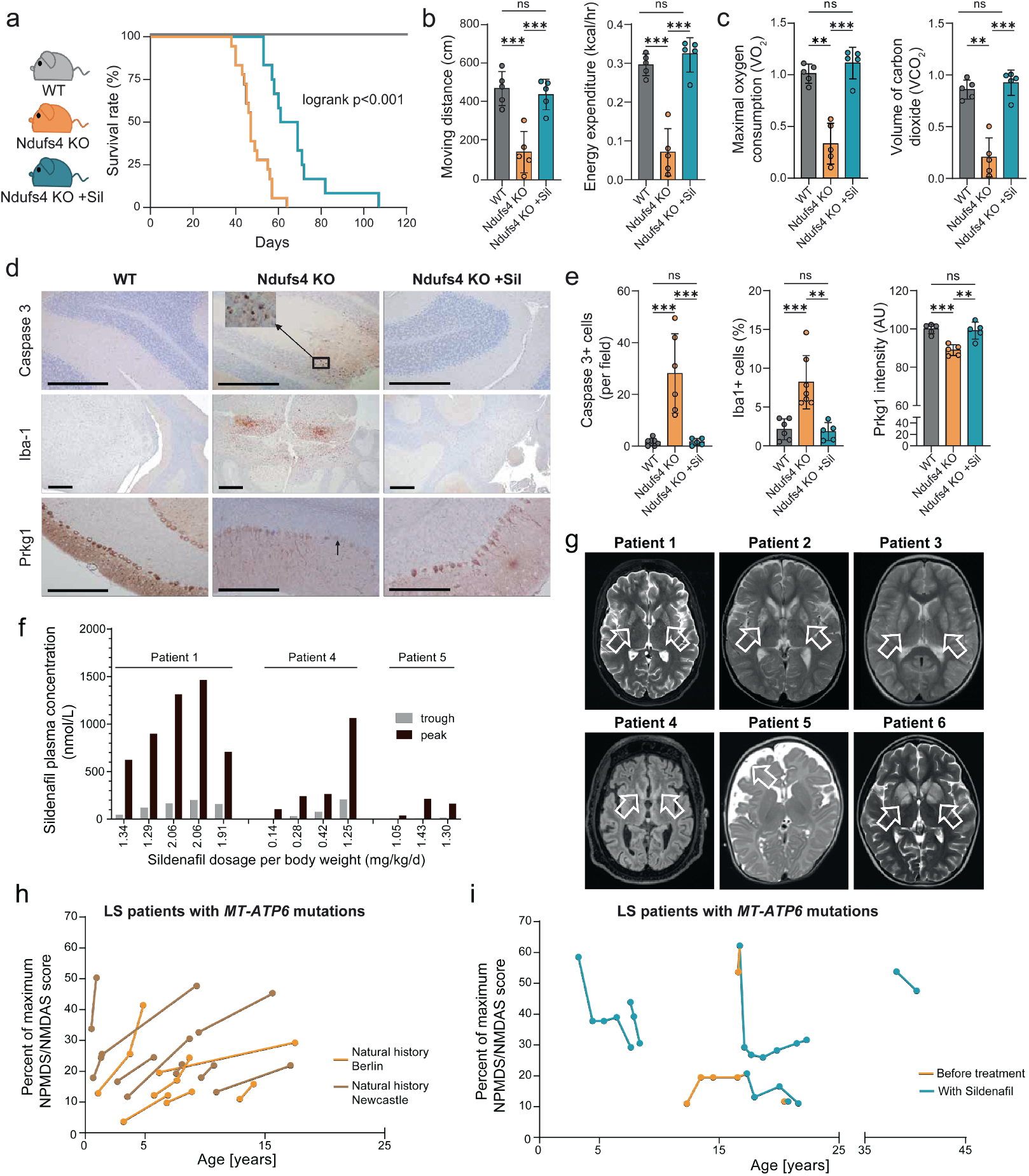
Sildenafil extends the lifespan of LS mice and improves clinical features in six patients with *MT-ATP6* mutations. **a**. Kaplan–Meier survival curve of untreated *Ndufs4* KO mice (10 females, 8 males, orange line) compared to *Ndufs4* KO mice exposed to sildenafil citrate added in the drinking water (6 females, 6 males, blue line). Logrank p<0.001. **b-c**. Functional parameters in untreated or sildenafil-treated 42-45-days-old *Ndufs4* KO mice (n=15 mice out of five independent experiments) collected by placing mice in a metabolic chamber (Figure S9a). **p<0.01, ***p<0.001, ns (not significant); ANOVA posthoc analysis. **d**. Representative immunohistochemistry showing cells positive for cleaved caspase3, Iba1 and Prkg1 in Purkinje cells in untreated or sildenafil-treated 50-days-old *Ndufs4* KO mice. **e**. Quantification of immunohistochemistry shown in d (n=5-6). **p<0.01, ***p<0.001, ns; ANOVA post-hoc analysis. **f**. Sildenafil plasma concentrations determined by LC-MS/MS at different daily oral dosages [in mg/kg/d total dose] divided into 3 single doses from three patients with LS undergoing off-label compassionate treatment with sildenafil. The trough level was determined in the morning and the peak level exactly one hour after application of the first morning dosage of sildenafil. For each patient, the depicted time points were separated by more than 3 months. **g**. Cranial magnetic resonance imaging (cMRI) of the six patients with LS undergoing off-label compassionate treatment with sildenafil. Arrows indicate lesions in the basal ganglia lesions and brainstem that are characteristic of LS (see supplementary methods for details). **h**. Clinical development of patients with LS carrying *MT-ATP6* mutations rated with the Newcastle mitochondrial disease adult scale (NMDAS) or Newcastle pediatric mitochondrial disease scale (NPMDS) plotted as percentage of the maximum reachable score for the respective age group. Brown lines indicate individuals previously described in a cohort from Newcastle^70^. Orange lines show individuals in our cohort from Charité Berlin^71^. **i**. Longitudinal clinical evaluation of off-label compassionate treatments with sildenafil in six patients with LS carrying different *MT-ATP6* mutations based on NMDAS/NPMDS score **(see Table 1 for patient details)**.

We and others have previously reported that the brain of *Ndufs4* KO mice exhibit increased apoptosis and higher levels of neuroinflammation^10,64,65^. Consistently, we observed positive cleaved caspase 3 staining in neuronal cells in the cerebellum and brain stem regions associated with increased Iba1 staining^66^ suggestive of microglial activation **(Figure 5d-e)**. Notably, the number of cells positive for caspase 3 or Iba1 significantly decreased in LS mice treated with sildenafil **(Figure 5d-e)**. The loss of Purkinje cells of cerebellum in LS mice was also ameliorated by sildenafil treatment **(Figure S9d-e)**. The expression of Prkg1, which is particularly evident in normal cerebellar Purkinje neurons^11^, was significantly reduced in LS mice and rescued by sildenafil treatment **(Figure 5d-e)**. These findings underscore Prkg1 as one of the mechanistic targets underlying the beneficial effect of sildenafil in delaying neuropathological deterioration. The improvement in neuropathology explains the less severe muscle weakness, reduced cerebellar ataxia, and prolonged survival of LS mice.

**Table 1.**
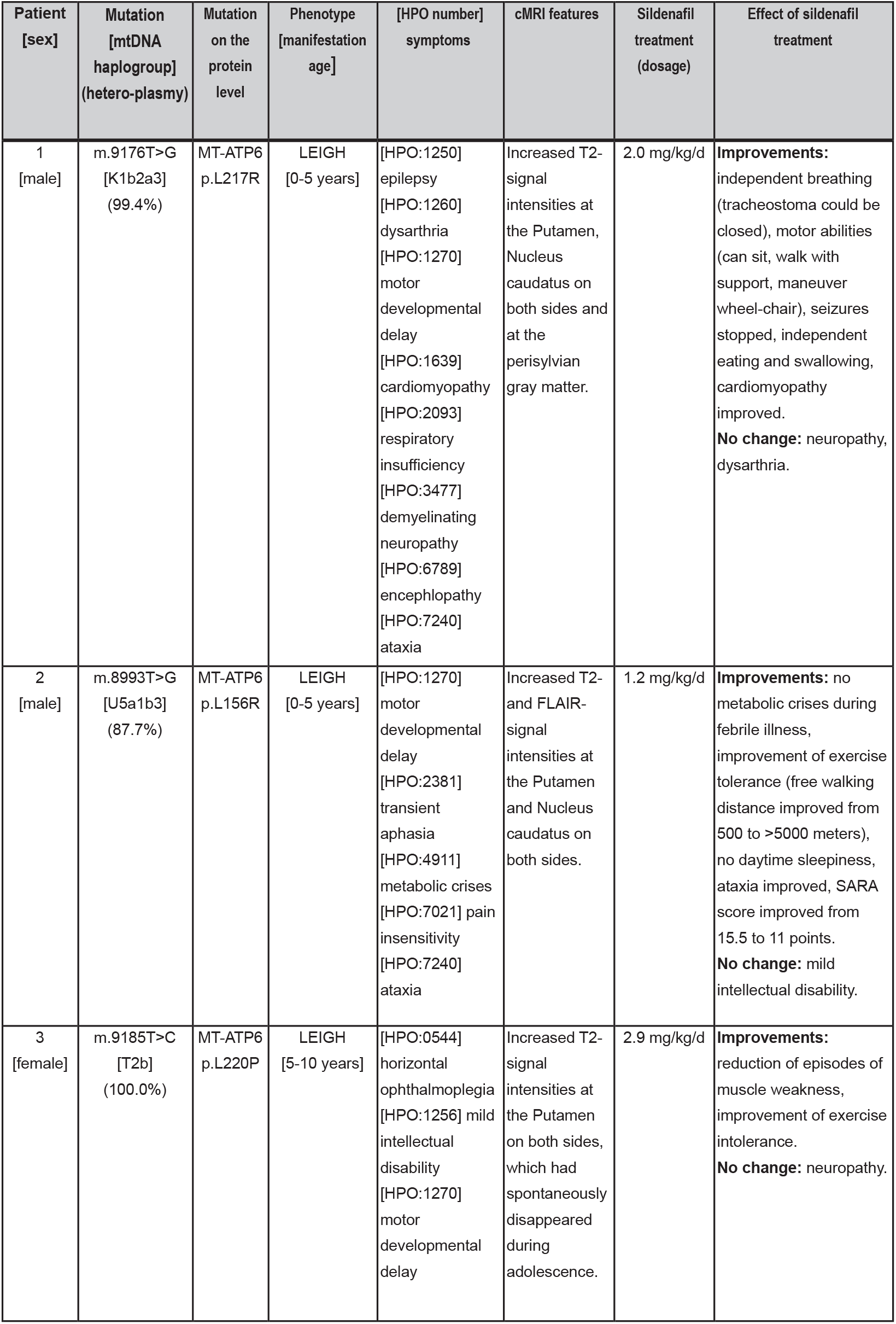
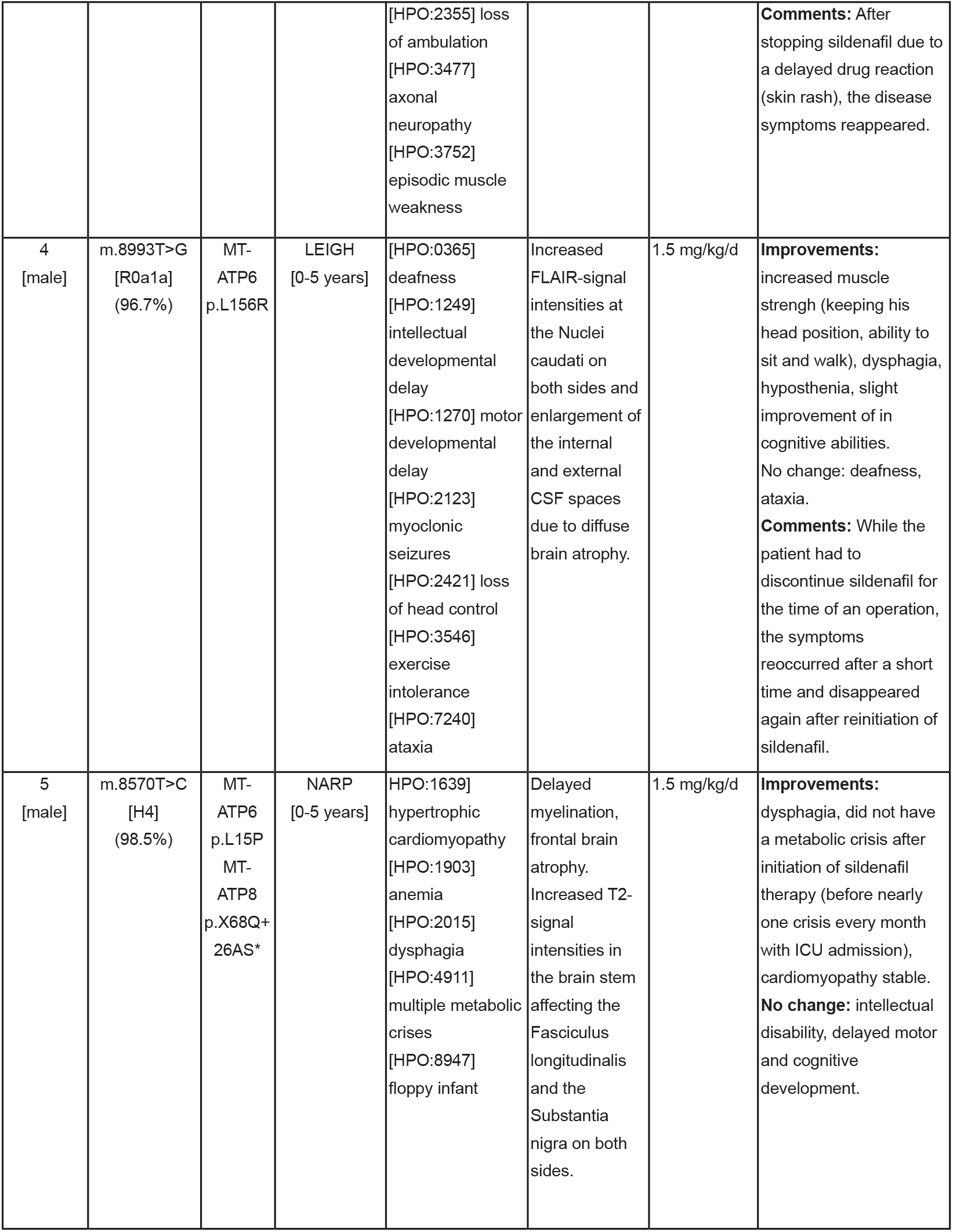
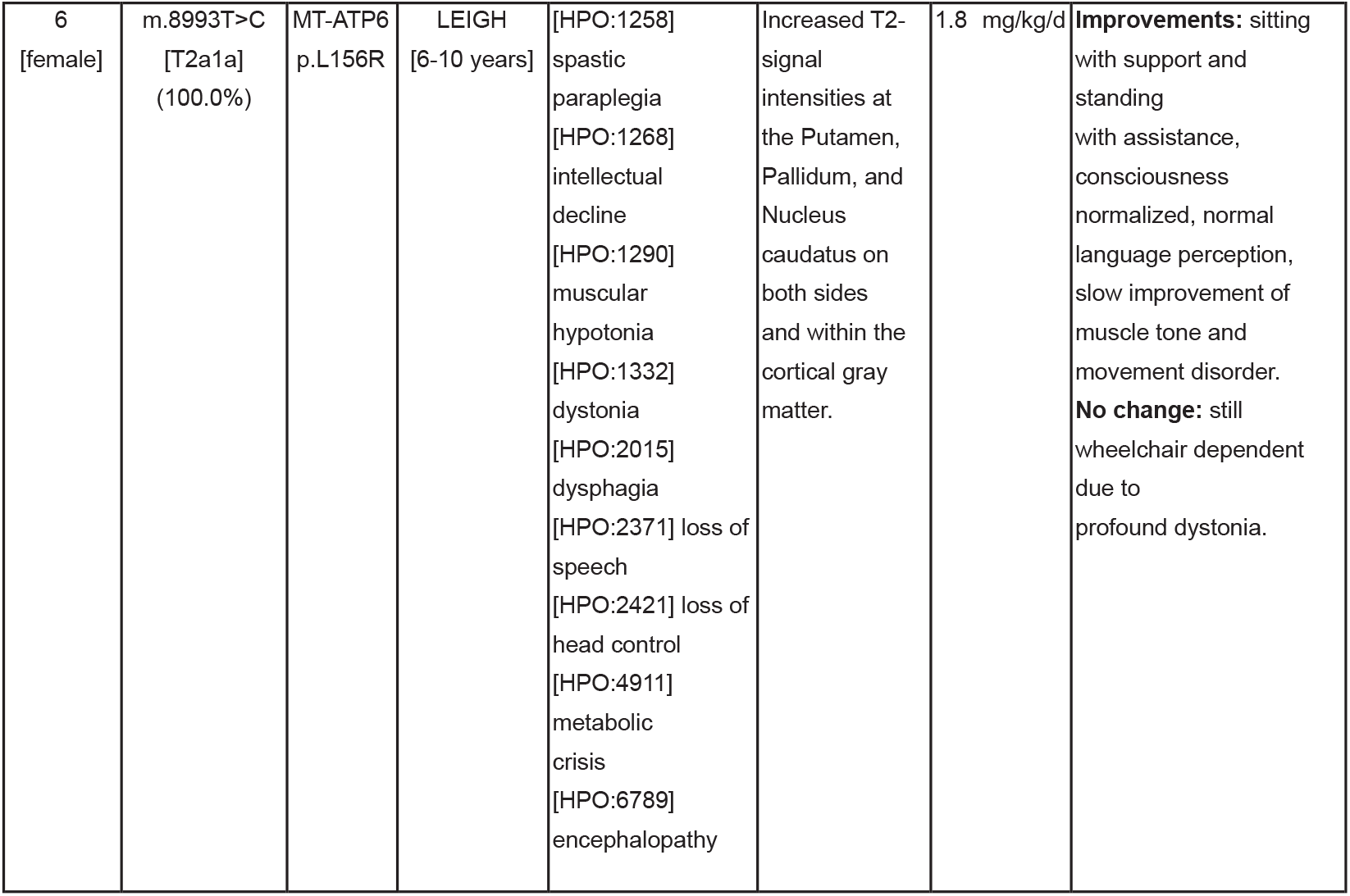
Clinical characteristics and response to sildenafil therapy in off-label compassionate studies in in six patients with LS carrying *MT-ATP6* mutations. NARP: Neuropathy, Ataxia, Retinitis pigmentosa; HPO: Human Phenotype Ontology; cMRI: cranial Magnetic Resonance Imaging; FLAIR: Fluid Attenuated Inversion Recovery; CSF: cerebrospinal fluid; SARA: Scale for the Assessment and Rating of Ataxia

| Patient<br>[sex] | Mutation<br>[mtDNA<br>haplogroup]<br>(hetero-plasmy) | Mutation<br>on the<br>protein<br>level | Phenotype<br>[manifestation<br>age] | [HPO number]<br>symptoms | cMRI features | Sildenafil<br>treatment<br>(dosage) | Effect of sildenafil<br>treatment |
| --- | --- | --- | --- | --- | --- | --- | --- |
| 1<br>[male] | m.9176T>G<br>[K1b2a3]<br>(99.4%) | MT-ATP6<br>p.L217R | LEIGH<br>[0-5 years] | [HPO:1250]<br>epilepsy<br>[HPO:1260]<br>dysarthria<br>[HPO:1270]<br>motor<br>developmental<br>delay<br>[HPO:1639]<br>cardiomyopathy<br>[HPO:2093]<br>respiratory<br>insufficiency<br>[HPO:3477]<br>demyelinating<br>neuropathy<br>[HPO:6789]<br>encephlopathy<br>[HPO:7240]<br>ataxia | Increased T2-signal<br>intensities at<br>the Putamen,<br>Nucleus<br>caudatus on<br>both sides and<br>at the<br>perisylvian<br>gray matter. | 2.0 mg/kg/d | **Improvements:**<br>independent breathing<br>(tracheostoma could be<br>closed), motor abilities<br>(can sit, walk with<br>support, maneuver<br>wheel-chair), seizures<br>stopped, independent<br>eating and swallowing,<br>cardiomyopathy<br>improved.<br>**No change:** neuropathy,<br>dysarthria. |
| 2<br>[male] | m.8993T>G<br>[U5a1b3]<br>(87.7%) | MT-ATP6<br>p.L156R | LEIGH<br>[0-5 years] | [HPO:1270]<br>motor<br>developmental<br>delay<br>[HPO:2381]<br>transient<br>aphasia<br>[HPO:4911]<br>metabolic crises<br>[HPO:7021] pain<br>insensitivity<br>[HPO:7240]<br>ataxia | Increased T2-and FLAIR-signal<br>intensities at<br>the Putamen<br>and Nucleus<br>caudatus on<br>both sides. | 1.2 mg/kg/d | **Improvements:** no<br>metabolic crises during<br>febrile illness,<br>improvement of exercise<br>tolerance (free walking<br>distance improved from<br>500 to >5000 meters),<br>no daytime sleepiness,<br>ataxia improved, SARA<br>score improved from<br>15.5 to 11 points.<br>**No change:** mild<br>intellectual disability. |
| 3<br>[female] | m.9185T>C<br>[T2b]<br>(100.0%) | MT-ATP6<br>p.L220P | LEIGH<br>[5-10 years] | [HPO:0544]<br>horizontal<br>ophthalmoplegia<br>[HPO:1256] mild<br>intellectual<br>disability<br>[HPO:1270]<br>motor<br>developmental<br>delay | Increased T2-signal<br>intensities at<br>the Putamen<br>on both sides,<br>which had<br>spontaneously<br>disappeared<br>during<br>adolescence. | 2.9 mg/kg/d | **Improvements:**<br>reduction of episodes of<br>muscle weakness,<br>improvement of exercise<br>intolerance.<br>**No change:** neuropathy. |
|  |  |  |  | [HPO:2355] loss of ambulation<br>[HPO:3477] axonal neuropathy<br>[HPO:3752] episodic muscle weakness |  |  | **Comments:** After stopping sildenafil due to a delayed drug reaction (skin rash), the disease symptoms reappeared. |
| 4<br>[male] | m.8993T>G<br>[R0a1a]<br>(96.7%) | MT-ATP6<br>p.L156R | LEIGH<br>[0-5 years] | [HPO:0365] deafness<br>[HPO:1249] intellectual developmental delay<br>[HPO:1270] motor developmental delay<br>[HPO:2123] myoclonic seizures<br>[HPO:2421] loss of head control<br>[HPO:3546] exercise intolerance<br>[HPO:7240] ataxia | Increased FLAIR-signal intensities at the Nuclei caudati on both sides and enlargement of the internal and external CSF spaces due to diffuse brain atrophy. | 1.5 mg/kg/d | **Improvements:** increased muscle strength (keeping his head position, ability to sit and walk), dysphagia, hyposthenia, slight improvement of in cognitive abilities.<br>**No change:** deafness, ataxia.<br>**Comments:** While the patient had to discontinue sildenafil for the time of an operation, the symptoms reoccurred after a short time and disappeared again after reinitiation of sildenafil. |
| 5<br>[male] | m.8570T>C<br>[H4]<br>(98.5%) | MT-ATP6<br>p.L15P<br>MT-ATP8<br>p.X68Q+26AS* | NARP<br>[0-5 years] | HPO:1639] hypertrophic cardiomyopathy<br>[HPO:1903] anemia<br>[HPO:2015] dysphagia<br>[HPO:4911] multiple metabolic crises<br>[HPO:8947] floppy infant | Delayed myelination, frontal brain atrophy. Increased T2-signal intensities in the brain stem affecting the Fasciculus longitudinalis and the Substantia nigra on both sides. | 1.5 mg/kg/d | **Improvements:** dysphagia, did not have a metabolic crisis after initiation of sildenafil therapy (before nearly one crisis every month with ICU admission), cardiomyopathy stable.<br>**No change:** intellectual disability, delayed motor and cognitive development. |

Table 1. Clinical characteristics and response to sildenafil therapy in off-label compassionate studies in six patients with LS carrying *MT-ATP6* mutations.
|  |  |  |  |  |  |  |  |
| --- | --- | --- | --- | --- | --- | --- | --- |
| 6<br>[female] | m.8993T>C<br>[T2a1a]<br>(100.0%) | MT-ATP6<br>p.L156R | LEIGH<br>[6-10 years] | [HPO:1258]<br>spastic<br>paraplegia<br>[HPO:1268]<br>intellectual<br>decline<br>[HPO:1290]<br>muscular<br>hypotonia<br>[HPO:1332]<br>dystonia<br>[HPO:2015]<br>dysphagia<br>[HPO:2371] loss of<br>speech<br>[HPO:2421] loss of<br>head control<br>[HPO:4911]<br>metabolic<br>crisis<br>[HPO:6789]<br>encephalopathy | Increased T2-signal<br>intensities at<br>the Putamen,<br>Pallidum, and<br>Nucleus<br>caudatus on<br>both sides<br>and within the<br>cortical gray<br>matter. | 1.8 mg/kg/d | **Improvements:** sitting<br>with support and<br>standing<br>with assistance,<br>consciousness<br>normalized, normal<br>language perception,<br>slow improvement of<br>muscle tone and<br>movement disorder.<br>**No change:** still<br>wheelchair dependent<br>due to<br>profound dystonia. |

### Compassionate sildenafil treatment improves clinical conditions in six LS patients

The promising preclinical findings of sildenafil in cellular and animal models of LS prompted us to ask whether this drug could be repurposed to treat patients with LS. Sildenafil has been used safely in several pediatric conditions including pulmonary hypertension and lymphatic malformations^37–40^. We therefore initiated off-label compassionate chronic oral application of sildenafil citrate (Revatio^®^) in six LS patients carrying different *MT-ATP6* mutations **(Table 1, Table S8, Figure S10)**. All patients had cranial magnetic resonance imaging (cMRI) signs consistent with basal ganglia lesions characteristic of LS **(Figure 5g, Figure S9h)**. Patient 5 was initially diagnosed with NARP (Neuropathy, Ataxia, Retinitis Pigmentosa). NARP and LS caused by *MT-ATP6* mutations are part of a disease continuum that is also referred to as “Leigh syndrome spectrum” disease^67^.

To determine that sildenafil was present in the blood of patients at appropriate concentrations, we performed mass spectrometry-based quantification of plasma samples collected in the morning and 1 hour after sildenafil administration **(Figure 5f)**. The drug reached a concentration of approximately 1 *μ*M in the blood. This concentration is close to the IC50 value of 3 *μ*M that was found to be effective for normalizing the MMP in LS NPCs **(Figure S3h)**. We also derived iPSCs from patient 1 (line ATP6_7), which we used to generate NPCs and brain organoids **(Figure S1e)**. Upon starting the treatment, we closely monitored the occurrence of potential adverse reactions. We asked patients and guardians to respond to a questionnaire on side effects after six months of sildenafil treatment **(Table S9)**. Sildenafil was generally well tolerated **(Table S9)** and continued in most patients **(Table 1, Table S8)**.

In all cases, we observed a clear improvement in different clinical parameters. This was particularly true for motor function, muscle strength, and resilience against metabolic crises **(Table 1, Table S8)**. Possible protection against metabolic crises was evident in patient 5, who experienced metabolic crises almost monthly before the initiation of the treatment. Mild improvements in cognitive abilities were reported in patients 4 and 6. A skin rash developed in patient 3 after 1.5 years of treatment. After discontinuing sildenafil, the rash disappeared but disease symptoms worsened again.

We monitored disease progression using the Newcastle mitochondrial disease adult scale (NMDAS)^68^ or the Newcastle pediatric mitochondrial disease scale (NPMDS)^69^. These rating scales cover all aspects of mitochondrial disease including clinical assessments and quality of life. The total score describes the overall severity of disease burden and typically increases over time in patients with mitochondrial diseases^68,69^. The NPMDS/NMDAS score can rise rapidly in patients with LS, especially following metabolic crises. This increase can be seen in the cohort of LS patients with *MT-ATP6* mutations previously described in Newcastle^70^, where the score increased by 8.8 SD 11.4 percent points per year **(Figure 5f**, brown lines), and in our cohort at Charité in Berlin^71^, where the score increased by 3.4 SD 2.2 percent points **(Figure 5f**, orange lines). Remarkably, the NPMDS/NMDAS scores of the six sildenafil-treated patients decreased or showed reduced rates of increase over time **(Figure 5g)**.

The results of the individualized compassionate studies suggest that chronic treatment with sildenafil may lead to clinical improvement in patients with LS carrying *MT-ATP6* mutations. A formal randomized controlled clinical trial is now required to evaluate the safety and potential efficacy of sildenafil in patients with Leigh syndrome.

## Discussion

Sildenafil is a well-studied molecule in clinic applications^30^. In addition to its widespread use in adult men with erectile dysfunction^72^, sildenafil is used in children to treat pulmonary hypertension and rare lymphatic malformation^37–40^. Recent studies suggest that sildenafil might be a repurposable candidate for treating disorders of the nervous system^73^. For instance, sildenafil use was associated with a reduced risk of developing Alzheimer’s disease (AD) and promoted neuronal outgrowth in iPSC-derived neurons from AD patients^74,75^. Sildenafil also lowered cognitive decline in mouse models of Huntington’s disease (HD)^76,77^ and increased motor and cognitive performances in patients with HD^78^. It also alleviated neuropathological sings in animal models of multiple sclerosis^79^ and ischemic stroke^80^. In this study, we found that sildenafil might slow down the neurological decline of the severe mitochondrial brain disease Leigh syndrome (LS).

Sildenafil is a phosphodiesterase 5 inhibitor (PDE5i) leading to the release of cyclic guanosine-3′,5′-monophosphate (cGMP), which in turn has a wide range of functions, likely contributing to a poly-pharmacological mode of action^30^. In the context of the nervous system, sildenafil may enhance neurogenesis in mice^81,82^. Consistently, we found that the signature of sildenafil primarily involved the regulation of nervous system development and, within brain organoids, acted specifically on progenitors and radial glia populations. Mechanistically, the second messenger cGMP and the cGMP-dependent protein kinase 1 (PRKG1) can modulate neurotransmitter release, neuronal survival, growth cone guidance, and axon/dendrite formation^57,58–60^. Mice deficient for Prkg1 showed substantial impairment in axon guidance and connectivity^61^. In agreement with previous findings^23,28^, we observed that human neurons carrying mutations causative of LS exhibited impaired neuronal outgrowth. We found that this feature was associated with lower PRKG1 levels, and that knock-down of PRKG1 in healthy neurons was sufficient to recapitulate the branching defects. PRKG1 expression was also reduced in LS human brain organoids and in cerebellar Purkinje cells of LS mice. In mouse brain, Prkg1 is known to be highly expressed in Purkinje cells, where its selective depletion result in specific motor learning defects^83^. Here, we found that treatment with sildenafil rescued Pkgk1 levels in LS mice *in vivo*. In human LS neurons, sildenafil enhanced neurite outgrowth capacity. These findings, together with the ability of the drug to cross the BBB, suggest that sildenafil may be beneficial in promoting neuronal morphogenesis and connectivity in the brain of patients with LS.

In addition to its neurological impact, sildenafil modulated bioenergetic processes *in vitro* and improved metabolic fitness of LS mice *in vivo*. Sildenafil is known to induce mitochondrial biogenesis and oxygen consumption^84,85^, and PRKG1 has been suggested to be a regulator of energy homeostasis^86^. This is supported by our findings showing that sildenafil failed to normalize the MMP of LS neural cells when PRKG1 was inhibited. Sildenafil also reversed changes in mitochondrial crista junctions distributions that may reflect defects of cristae morphology caused by *MT-ATP6* mutations^41^. We postulate that the bioenergetic improvement of sildenafil may be related to the modulation of ion homeostasis. We have previously reported that PDE5i ameliorated calcium defects in LS NPCs^25^. Here, we found that pre-treatment of LS brain organoids with the PDE5i sildenafil reduced their abnormal calcium response to acute metabolic stress. It is tempting to speculate that this effect may also be related to PRKG1, given its known role in mediating neuronal calcium signalling^87,88^. Whether these functions of sildenafil are linked to its vasodilatory effects^30^ remains to be studied. The increase in blood flow and oxygen to muscle and brain tissue may potentially contribute to beneficial metabolic effects. Indeed, sildenafil led to vascular and metabolic improvements in individuals with AD^89^ and in subjects with vascular cognitive impairment due to cerebral small vessel disease^90^. The known effect of sildenafil on the vasculature^30^ and the beneficial consequences of hypoxia seen in animal models of LS^20,21^ might indicate that modulation of brain vascularization and associated oxygenation^91^ could represent important therapeutic targets in LS.

Nonetheless, our findings underscore the power of conducting drug discovery studies driven by cellular reprogramming and iPSC models. Similar strategies could be applied to other rare neurological diseases for which effective model systems are lacking. Finally, because mitochondria play a crucial role in cellular health and brain function and homeostasis^92–94^, understanding the potential effect of PDE5 inhibitors in mitochondrial diseases may also have important implications in the context of other more common aging-related neurological disorders.

## Limitations of the study

Our pre-clinical and clinical data suggest that sildenafil may prevent or delay the progression of LS. Compassionate treatment with sildenafil in six LS patients carrying *MT-ATP6* mutations resulted in substantial improvements in motor and functional aspects, overall quality of life, and resilience against metabolic crises. The reduction of the upward sloping trend in NPMDS/NMDAS scores over time possibly implies that sildenafil treatment modified the disease trajectory. However, it is important to highlight that we only report the clinical effects observed in a small cohort of mitochondrial disease patients. Therefore, it is imperative that the safety and efficacy of sildenafil in patients with Leigh syndrome should be assessed in interventional phase II and III randomized controlled clinical trials.

## Supporting information

Resource Table

## Data Availability

There are restrictions to the availability of the
patient derived iPSC lines used in this study
due to nature of our ethical approval that does
not support sharing to third parties without a
specific amendment and does not allow to
perform genomic studies in respect to the
European privacy protection law. All omics
datasets have been deposited in international
repositories and are availiable upon reasonable request to the authors

## Acknowledgments

We acknowledge financial support from the Deutsche Forschungsgemeinschaft (DFG) (PR1527/13-1 to A.P., PR1527/14-1 to A.P., RO5380/1-1 to A.R., and FOR 2795 “Synapses under Stress”: Ro2327/13-2 to C.R.R and PR1527/6-1 to A.P.), the German Excellence Strategy (EXC-2049-390688087) through the NeuroCure Consortium at Charité-Universitätsmedizin Berlin (to M.S.), the SPARK program at the Berlin Institute of Health (BIH) (to A.P. and M.S.), the European Joint Programme for Rare Diseases (EJPRD) and Bundesministerium für Bildung und Forschung (BMBF) (CureMILS, #01GM2002A to A.P., #01GM2002B to O.P., and to W.K., A.D-S., P.L., E.B.; mitoNET, #01GM1906A to T.K., www.mitoNET.org; GENOMIT #01GM1920A to T.K., www.GENOMIT.eu, and the German Center for Child and Adolescent Health to M.S.), and the European Commission’s Horizon Europe Programme (SIMPATHIC #101080249 to A.P., M.S., W.K.), the United Mitochondrial Disease Foundation (UMDF) (to A.P., D-F.D), the Leigh Syndrome International Consortium (LSIC) (to A.P), People Against Leigh syndrome (PALS) (to A.P), Foundation Maladies Rare and Association (AMMi) (to A.P), Cure Mito and Cure ATP6 (to A.P.), MitoHelp (to A.P. and M.S.), Mito Foundation (to A.P.), Mitocon Italy (to A.P.), the Medical Faculty of Heinrich Heine University (FoKo project to A.P.), AFM-Téléthon (#25179 to A.R.), North Rhine-Westphalia and European Union (IN-SU-3-001 to A.R.), the European Union (FETPROACT-2018-2020 HERMES #824164 to I.D.), the Eva Luise Köhler Foundation (to A.P., M.S.), the National Recovery and Resilience Plan (NRRP), Mission 4, Component 2, Investment 1.1, Call for tender No. 104 published on 2.2.2022 by the Italian Ministry of University and Research (MUR), funded by the European Union – NextGeneration EU– Project CUP B53D23018710001, Grant Assignment Decree No. n. 1110 adopted on 20/07/2023 by the Italian Ministry of University and Research (MUR) (to E.B.), Fondazione Telethon (#GSA23F002 to E.B., #GMR23T2152 to D.B.), the Italian Foundation A.M.Me.C. (to E.B.), Fondazione Regionale per la Ricerca Biomedica (Regione Lombardia-FRRB) (#1740526 to D.B.), and the Italian Ministry of Health (RRC). We acknowledge technical support from Dr. Francesca Griggio (Genomics and Transcriptomics Platform), Dr. Samantha Solito and Dr. Giulia Finotti (Flow Cytometry and Cellular Analysis Platform) of “Centro Piattaforme Tecnologiche” (CPT) of the University of Verona, Italy. We also thank Dr. Francesco De Sanctis and Dr. Annalisa Adamo, University of Verona, Italy for support with flow cytometry data analysis. S.H. was supported by a fellowship from the Jürgen Manchot Stiftung. V.T., C.L.M., V.C., and M.S. are members of the European Reference Network for Rare Neuromuscular Diseases (ERN EURO-NMD). During this study, A.S. was funded by the Research Council of Finland, the Jane and Aatos Erkko foundation, the PolG foundation, and the Sigrid Jusélius foundation.

## Author contributions

Conceptualization, A.P., M.S., O.P., D-F.D., E.B., W.K., A.D-S., A.S., P.L., F.E.; Methodology, A.Z., A.W., M-T.H., G.P., S.H., C.J., L.P., M.S-C., J.B., C.B., L.E., A.D.D., F.S., D.M., A.R-W., U.H., L.U., A.D-F., I.T., D.B., D.H., G.C., S.W., D-F.D., C.M., L.S., T.K., M.S., J.K-F., D.S., A.Za., U.H., C.S., E.C., M.P-G., S.M-G., C.L-M., F.D.; Formal Analysis, G.S.A., T.M.P., S.N., A.F., J.R., A.R.U., M.S.. A.Zh., O.B., D.M., M.M., A.M.; Resources, E.M., V.C., V.T., A.R., N.R., P.L., M.S., B.K., I.D., S.J.; Writing –Original Draft, A.P.; Writing – Review & Editing, A.P., O.P., M.S., D-F.D., E.B., C.R.R., L.P.; Supervision, A.P., A.D.S., O.P., C.R.R., I.S., M.J.W.A-H., E.M., S.J., G.U., C.R.R., S.P., W.K., M.J.W.A-H., A.S., F.E., E.B., M.S., L.C., V.C., S.J., A.D-S., D.B.; Visualization, A.Z., A.W., O.P., A.P.,M-T.H., G.P., S.H., M.S.; Funding Acquisition, A.P., A.D-S., O.P., W.K., P.L., A.S., F.E., E.B., M.S., C.R.R.

## Resource availiability

### Lead contact

Further information and requests for resources and reagents should be directed to and will be fulfilled by the lead contact, Alessandro Prigione, MD, PhD.

### Material availiability

There are restrictions to the availability of the patient-derived iPSC lines used in this study due to nature of our ethical approval that does not support sharing to third parties without a specific amendment and does not allow to perform genomic studies in respect to the European privacy protection law.

### Data and code availiability

All omics datasets have been deposited in international repositories.Raw and processed RNA sequencing data have been deposited in the NCBI Gene Expression Omnibus (GEO). The dataset includes raw FASTQ files, gene expression matrices, and associated metadata. The mass spectrometry data have been deposited in the ProteomeXchange Consortium (http://proteomecentral.proteomexchange.org) via the PRIDE partner repository^95^. The metabolomics dataset has been deposited in the MassIVE repository (https://massive.ucsd.edu/).

## Experimental model and study participant details

We obtained written informed consent to use patient material and health data from the patients and their guardians according to the Declaration of Helsinki. The use of iPSCs was approved by the local ethic committees of Charité-Universitätsmedizin Berlin (EA2/131/13 and EA2/107/14) and Heinrich Heine University Düsseldorf (Study number 2020-967_5). Off-label compassionate treatments were conducted under the framework of ClinicalTrial.gov identifier NCT06967831. Patient data were pseudoanonymized. Age ranges were used instead of actual age to avoid the risk of identifying individuals. Animal experiments were approved by the Institutional Animal Care and Use Committee (IACUC) of Johns Hopkins University.

MT-ATP6 mutant iPSC lines were previously derived from three patients with m.9185T>C mutation, one patient with m.8993T>C mutation, two patients with m.8993T>G mutation, and two patients with m.9176T>G mutation **(Figure S1e)**^25,31–33^. SURF1 iPSC isogenic lines and NDUFS4 iPSC isogenic lines were previously derived **(Figure S8j)**^23,66^. Among healthy control iPSCs **(Figure S1e)**, CTRL_1, CTRL_2, CTRL_4, CTRL_7 were previously described^25,31,95^, CTRL_3 (CRMi003-A) was purchased from RUCDR Infinite Biologics, CTRL_5 (HVRDi004-B-1, PGP1) was purchased from Synthego, CTRL_6 (IUFi004-A, iPS12) was purchased from Cell Applications, and CTRL_8 (WISCi004-B) was purchased from WiCell. We cultured all iPSC lines on Matrigel (Corning)-coated plates using StemMACS iPS-Brew XF medium (Miltenyi Biotec), supplemented with MycoZap (Lonza). CTRL_8 and ATP_2 iPSCs were cultured in mTeSR Plus Medium (STEMCELL Technologies) prior to BCEC differentiation. We cultivated all iPSCs in a humidified atmosphere of 5 % CO_2_ at 37 °C and 5 % oxygen. iPSCs were passaged at 70-80 % confluence with 0.5 *μ*M EDTA (Invitrogen) in 1 x PBA (Gibco). We added 10 *μ*M ROCK inhibitor (Enzo Biochem Inc) after splitting to promote survival. Cultures were routinely monitored for mycoplasma contamination by PCR using nine primers to amplify the six most common mycoplasma strains **(Resource Table)**. The positive control and internal control were kindly provided by Dr. Cord Uphoff (DZMS, Germany). We monitored the identity of iPSCs using STR analysis, which was performed by the Forensic Department of the University Hospital Düsseldorf (Dr. phil. nat. Petra Böhme); 21 microsatellite loci were amplified using PCR and tagged products were analyzed using GeneMapper ID v.3.2.1 (Applied Biosystems).

## Method details

### Neural progenitor cells (NPCs) and differentiated neurons

We obtained NPCs using a previously published protocol^97^. Briefly, we detached iPSCs from Matrigel-coated plates using Accutase (Sigma-Aldrich) and transferred the collected cells into low-attachment 6-well plates where they were kept for two days in: KnockOut-DMEM (Thermo Fisher Scientific), KnockOut-SR (Thermo Fisher Scientific), non-essential amino acids (NEAA) (Thermo Fisher Scientific), plus 1 mM pyruvate (Thermo Fisher Scientific) 2 mM L-glutamine (Thermo Fisher Scientific), 1 x MycoZap Plus-CL (Lonza) with the addition of 0.5 *μ*M purmorphamine (PMA) (Merck Millipore), 3 *μ*M CHIR 99021 (Cayman Chemical), 10 *μ*M SB-431542 (Selleckchem), and 1 *μ*M dorsomorphin (Sigma-Aldrich). From day 2 to day 4, the media was switched to: Neurobasal:DMEM/F12 [1:1], 0.5 x N2, 0.5 x B27 without vitamin A, 1 x MycoZap Plus-CL, with the addition of 0.5 *μ*M purmorphamine (PMA) (Merck Millipore), 3 *μ*M CHIR 99021 (Cayman Chemical), 10 *μ*M SB-431542 (Selleckchem), and 1 *μ*M dorsomorphin (Sigma-Aldrich). On day 6, we transferred the suspended cells onto Matrigel-coated well plates using: the same media without SB-431542 and dorsomorphin, but with the addition of 150 *μ*M ascorbic acid (Sigma-Aldrich). We maintained NPCs on this media and used them for experiments between passage 7 and passage 30.

Differentiation into neuronal cultures containing dopaminergic neurons was performed as previously described^23^. Starting from NPCs at low confluence (10-30 %), we switched to media containing Neurobasal:DMEM/F12 (1:1), 0.5 x N2, 0.5 x B27 with vitamin A, 1 x MycoZap Plus-CL, with the addition of 200 mM vitamin C, 100 ng/ml FGF8 (R&D Systems) and 1 mM PMA. After 8 days, we used the media Neurobasal:DMEM/F12 (1:1), 0.5 x N2, 0.5 x B27 with vitamin A, 1 x MycoZap Plus-CL, supplemented with 200 mM vitamin C, 0.5 mM PMA, 500 mM cAMP (STEMCELL Technologies), 10 ng/mL BDNF (MACS Miltenyi), 10 ng/ml GDNF (MACS Miltenyi) and 1ng/mL TGFbeta3 (MACS Miltenyi). After ten days, the medium was changed to Neurobasal:DMEM/F12 (1:1), 0.5 x N2, 0.5 x B27 with vitamin A, 1 x MycoZap Plus-CL, supplemented with 200 mM vitamin C, 500 mM cAMP, 10 ng/ ml BDNF, 10 ng/ml GDNF, and 1 ng/mL TGFbeta3.

### Cortical brain organoids

Cortical brain organoids were generated following two protocols **(Figure S6a)**. The first protocol was based on our modifications^47^ of a previous publication^98^. Briefly, at day 0, iPSCs of 80 % confluence were seeded into low-attachment U-bottom 96-well plates (Corning) to induce neurosphere formation. iPSCs were washed with PBS (Gibco) and detached with Accutase (Gibco) for 5 min at 37 °C. Cell pellets were resuspended in cortical differentiation medium I (CDMI) consisting of Glasgow-MEM, 20 % Knockout Serum Replacement, MEM-NEAA, sodium pyruvate, 2-mercaptoethanol and penicillin-streptomycin (all from Gibco). After cell counting, the suspension was diluted with CDMI to a final cell concentration of 90,000 cells per ml and supplemented with 20 μM ROCK inhibitor (Enzo), 5 μM TGF-β inhibitor SB431542 (Cayman Chemical Company) and 3 μM WNT antagonist IWR1 (EMD Millipore Corp). The supplemented seeding suspension was distributed to low attachment U-bottom 96-well plates by adding 100 μl per well and incubated at 37 °C and 5 % CO_2_. On day 3, the sides of the 96-well plate were carefully tapped to detach dead cells, and then 100 μl of CDMI supplemented with 20 μM ROCK inhibitor, 3 μM IWR1 and 5 μM SB431542. On day 6, 9, 12 and 15, 80 μl of the supernatant medium were removed from each well and replaced with 100 μl of CDMI supplemented with 3 μM IWR1 and 5 μM SB431542 per well. On day 18, the developing organoids were transferred to 100 mm petri dishes filled with cortical differentiation medium II (CDMII) consisting of DMEM/ F12, 1 % GlutaMAX, 1 % N2 supplement, 1 % chemically defined lipid concentrate and penicillin-streptomycin (Gibco). The petri dishes were placed on an orbital shaker at 70 rpm at 37 °C and 5 % CO_2_. CDMII was changed every second day. On day 35, CDMII was replaced with cortical differentiation medium III (CDMIII) that in addition contains 10% FBS (Gibco) and heparin (Merck). CDMIII was changed every two to four days, depending on the colour of the medium. From day 70 on, organoids were kept in cortical differentiation medium IV (CDMIV) that additionally was supplemented with 1 % B27 with vitamin A.

The second protocol was based on a different publication^48^. For this, AggreWell plates (STEMCELL Technologies) were prepared by incubation with Anti-Adherence Rinsing Solution (STEMCELL Technologies) for 15 min. iPSCs were detached from the 6-well plates using Accutase as described above. The cell suspension was pelleted at 200 x g for 4 min and resuspended in iPS Brew (Miltenyi Biotec, Germany) with 10 *μ*M ROCK inhibitor. 2.75 million cells were seeded in 1.5 ml per AggreWell. AggreWell plates were centrifuged at 100 x g for 3 min and left at 37 °C and 5 % CO_2_ for 24 h. At day 1, spheroids were transferred to a petri dish using a cut P1000 tip and placed on a shaker. Medium was changed to embryonic stem (ES) medium composed of KO-DMEM, 20% Knockout Serum Replacement, MEM-NEAA, sodium pyruvate, GlutaMAX, penicillin-streptomycin (all from Gibco), and MycoZAP Plus-CL (Lonza), and supplemented with 10 *μ*M SB431542 and 2.5 *μ*M dorsomorphin (Sigma). Until day 6, ES medium was changed daily. Neurospheres were fed with Neural Differentiation Medium (NDM) consisting of Neurobasal A medium (Gibco), GlutaMAX, penicillin-streptomycin, MycoZAP and 2 % B27 without vitamin A (Gibco). From day 6-15, medium was changed daily and supplemented with 20 ng/ml EGF (R&D Systems) and 20 ng/ml FGF2 (R&D Systems). From day 16-21, medium was changed every second day, and from day 22-45, NDM medium was supplemented with 20 ng/ml BDNF (MACS Miltenyi), 20 ng/ml NT-3 (Peprotech), 200 *μ*M (+)-sodium L-ascorbate (Sigma-Aldrich), 50 *μ*M dibutyryl cAMP (STEMCELL Technologies), and 10 *μ*M DHA (Sigma), and changed every other day. For long-term maturation after day 46, NDM medium was prepared with 2 % B27 with vitamin A, and organoids were fed according to their needs. Images of brain organoids were acquired using the Eclipse Ts2 light microscope (Nikon), and at later growth stages, when the organoids were larger, using a DMS1000 microscope (Leica). The size of the organoids was determined by area calculation using ImageJ.

### Bulk RNA sequencing

For bulk transcriptomics of NPCs, we used four MT-ATP6 mutant NPC lines (ATP6_2, ATP6_4, ATP6_5 and ATP6_7.) and four healthy control NPC lines (CTRL_1, CTRL_2, CTRL_3 and CTRL_4) either treated for 16 h with 0.1 % of DMSO alone or with 10 *μ*M sildenafil resuspended in 0.1 % DMSO. We used n=3 biological replicates per condition. Total RNA was isolated from pelleted NPCs using the NucleoSpin RNA Plus kit (Macherey Nagel) and eluted in 30 *μ*l RNase-free H2O. RNA concentration was measured with NanoDrop 2000. For quality check, 300 ng RNA was run on a 2 %TBE gel.

For bulk transcriptomics of brain organoids, total RNA was isolated from day 70 cortical brain organoids generated using the first protocol^47,98^. We used five pelleted organoids per sample with n=3 biological replicates per condition, grown under normal conditions (CTRL_1, CTRL_2, ATP6_4 and ATP6_7), or treated with 10 μM sildenafil for 24 hours (ATP6_4 and ATP6_7). Total RNA was isolated using the RNeasy Mini Kit by Qiagen. RNA concentration as well as purity was measured at the Nanodrop Spectrophotometer ND1000 (peQlab) using ND-1000 software (V3.8.1). Total RNA was mixed with 1 μg of a DNA oligonucleotide pool comprising 50-nt long oligonucleotide mix covering the reverse complement of the entire length of each rRNA (28S rRNA, 18S rRNA, 16S rRNA, 5.8S rRNA, 5S rRNA, 12S rRNA), incubated with 1U of RNase H (Hybridase Thermostable RNase H, Epicentre), purified using RNA Cleanup XP beads (Agencourt), DNase treated using TURBO DNase rigorous treatment protocol (Thermo Fisher Scientific) and purified again with RNA Cleanup XP beads. rRNA-depleted RNA samples were further fragmented and processed into strand-specific cDNA libraries using TruSeq Stranded Total LT Sample Prep Kit (Illumina) and sequenced using the NovaSeq 6000 system with stranded technology, generating paired-end reads of 150 bp. Raw RNA sequencing data were processed to generate FASTQ files using Illumina bcl2fastq. Briefly, base calling was performed using Real-Time Analysis (RTA) software, and adapter sequences were trimmed. The resulting BCL files were converted to FASTQ format, followed by quality control assessment using FastQC before downstream analysis.

The preprocessing of the FASTQ raw sequencing files was performed with nfcore/rnaseq (version 3.12.0) pipeline. Briefly, reads were trimmed with Trim Galore! (version 0.6.10) and mapped to the human genome (GRCh38 assembly) using STAR (version 2.7.10a) aligner. We performed different comparisons: i) LS with DMSO vs controls with DMSO (disease signature), ii) LS + sildenafil vs LS + DMSO (sildenafil signature), iii) controls + sildenafil vs controls + DMSO (negative control test). Transcript counts were determined using Salmon (version 1.10.1). DESeq2 (version 1.40.2)^99^ was used to identify differentially expressed genes in the NPC samples (Table S1) or in brain organoid samples (Table S4) (Benjamini-Hochberg-adjusted adjusted p-value £0.05). Gene Ontology (GO) enrichment analysis was performed using the enrichGO function within ClusterProfiler (4.8.3), using the annotations of org.Hs.eg.db (version 3.18.0). Visualizations were generated with ggplot2 (version 3.5.1).

### Single-nucleus RNA sequencing

For single-nucleus RNA sequencing (snRNAseq), we used day 72 brain organoids generated with the second protocol^100^ from control iPSCs (CTRL_1) and LS iPSCs (ATP6_7). Brain organoids were treated with either DMSO or 10 *μ*M sildenafil resuspended in DMSO for 45 days from day 27 to day 72. We used 15 pelleted organoids per sample (n = 8 samples) with n=2 biological replicates per condition (n = 120 cortical brain organoids in total). snRNA-seq was performed using the 10X Genomics Chromium system, following the manufacturer’s protocol. Nuclei were isolated using mechanical dissociation with gradient centrifugation and stained with DAPI to assess nuclear integrity. Barcoding and cDNA amplification were carried out using the Chromium Single Cell 3’ (vNext) Reagent Kit. Libraries were prepared and sequenced on the Illumina NovaSeq 6000, generating paired-end reads of 100 bp. Data processing, including demultiplexing, alignment to the reference genome (GRCh38-2020-A), and gene quantification, was performed using Cell Ranger (v7.2.0) from 10x Genomics with default parameters. To generate digital gene expression (DGE) matrices for each sample. DGEs were further processed in R (v. 4.4.1) using Seurat (v. 5.1.0)^101^. Filtered DGEs were imported using the function “Read10x” and only genes detected in at least 5 cells were kept for downstream analyses. Similarly, cells will less than 200 genes and 3,000 unique transcripts or more than 0.5 % mitochondrial transcripts were discarded. After merging all samples in a single Seurat object, we performed gene expression normalization and scaling for each sample independently using SCTransform^102^. We performed unbiased clustering and dimensionality reduction using the first 20 principal components on all samples together. Clusters were manually annotated in 7 cell types analyzing their marker genes **(Table S5)**. To identify differential genes induced by *MT-ATP6* mutations and sildenafil treatment in robust and reproducible way, we performed a pseudo-bulk analysis using the DESeq2 package^10^2 (v. 7.5.1), as described previously^10^3. Pseudo-bulk analysis was used to determine differentially expressed genes within the individual populations (Table S6). For each population, we generated a pseudo-bulk expression profile for each organoid by summing the expression of 250 randomly selected cells. We then filtered genes with less than 50 counts in at least 2 samples, normalized expression values by library size, estimated negative binomial dispersions. We performed different comparisons: i) LS + DMSO vs controls + DMSO (disease signature), ii) LS + sildenafil vs LS + DMSO (sildenafil signature), iii) controls + sildenafil vs controls + DMSO (negative control test). For all comparisons, we identified differentially expressed genes and adjusted P value was calculated using Bonferroni correction for multiple testing correction. To estimate glycolytic pathway activity in single cells, we leveraged the AUCell package (v. 1.26.0)^10^4 and the ‘Glycolysis’ gene set in the ‘Hallmark’ database accessed from the MsigDB package (v 7.5.1)^10^5. Plots were generated with ggplot2 (v. 3.5.1) and piping using dplyr (v. 1.1.4).

### Proteomics analysis

We carried out proteomics for four MT-ATP6 mutant NPC lines (ATP6_2, ATP6_4, ATP6_5, and ATP6_7) and four healthy control NPC lines (CTRL_1, CTRL_2, CTRL_3, and CTRL_4) with n=3 biological replicates per condition that were treated with either DMSO or 10 *μ*M sildenafil in DMSO for 16 h. Cells were washed 3x with ice-cold PBS, detached using a cell scraper, transferred to pre-chilled 1.5 ml tubes and centrifuged at 1000-3000 rpm for 10 min at 4 °C. Subsequently, the supernatant was removed, and intact cells were snap-frozen. Cells were lysed under denaturing conditions in 300 *μ*l of a buffer containing 3 M guanidinium chloride (GdmCl), 10 mM Tris(2-carboxyethyl)phosphine (TCEP), 40 mM chloroacetamide, and 100 mM Tris-HCl pH 8.5. Lysates were denatured at 95 °C for 10 min shaking at 1000 rpm in a thermal shaker and sonicated in a water bath for 10 min. The protein concentration of each sample was measured with a BCA protein assay kit (23252, Thermo Scientific). 500 ng protein was used per sample and diluted with a dilution buffer containing 10 % acetonitrile and 25 mM Tris-HCl, pH 8.0, to reach a 1 M GdmCl concentration. Then, proteins were digested with LysC (Roche; enzyme to protein ratio 1:50, MS-grade) shaking at 800 rpm at 37 °C for 3.5 hours. The digestion mixture was diluted again with the same dilution buffer to reach 0.5 M GdmCl, followed by tryptic digestion (Roche, enzyme to protein ratio 1:50, MS-grade) and incubation at 37 °C overnight in a thermal shaker at 800 rpm. Peptides were acidified with formic acid to a final concentration of 2 %. LC-MS/MS was performed by nanoflow reversed-phase liquid chromatography (Dionex Ultimate 3000, Thermo Scientific) coupled online to a timsTOF SCP mass spectrometer (Bruker Daltonics) using the data-independent acquisition (DIA) method with parallel accumulation serial fragmentation (PASEF). Briefly, LC separation was performed using the Aurora Ultimate column (25 cm x 75 *μ*m ID, C18, 1.7 *μ*m beads, IonOpticks, Victoria, Australia). 150 ng of each desalted digest was applied to the column, peptides were eluted using a gradient of 3.8 to 38 % solvent B in solvent A over 60 min (total run time) at a flow rate of 400 nL per minute. Solvent A was 0.1 % formic acid and solvent B was 79.9 % acetonitrile, 20 % H2O, and 0.1% formic acid. MS data were processed with Dia-NN (v1.8.1) and searched against an in silico predicted human spectral library. The “match between run” feature was used and the mass search range was set to m/z 400 to 1000.

### Targeted metabolomics profiling

We carried out metabolomics in four MT-ATP6 mutant NPC lines (ATP6_2, ATP6_4, ATP6_5 and ATP6_7) and four healthy control NPC lines (CTRL_1, CTRL_2, CTRL_3 and CTRL_4) using n=4 replicates per condition that were treated with either 0.1 % DMSO or 10 *μ*M sildenafil in 0.1 % DMSO for 16 h. NPCs were washed 1 x with PBS, detached in 600 *μ*l ice-cold extraction buffer (methanol, acetonitrile and Milli-Q water at a ratio of 40:40:20) using a cell scraper, transferred into an ice-cold 1.5 ml tube, and stored at −80 °C until use. Metabolites were extracted from cells with 500*μ*l of cold extraction solvent (Acetonitrile:Methanol:MQ; 40:40:20, Thermo Fischer Scientific). Subsequently, samples were processed with three cycles of sonication (60 s) and vortexing (120 s) followed by centrifugation at 14,000 rpm at 4 °C for 5 min. Next, the samples were centrifuged, the supernatants transferred to evaporation tube and evaporated to dry under nitrogen stream. Samples were reconstituted in 40 *μ*l extraction buffer (40:40:20; acetonitrile:methanol:MilliQ) and transferred to LC-MS vials. 2 *μ*l of the samples were analyzed with Thermo Vanquish UHPLC coupled with Q-Exactive Orbitrap mass spectrometer equipped with a heated electrospray ionization (H-ESI) source probe (Thermo Fischer Scientific). A SeQuant ZIC-pHILIC (2.1 × 100 mm, 5 μm particle) column (Merck) was used for chromatographic separation. The gradient elution was carried out with a flow rate of 0.100 ml/min and mobile phase gradient with 20 mM ammonium hydrogen carbonate, adjusted to pH 9.4 with ammonium solution (25 %) as mobile phase A and acetonitrile as mobile phase B, 0-2 min 80 % B, 2-17 min 80-20 % B, 17-24 min 80 % B. The column oven and auto-sampler temperatures were set to 40 ± 3 °C and 5 ± 3 °C, respectively. Following setting were used for MS: full scan range: 55-825 m/z, polarity switching; resolution of 35,000, the spray voltages: 4250 V for positive and 3250 V for negative mode; the sheath gas: 25 arbitrary units (AU); the auxiliary gas: 15 AU; sweep gas flow 0; capillary temperature: 275 °C; S-lens RF level: 50.0. Instrument control was operated with the Xcalibur software (Thermo Fischer Scientific). The metabolite annotation and integration were done with the TraceFinder 5.1 software (Thermo Fischer Scientific) using confirmed retention times by in-house standard library (MSMLS-1EA, Merck) and their m/z. The data quality was monitored throughout the run using pooled QC sample prepared by pooling 5 *μ*L from each suspended sample and interspersed throughout the run every 10 samples. The data was quality controlled for peak quality (poor chromatography), prefiltered with 20 % RSD cutoff of the pooled QC and noise. We identified metabolites that were differentially present within samples **(Table S3)**.

Quantitative analysis of NAD+, NADH, NADP+, NADPH, and reduced and oxidized glutathione was carried out as a service in NADMED laboratory (Helsinki, Finland) from same homogenates submitted for metabolomics analysis. NPCs were washed on the plate with PBS buffer to remove protein of the culture media followed by the addition of cold extraction solvent acetonitrile:methanol:MilliQ; 40:40:20 to quench cellular metabolism. Obtained sample homogenates were shipped on dry ice to the measurement facility. Before analysis the homogenates were equilibrated to room temperature and centrifuged at 20000 x g for 10 min at 4 °C to remove proteins. Next, NAD+, NADH, NADP+, NADPH, GSH pool, and GSSG were measured individually from every cell extract using modified cyclic enzymatic reactions with colorimetric detection. For normalization of the results, protein content was measured using the pellets obtained after centrifugation of the homogenate.

### Multi-omics integration

We performed multi-omics integration as previously described^45^. We integrated bulk transcriptomics, proteomics, and metabolomics datasets derived from LS NPCs and control NPCs (disease signature), as well as transcriptomics and metabolomics datasets from sildenafil-treated LS NPCs and untreated LS NPCs (sildenafil signature). For both cases, we used all significantly deregulated molecules (adjusted p-value ≤ 0.05) as input lists for the OmicsNet platform (v.2.0) and retrieved their interactions based on STRING (confidence score 0.7) and Recon3D databases. Molecular interactions were filtered with the Prize-Collecting Steiner Forest (PCSF) algorithm, retaining only the most informative nodes regarding network organization and functionality. The final network was analyzed for centrality measures using the CytoNCA plugin in Cytoscape (v.3.10.2) and enrichment analysis using the KEGG database (Release 112.0). For network visualization, we chose the Organic Layout from yFiles plugin in Cytoscape. To dissect the rescue mechanism of sildenafil, we searched for molecules with reversed expression in the disease signature and sildenafil signature datasets. Molecules were annotated in the REACTOME and GeneOntology databases for pathway and biological processes enrichment analyses, respectively, using the clusterProfiler package (v4.10.0).

### Assessment of mitochondrial membrane potential (MMP)

For MMP quantification, we applied a high-content analysis (HCA)-based live-cell detection assay that we previously established^29^. Briefly, we seeded NPCs onto black-wall, clear-bottom 96-well plates (SCREENSTAR, Greiner) pre-coated with Geltrex (Thermo Fisher Scientific) at a density of 1.5 × 10^5^ cells/cm^2^, and incubated them in NPC medium overnight at 37 °C, 5 % CO_2_. The next day, NPCs were treated with either DMSO or Sildenafil dilution series in 100 *μ*l and incubated for 16 h at 37°C, 5% CO_2_. On the day of the assay, we live-stained NPCs with 10 nM TMRM (Molecular Probes, Life Technologies) and 1 *μ*g/*μ*l Hoechst (33342, Thermo Fisher Scientific) in NPC medium for 30 min at 37 °C and 5 % CO_2_. After 30 min incubation, cells were washed with 100 *μ*l NPC medium without phenol red. NPCs were kept in 100 *μ*l medium (w/o phenol red) for the duration of the assay. Imaging was performed with an Operetta CLS high content imaging system (Revvity) with 20 x objective in confocal mode. Imaging time did not exceed 30 min. TMRM and Hoechst 33342 intensity within the cells was calculated using Columbus software (Revvity, version 2.9.0).

For MMP assessment using cytofluorimetry, we used LSR-Fortessa X-20 (Becton Dickinson) to monitor the fluorescence of TMRM (Invitrogen). Cells were harvested and resuspended in culture medium at approximately 1×10^6^ cells/ ml. The uncoupling agent carbonyl cyanide 3-chlorophenylhydrazone (CCCP, 50 μM final concentration) was added to the sample and incubated for 5 min at 37°C, 5% CO_2_ to depolarize mitochondria. TMRM probe (20 nM final concentration) was added and incubated for 30 min at 37 °C and 5 % CO_2_. After incubation, cells were washed once in 1 ml of culture medium, then resuspended in 300 μl of D-PBS and analyzed on the flow cytometer. The TMRM-emitted fluorescence was detected by emission filters appropriate for R-phycoerythrin “PE” (585 nm ± 42 nm). An appropriate gating strategy was applied to select only live and single cells. Data were analyzed with FlowJo software (version 7.6); the median fluorescence intensity (MFI) per each sample was calculated and normalized on the relative unstained sample. TMRM intensity was quantified in at least 20.000 cells per sample; at least three biological replicates were analyzed. Statistical analysis was performed by one-way Anova. p < 0.05 was considered statistically significant.

### Compound screening

LS NPCs (ATP6_2) were seeded at a density of 1.5 × 10^5^ cells/ cm^2^ onto Geltrex-coated 384-well black-wall, clear-bottom plates (Revvity) in 30 *μ*l NPC medium using a Multidrop liquid dispenser (Thermo Fisher Scientific), which was calibrated prior to each usage. Assay plates with cells were placed into a rotating incubator (Thermo Electron, Cytomat) at 37 °C and 5 % CO_2_ for 24 h. Cells were treated with 5 *μ*M (0.05 % DMSO) of 5,632 different compounds from a drug repurposing library^35^. Compounds were transferred to 384-well assay plates using an Echo 550 (LabCyte). Columns 23 and 24 contained controls on each plate. In row 23, a combination of 5 *μ*M FCCP (Biozol) and antimycin A (Sigma/Merck) was used as positive control to achieve complete mitochondrial depolarization. Row 24 contained 0.05 % DMSO as negative control. After incubation of 16 h, cells were live-stained for 30 min at 37 °C and 5 % CO_2_ with a final concentration of 10 nM TMRM and 1 *μ*g/*μ*l Hoechst in 30 *μ*l/well. Cells were then automated washed twice with 30 *μ*l NPC medium (w/o phenol red) using the Janus MDT (Revvity). Images were directly acquired using an Operetta CLS high content imaging system with 20 x objective in confocal mode. Imaging time did not exceed 30 min. TMRM intensity within the cells and nuclei count was calculated using a customized Columbus software script and downstream data processing was performed with Activity Base (IDBS). To quantify the effects of compounds on depolarizing the hyperpolarized MMP of LS NPCs (ATP6_2), we normalized the percentage of MMP depolarization (TMRM signal) and the percentage of cell count (Hoechst signal) after treatment for 16 h with 5 *μ*M of each compound in LS NPCs with respect to LS NPCs treated for 16 h with either 5 *μ*M FCCP+AA or DMSO only. To achieve this normalization, we used the formula z = (y – min(x)) / (max(x) – min(x)) * 100, where z is the final normalized value, y is the value of interest in the dataset, min(x) is the mean value of DMSO-treated LS NPCs, and max(x) is the mean value of FCCP+AA-treated LS NPCs. We next scored the normalized values (z) based on their ability to depolarize the MMP (inhibition) and classified the compounds in mild, intermediate, and strong uncouplers.

### Bioenergetic assessment

For biochemical activity of Complex V, approximately 1 × 10^7^ NPCs were suspended in 0.5 ml of 10 mM ice-cold hypotonic Tris buffer (pH 7.6) and homogenized with glass/glass Dounce homogenizer with a tight pestle by 10 strokes. 200 *μ*l of 1.5 M sucrose was added and then samples were centrifuged at 600 g for 10 min at 4 °C. Supernatants were collected and centrifuge at 14,000 g for 10 min at 4 °C. Mitochondrial pellets were suspended in 0.2 ml of 10 mM ice-cold hypotonic Tris buffer (pH 7.6) and subjected to three freeze-thaw cycles. Protein concentration was quantified with the Bicinchoninic Acid (BCA) protein assay Kit (Thermo Fisher Scientific). Complex V activity (ATP hydrolysis) was measured at 340 nm cleared from oligomycin-insensitive ATPase activities as described^106^ and normalized to citrate synthase activity^107^.

For ATP content analysis, NPCs were grown in NPC medium and then switched to DMEM glucose-free (Gibco) and Neurobasal A-Medium glucose-free (Gibco) 1:1 medium supplemented with 1 mM sodium pyruvate, 1 % B27, 0.5 % N2, 1 % Pen/Strep, 1 % L-glutamine and 0.01 % MycoZap Plus-CL with 5 mM galactose as carbon source to measure OXPHOS-derived ATP-content for 24 h. Luminescence-based ATP levels were detected in cellular suspension (5 × 10^3^ cells) with ViaLightTM Plus Kit (Lonza). ATP levels were determined by luminometry using Victor Multiple Plate Reader Spectrophotometer (Revvity). For normalization purposes, we employed the DNA quantification with CyQUANT Cell Proliferation Assay (Invitrogen). Results were then expressed as luminescence AU relative to control lines cultured in galactose.

### Neurite outgrowth quantification

We quantified neuronal outgrowth capacity following our HCA-based protocol^56^. Briefly, neurons were split with 500 μl of Accutase at 37 °C for 10-15 min and were centrifuged for 5 min at 120 x g. All pipetting steps to dissociate the cells were performed slowly and gently to minimize damage to the more sensitive neurons. 10 μM ROCK inhibitor was added to the final media after splitting to promote neuron survival. Experiments and quantification were performed as previously described^56^. Neurons generated for neurite outgrowth quantification were treated with 10 *μ*M sildenafil in 0.1 % DMSO or 0.1 % DMSO alone following media changes on days 8, 10, and 12. On day 14, the neurons were split and seeded onto black-wall, flat-clear-bottom 96-well plates (Greiner) at a density of 5,000 cells/well, and treated once more with Sildenafil or DMSO. Neurons were then cultured until day 16, when they were fixed with PFA, blocked, and stained for TUJ1 and Hoechst. Neurite images were then acquired using the Operetta CLS high content imaging system (Revvity) the TUJ1 and Hoechst signals at 20 x. Following acquisition, the neurite images were analyzed using CellProfiler (version 4.2.5). Briefly, the Hoechst signal was used to identify the nucleus as a primary object and the TUJ1 signal was used to identify the neurites as secondary objects. The nuclear objects were then expanded to serve as a proxy for the cell body and subtracted from the TUJ1 signal to leaving only the individual neurites. The results were exported as CSV for data analysis.

For *PRKG1* knock-down (KD) in dopaminergic-enriched neuronal cultures, we used small interfering RNA (siRNA) against *PRKG1* using jetPRIME transfection (Polyplus, Cat#101000027) following the manufacturer’s instruction. On day 12, neurons were transfected with 10 nM of *PRKG1* siRNA (Cat#4392420, Ambion) or scramble siRNA (Cat#4390843, Ambion) by adding the transfection mix directly to the well. On day 14, after re-seeding the neurons onto the 96-well plates, neurons were transfected again with 10 nM of *PRKG 1* siRNA or scramble siRNA. On day 16, *PRKG 1* knock-down efficiency was evaluated by qPCR and neurite length was quantified with our HCA protocol^56^.

### Calcium imaging in cortical brain organoid slices (cBOS)

cBOS were generated as previously described^54,10^8. Briefly, 3 % low melting point agarose (Gibco) was diluted in Hank’s Balanced Salt Solution (HBSS, Sigma), heated to 90 °C and slowly cooled down to 40 °C. Organoids were washed in HBSS, embedded in the agarose and cooled on ice until the agarose was solid. For slicing, a Vibratome Microm HM 650 V (Thermo Fisher Scientific) was adjusted to the following settings: frequency of 60 Hz, amplitude of 1.0 mm, velocity of 33 mm/s, and slice thickness of 300 μm. The slices were directly transferred to a petri dish filled with HBSS, placed in a cell culture hood and washed four times. Finally, the slices were collected on Millicell-CM inserts (Millipore) and placed in a 6-well plate. 750 μl of cortical differentiation medium IV (CDMIV) were added in each well, so that the membrane was soaked but the top side of the slices was still exposed to air. cBOS were cultured at 36 °C and 5 % CO_2_ for around 30-59 days before starting calcium imaging experiments.

For calcium imaging experiments, membrane permeable Oregon Green 488 BAPTA-1 AM (OGB-1, Invitrogen) was first solved in 20 % pluronic and 80 % DMSO and diluted to 200 μM in HEPES-buffered saline, which was then injected into cBOS and incubated for 30 minutes. During the incubation and throughout the experiments, cBOS were perfused with artificial cerebrospinal fluid (ACSF), containing 138 mM NaCl, 2.5 mM KCl, 2 mM CaCl_2_, 1 mM MgCl_2_, 1.25 mM NaH2PO4, 18 mM NaHCO3, and 10 mM glucose. The ACSF was bubbled with 95 % O_2_ and 5 % CO_2_, resulting in a pH of 7.4. OGB-1 was excited at 488 nm and signals were detected using the imaging software NIS-Elements and a variable scan digital imaging system (Nikon) attached to an upright microscope (Eclipse FN-1, Nikon). The microscope was equipped with a Fluor 40x/ 0.8 DIC M/N2 ∞ /0 WD 2.0 water immersion objective (Nikon) and an orca FLASH 4.0 LT camera (Hamamatsu Photonics). Images were routinely obtained at 1 Hz and emission was collected at >500 nm. Regions of interest (ROIs) representing cell somata were identified, and their signals were background-corrected. Additionally, the fluorescence signals were corrected for bleaching and analysed with OriginPro Software (OriginLab Corporation). To probe for the cellular response to acute metabolic stress, ACSF was switched to glucose-free ACSF containing 5 mM sodium azide (Honeywell) and 2 mM 2-Deoxy-D-glucose (Apollo Scientific) for 2 min. Then, it was switched back to ACSF for 20 to 30 min until cellular calcium levels recovered to their original baseline. Analysis of calcium signals was performed as described before^54^.

## Quantification and statistical analysis

We analyzed the data using GraphPad-Prism software (Prism 10, GraphPad Software, Inc.) and employed the R environment for statistical computing. For all datasets, we tested the normality of the distribution using GraphPad-Prism (D’Agostino-Pearson test for large sample sizes and Shapiro-Wilk test for sample sizes under 50). After having performed outlier test analyses, we assessed statistical significance using parametric tests for normally-distributed data (Student’s t-test, ANOVA) and non-parametric tests when normal distribution could not be verified (Mann-Whitney U and Kruskal-Wallis test). When possible, data are presented as scatter plots with individual data points showing all individual measurements. Unless otherwise indicated, we expressed the data as mean and standard deviation (mean ± SD). Experiments were repeated in independent biological experiments. Information on statistical details and number of replicates can be found in the respective figure legends.

