## Supplementary information for "Pluripotent stem cell-based drug discovery uncovers sildenafil as a treatment for mitochondrial disease"

^*^Equal first author

^§^Equal corresponding author

**Supplementary methods**

**Quantification of sildenafil concentration in patient plasma**

EDTA blood for determination of the sildenafil plasma concentration was drawn in the morning before the first dose and one hour after oral drug application. The EDTA-blood sample was centrifuged at 3000 x *g* for 10 min at 4 °C and the plasma was aliquoted and frozen at -80 °C until measurement. For mass spectrometric determination of the plasma sildenafil concentration, 100 µl sample was mixed with 400 µl of the internal standard d_3_-sildenafil (10 nM in acetonitrile) (CDN Isotopes). Proteins were precipitated by vortexing for 1 min and subsequent storage for 60 min at -20 °C. Supernatants were obtained by centrifugation at 16,000 x *g* at 4 °C for 10 min and subjected to LC-MS/MS sildenafil quantification applying the multiple reaction monitoring (MRM) approach. Chromatographic separation was achieved on a 1290 Infinity II HPLC (Agilent Technologies) equipped with a Poroshell 120 EC-C18 column (3.0 x 150 mm, 2.7 μm; Agilent Technologies) guarded by a pre-column (3.0 x 5 mm, 2.7 μm) of identical material. Water (eluent A) and acetonitrile (eluent B), both acidified with 0.1 % formic acid, were pumped with 0.4 ml/min. Elution of sildenafil and its internal standard was achieved with a 10-min linear gradient from 5 % to 90 % eluent B. Total run-time was 17 min including re-equilibration of the LC system. MS/MS analyses were carried out using an Ultivo triple-quadrupole mass spectrometer (Agilent Technologies) operating in the positive electrospray ionization mode (ESI+). The following ion source parameters were set: sheath gas temperature, 400 °C; sheath gas flow, 12 l/min of nitrogen; nebulizer pressure, 20 psi; drying gas temperature, 100 °C; drying gas flow, 7 l/min of nitrogen; capillary voltage, 3.0 kV; nozzle voltage, 0 kV. The following mass transitions were recorded (fragmentor voltage [FV] and collision energies [CE] in parentheses): sildenafil: *m/z* 475.2 → 58.0 (FV: 220 V, CE: 68 eV, *quantifier*), *m/z* 475.2 → 100.0 (FV: 220 V, CE: 28 eV), *m/z* 475.2 → 283.1 (FV: 220 V, CE: 44 eV); d_3_-sildenafil: *m/z* 478.2 → 60.9 (FV: 220 V, CE: 68 eV, *quantifier*), *m/z* 478.2 → 103.0 (FV: 220 V, CE: 28 eV), *m/z* 478.2 → 283.1 (FV: 220 V, CE: 44 eV). Peak areas were determined with MassHunter Software (Agilent Technologies) and sildenafil was directly quantified *via* its internal standard d_3_-sildenafil that was concentrated to 8 nM in the samples.

**Genotyping and heteroplasmy quantification**

Heteroplasmy levels and mtDNA haplotypes in the whole exome sequencing (WES) datasets were determined using the software MToolBox v.1^1,2^ Results were visualized in the IGV viewer v2.16^3^ **(Figure S10)**. Heteroplasmy levels of the *MT-ATP6* mutations in fibroblasts, iPSCs, and NPCs were determined using polymerase chain reaction-restriction fragment length polymorphism (PCR-RFLP) analysis, as described before.^4^ Genomic DNA was isolated using the Nucleo-Spin Tissue kit (Macherey-Nagel). We used restriction enzyme StuI (NEB, R0187, 10,000 units/ml) for the m.9185T>C mutation (wild-type: 24+90 pb, mutation: 114 pb), HpaII (NEB, R0171, 10,000 units/ml) for m.8993T>C and m.8993T>G (wild type: 25+155 bp; mutant: 180 bp), XbaI (NEB, R0145, 10,000 units/ml) for m.9176 T>G (wild type: 24+155 bp, mutant: 179 bp). The percentage of cleaved versus uncleaved fragments was determined by capillary electrophoresis and laser detection of the FAM-labelled RFLP-fragments using the 3,500 Series Genetic Analyzer (Applied Biosystems, RRID:SCR_021901) and normalized to a standard curve of known degrees of heteroplasmy.

**Cranial magnetic resonance imaging (cMRI)**

cMRI was performed for the six patients with LS carrying *MT-ATP6*. For Patient 1, imaging showed areas of increased T_2_-signal intensity in the *Putamen* and *Nucleus caudatus* on both sides **(Figure 5g)** and at the perisylvian gray matter **(Figure S9h)**. For patient 2, imaging showed lesional areas in the *Putamen* and *Nucleus caudatus* in both T_2_-weighted images **(Figure 5g)** and Fluid Attenuated Inversion Recovery (FLAIR) images **(Figure S9h)**. For patient 3, imaging was carried out in early childhood **(Figure 5g)** and adolescence **(Figure S9h)**. Areas with increased T_2_-signal intensity in early childhood (left) had spontaneously resolved in adolescence. In patient 4, FLAIR imaging showed areas of increased signal intensity at the *Nuclei caudati* on both sides **(Figure 5g)** and enlargement of the internal and external liquor spaces due to diffused brain atrophy **(Figure S9h)**. For patient 5, imaging showed T_2_-weighted images delayed myelination, enlarged external cerebral spinal fluid spaces, and a general frontal brain atrophy **(Figure 5g)**. The T_2_-signal intensity in the brain stem of patient 5 affected the *Fasciculus longitudinalis* and the *Substantia nigra* **(Figure S9h)**. For patient 6, imaging showed areas of increased T_2_-signal intensity in the *Putamen, Pallidum*, and *Nucleus caudatus* on both sides **(Figure 5g)** and in the cortical gray matter **(Figure S9h)**.

**Toxicity assays**

Cell Titer-Glo assay (Promega) was used to determine viability based on cellular ATP concentration. NPCs were isolated using Accutase and seeded onto Geltrex-coated 96-well plates at a density of 1.5 x 10^5^ cells/cm^2^ and incubated in NPC medium overnight at 37 °C and 5 % CO_2_. The day after, NPCs were treated with DMSO, FCCP+AA or sildenafil dilution series in 100 µl and incubated for 16 h at 37 °C and 5 % CO_2_. One column contained an untreated reference. 50 µl Cell Titer-Glo reagent solution was added to each well and placed on a plate shaker (1 min, 100 rpm) and incubated at ambient temperature for 10 min to ensure cell lysis. 100 µl of the solution was transferred onto a new white wall, white bottom 96-well plate, and luminescence was measured using an EnSight multimode plate reader (Revvity).

For proliferation, NPCs were seeded in a coated 96-well plate (30,000 cells/well) and incubated at 37 °C and 5 % CO_2_. Medium glucose concentration was either 21 mM or 4.5 mM, or glucose was substituted with galactose. With the first medium change, the medium was supplemented with 1 µM sildenafil, 10 µM sildenafil or DMSO (vehicle), respectively, or left without treatment. For toxicity analysis, the medium was additionally supplemented with Incuyte dye red (Sartorius, REF: 4717) to indicate dead cells. Supplemented medium was changed every other day. Over the course of the experiment, the well plates were placed in the Cellcyte X (Cytena) microscope with a 10 x objective. Four pictures of each well were taken every three hours. Pipeline setup and analysis were done with the software CELLCYTE Studio (Cytena). Channels were “enhanced contour” for confluency and “red channel” (600 ms, 9 dB) for toxicity. Autofocus was set to “Classifier-based”. Pictures taken with the “enhanced contour” channel were analyzed with the software-specific protocol “Cell Confluence” with the following settings: 58 a.u. contrast sensitivity, 2 a.u. smoothing, 200 µm^2^ filled hole size, 100 µm^2^ min. object size. Pictures taken with “red channel” were analyzed with the software-specific protocol “object count” with the following settings: 30 a.u. contrast sensitivity, 50 a.u. separation sensitivity, 0 µm^2^ filled hole size, 10 µm^2^ min object size. Analysis results were exported to Excel and further processed with R studio.

**Analysis of mitochondrial nanostructure**

To examine the impact of sildenafil on the mitochondrial nanostructure, we immunolabeled LS NPCs (ATP6_2) and control NPCs (CTRL_1) for MIC60, a core protein of the mitochondrial contact site and cristae organizing system (MICOS). For immunolabelling of MIC60, cells were fixed with prewarmed (37 °C) 4 % formaldehyde in PBS (137 mM NaCl, 2.68 mM KCl and 10 mM Na2HPO4, pH 7.4) for 5 min at RT. Fixed cells were extracted with 0.5 % (v/v) Triton X-100 in PBS, blocked with 5 % (w/v) BSA in PBS. Afterwards, cells were incubated with diluted primary antibodies against MIC60 (Abcam, Cambridge, UK) in 5 % (w/v) BSA in PBS for 1 h at RT. After washing with PBS, the primary antibodies were detected with secondary goat anti-rabbit antibodies labelled with Abberior STAR RED (Abberior, Göttingen, Germany)- After washing with PBS, the cells were mounted in Mowiol with 0.1 % 1,4-Diazabicyclo[2.2.2]octan (DABCO). Confocal and stimulated emission depletion (STED) microscopy were performed using an INFINITY platform (Abberior Instruments, Göttingen, Germany). The objective was an UPLXAPO60xO, NA: 1.42, oil objective (Olympus, Tokyo, Japan). For STED super-resolution microscopy, Abberior STAR RED was excited at 640 nm and depleted with 775 nm. The fluorescence was collected between 650 nm and 720 nm. Images were recorded with a pixel size of 20 nm and a dwell time of 30 µs per pixel.

To detect structural differences between control and LS NPCs, we captured approximately 4000 STED images and trained a neural network classifier on multiple biological replicates to distinguish control NPCs (P[healthy]-score = 1) from LS NPCs (P[healthy]-score = 0). A deep residual neuronal network (ResNet34) was trained with STED data of DMSO-treated LS NPCs (ATP6_2) and control NPCs (CTRL_1) in a supervised setting to distinguish between both conditions. For the training, STED images were percentile normalized and divided into 512 x 512 pixel-sized regions (patches). Every patch was assigned to the class “healthy” when it originated from control NPCs and to the class “ill” when it originated from LS NPCs. Using a mean intensity threshold, patches without a sufficient fluorescence signal were allocated to a third class (“low information”). The final training dataset consisted of equal numbers of patches from three different biological replicates and included in total 75,000 patches of the “healthy, “ill” and “low information” classes in a 0.4:0.4:0.2 ratio. The performance throughout the trainings process was monitored with a validation set of 11,000 patches. For every patch, the model predicts logits values corresponding to the three classes, that are then transformed with softmax function to receive a per class probability. The per-class accuracy computed from a confusion matrix was used as mean validation metric after each epoch and as an indicator for the best checkpoint. The final evaluation of the neuronal network after the training was carried out with a test set of 140,000 patches with a 0.4:0.4:0.2 ratio. The trained model was used to assign a P[healthy] score from zero to one to single STED images. For each STED image with a size of 2048 x 2048 pixels, 49 partially overlapping patches of 512 x 512 pixels were extracted and evaluated with the model. Patches with insufficient fluorescence signal were assigned to the “low information” class and excluded from the evaluation. For the remaining patches a P[healthy]-score was predicted. The P[healthy]-score of the single patches were averaged to assign a P(healthy)-scores to each STED image of 2048 x 2048 pixels. STED images which had a proportion of “low information” patches above 90 % were excluded. After training, the classifier could reliably differentiate the two conditions on images that were not used for the training, indicating alterations of the MIC60 fluorescence signal in LS NPCs.

To gain further insights, we reviewed a subset of images individually. Confocal microscopy revealed a tubular mitochondrial network in both NPC cultures, with no evident phenotypic alterations in LS NPCs. STED imaging showed a largely uniform distribution of MIC60 clusters along mitochondrial tubules in control NPCs, whereas in LS NPCs, MIC60 appeared to accumulate at the sides of mitochondrial tubules. To investigate this phenotype, we manually analyzed the MIC60 pattern across the mitochondrial tubules in approximately 300 images. A semi-automated MATLAB-based analysis was employed to assess the MIC60 distribution in LS NPCs (ATP6_2) and control NPCs (CTRL_1). After blinding, individual elongated mitochondria were manually analyzed. The mitochondrial centerline was determined by marking discrete points along the mid-axis, which were then fitted to a Bézier curve to approximate the mitochondrial central axis. The fluorescence intensity profile across the mitochondrial transverse axis was measured every 10 nm at a length of 600 nm with its center at the Bézier curve. The profiles were averaged for all selected isolated mitochondrial fragments in a single STED image. A homogeneous MIC60 distribution results in a fluorescence intensity profile with a broad plateau at the central maximum along the transversal mitochondrial axis. MIC60 accumulation at the outer edge of the mitochondria creates a central intensity dip. LS NPCs revealed a clear dip in the distribution of fluorescence intensity. The altered distribution of MIC60 was expressed as a center-to-maximum fluorescence intensity ratio across the transversal mitochondrial axis. In LS mitochondria this ratio was reduced compared to control samples with sildenafil treatment of LS NPCs restoring the fluorescence intensity ratio to healthy levels.

**Animal experiments**

Germline *Ndufs4* KO mice were obtained from the University of Washington.^5^ All mice were on the C57/BL6/J background and were fed the regular diet from Harlan Teklad.^5^ All animal experiments were approved by the Institutional Animal Care and Use Committee (IACUC). Sildenafil citrate (Sigma) was dissolved in DMSO, then diluted with tap water to obtain the final concentration of 400 mg/L drinking water and given *ad libitum*.

Electrocardiography (ECG) recordings were recorded from conscious mice, using the INDUS Rodent Surgical Monitoring system without anesthesia. Transthoracic echocardiograms to measure ventricular size, wall thickness, and ejection fraction were performed on mice using the Vevo 2700 VisualSonics System (Toronto, ON, Canada). Intraperitoneal injection with 0.1 mg midazolam was applied for anxiolytic muscle relaxant effects, without any anesthetic agent that may cause cardiac suppression. Standard 2D, M-mode, Doppler, Tissue Doppler images were obtained using a 30-MHz linear array transducer, including the parasternal short and long axis. All image analysis was performed by Vevo 2100 analysis software (v.1.5) after rapid image collection. The diastolic function was measured using tissue Doppler imaging of the mitral annulus (E`) and conventional mitral inflow (E wave).

Metabolic chamber by Promethion (Sable Systems International) was applied to characterize the metabolism of whole-animal energy expenditure.^6^ The 16-cage metabolic chamber system directly measures various parameters over a 72‐hour period, such as heat production, oxygen consumption (VO_2_), CO_2_ production, and XY activity movement. The respiratory exchange ratio (RER) is calculated as the ratio of carbon dioxide production (VCO_2_) to oxygen consumption. This parameter is useful to estimate the metabolic status. The xy axis detection of animal motion measure activity during day-time and night-time*.* The formula applied was Heat = 1.232*VCO_2_+3.815*VO_2_.

**Immunostaining**

We fixed NPCs and neurons grown on Matrigel-coated coverslips with 4 % PFA (Thermo Fisher Scientific) in PBS for 20 min at RT and washed two times with PBS. For permeabilization, we incubated the fixed cells with a blocking solution containing 10 % normal donkey serum (DNS) (Merck Millipore) and 1% Triton X-100 (Sigma-Aldrich) in PBS with 0.05 % Tween 20 (Sigma-Aldrich, St. Louis, MO, USA) (PBS-T) for 1 h at RT. We diluted primary antibodies in blocking solution and incubated them overnight at 4 ^o^C on a shaker. Next, the primary antibody was removed, and the wells were rinsed three times with PBS. Corresponding secondary antibodies (all Alexa Fluor, 1:2000, Thermo Fisher Scientific) together with 1:2500 Hoechst 33342 (Thermo Fisher Scientific) were diluted in blocking solution and added to the wells for 1 h at RT on a shaker. Finally, the staining solution was removed, and the wells were rinsed with PBS three times following mounting of the coverslips on microscopic slides. Details on the primary and secondary antibodies used are reported in **Resources Table**. We acquired the images of 2D cultures using the fluorescence microscope ZEISS Axio Observer Apotome 3 (Zeiss) in combination with the ZEN Microscopy Software (Zeiss) and further processed with ImageJ.

For staining cortical brain organoids, we used 4 % PFA (Thermo Fisher Scientific) in PBS for 1 h at RT and sliced them using the Vibratome Microm HM 650 V (Thermo Fisher Scientific). The samples were placed in a 3 % LB agar (Sigma) solution in PBS and put in a cool place until the agar was solidified. The agar shape was freed from its mould and attached on the carrier plate of the vibratome and placed in cold PBS. The cutting procedure was performed at an amplitude of 1.0 mm, a frequency of 60 Hz and a velocity of 13 mm/s. The organoids were sliced with a thickness of 100 μm and gently transferred to SuperFrost Plus glass slides (VWR) for staining. Blocking was carried out for 1 h at RT with a blocking solution containing 1 x PBS, 10 % donkey serum (Sigma Aldrich), 0.1 % Tween-20 (Sigma), and 1 % Triton-X (Merck). Primary antibodies were dissolved in blocking solution and applied on the slides and incubated at 4 °C overnight. The next day, the slides were rinsed three times for 10 min in 1 x PBS and then exposed to secondary antibodies in blocking solution at a dilution of 1:300 counterstained with 1:2500 Hoechst 3342 (Invitrogen). The slides were incubated for 1 h at RT protected from light and subsequently rinsed three times for 10 min with 1 x PBS. Finally, coverslips were mounted using Pro-Long Glass Antifade Mountant (Invitrogen) on microscopic slides. Details on the primary and secondary antibodies used are reported in **Resources Table**. Images of brain organoids were acquired using the Eclipse 90i upright widefield microscope (Nikon Microscope solutions) equipped with the imaging software NIS-Elements Advanced Research 3.2 (Nikon). Large images (7 x 7 stitches) were taken with a dry 20 x objective (Plan Apo VC 20 x / 0.75 air DIC N2 ∞/0.17 WD 1.0, Nikon Microscope Solutions) and three different filter channels (DAPI, FITC, TRITC). Specific structures of interest within the slices were imaged with the Confocal laser scanning microscope C1 (Nikon Microscope Solutions) and a dry 20 x objective (Plan Apo VC 20 x / 0.75 air DIC N2 ∞/0.17 WD 1.0, Nikon Microscope Solutions). Images were taken using the imaging software EZ-C1 Silver Version 3.91 at z-stack settings (1.1 μm step size, 10-30 steps).

**Quantitative polymerase chain reaction (qPCR)**

Isolated total RNA was transcribed into complementary DNA (cDNA). Into a nuclease-free microcentrifuge tube, 1 μl of oligo d(T)_18_ primers (Thermo Fisher Scientific), 1 μl of 10 mM dNTP mix (Thermo Fisher Scientific) and the respective volume of RNA solution that contained 1 μg RNA were filled up to 12 μl with RNase-free, distilled water. In the Mastercycler X50s (Eppendorf), the mixture was heated to 65 °C for 5 min and then quickly chilled on ice. 4 μl of 5 x First-Strand Buffer, 2 μl of 0.1M DTT and 1 μl of RNaseOUT (Invitrogen) were added to the tube, gently mixed and incubated at 37 °C for 2 min. 1 μl (200 units) of Moloney Murine Leukemia Virus Reverse Transcriptase (M-MLV RT) was added to the reaction tube and mixed. The reaction mixture was incubated for 50 min at 37 °C, and at last inactivated by heating at 70 °C for 15 min. For the qPCR reaction, the synthesized cDNA was mixed with 10 µM primer mix (primers designed with Integrated DNA technologies, see Tab. X) and SYBR^TM^ Green PCR Master Mix (Applied Biosystems) diluted in water in a 96-well plate. The plate was transferred to CFX96™ Real-Time System qPCR machine (Bio-Rad) and the following program was started using CFX96 software: 2 min at 50 °C, 10 min at 95 °C, 39 x (15 s at 95 °C, 30 s at 62 °C, 30 s at 72 °C), 15 min at 95 °C, 5 s at 62 °C, and 50 s at 95 °C. To check the quality of the qPCR products, melting curves were assessed first. The obtained CT values were analyzed using the 2^-ΔΔCT^ method. For this, the CT values of the genes of interest were subtracted from the CT values of the housekeeping genes *GAPDH* and *OAZ1*. Primer sequences are reported in **Resources Table**.

**Immunoblot analysis**

Pooled cells (at least 1.5 x 10^6^ NPCs) were homogenized in RIPA buffer (150 mM NaCl, 5 mM EDTA pH 8, 50 mM Tris pH 8, 1 % NP- 40, 0.5 % sodium deoxycholate (DOC), 0.1 % sodium dodecyl sulfate (SDS)) in the presence of protease and phosphatase inhibitors. Samples were sonicated in an ultrasonic bath (Branson 1800-E) for 1 min, incubated in ice for 30 min and then centrifuged at 10,000 x g for 15 min at 4 °C. Protein concentration was determined with the Bicinchoninic Acid (BCA) protein assay kit (Thermo Scientific). 30 μg of proteins were run through a 4 % separating /10 % resolving polyacrylamide gel with stain-free technology (Bio-Rad laboratories), to allow detection and quantification. Proteins were then electroblotted onto a PVDF membrane. After 1 h of blocking with 3 % BSA in TBS-Tween at room temperature (RT), the membranes were incubated with the primary antibodies overnight at 4 °C and subsequently probed with HRP-conjugated secondary antibodies for 2 h at RT. Chemiluminescence-based immunostaining (SuperSignal West Pico PLUS Chemiluminescent Substrate, Thermo Scientific) was performed. Images were acquired with the ChemiDoc MP Imaging system. All primary and secondary antibodies used are reported in **Resources Table**. Data analysis and quantification was performed using Image Lab software, 6.1 (Bio-Rad Laboratories).

**Computational modeling based on calcium signaling**

To determine the effect of impaired calcium signaling on mitochondrial function in LS, we converted the calcium fluorescence traces from cBOS **(Figure 4b)** into corresponding cytosolic calcium concentration (${[{Ca}^{2+}]}_{c}$) using the formula developed before^7^. That is,

$\Delta{[{Ca}^{2+}]}_{c}= k_{D}\left( \frac{F_{max}}{F_{0}}\left( 1-\frac{1}{R_{F}} \right)\frac{\Delta F}{{(\Delta F}_{max}-\Delta F){\Delta F}_{max}} \right)$. (1)

Where ${\Delta[{Ca}^{2+}]}_{c}= {[{Ca}^{2+}]}_{c}- {[{Ca}^{2+}]}_{0}$ is the change in ${[{Ca}^{2+}]}_{c}$, ${[{Ca}^{2+}]}_{0}$ = 0.1 µM is the baseline cytosolic Ca^2+^ concentration, $k_{D}$ = 0.2 μM in the dissociation constant for OBG-1, and $R_{F}= \frac{F_{max}}{F_{min}}$. *F_max_* and *F_min_* are the maximum and minimum fluorescent values in a trace, respectively. The instantaneous change in fluorescence is $\Delta F=F-F_{0}$, where $F_{0}$ = 0.01 is the baseline fluorescence. Finally, ${\Delta F}_{max}=F_{max}-F_{0}$ with *F_max_* as the maximum fluorescence in the trace. To match the smaller timestep used in simulating mitochondrial bioenergetics, the time trace was interpolated. The resulting ${[{Ca}^{2+}]}_{c}$ values as functions of time were fused with the computational model for mitochondrial function discussed below and solved using RK4 method in MATLAB 2023b. Rate equations for mitochondrial Ca^2+^ ([Ca^2+^]_m_), NADH ([NADH]_m_), ADP ([ADP]_m_), membrane potential ([Δ𝜓]), and cytosolic ADP ([ADP]_c_) are adopted from^8,9^ and given as:

$\frac{d{[{Ca}^{2+}]}_{m}}{dt}=f_{m}(J_{MCU}- J_{NCX}+ J_{X})$,	(2)

$\frac{d{[NADH]}_{m}}{dt}= J_{PDH}+J_{AGC}- J_{o}$,	(3)

$\frac{{d[ADP]}_{m}}{dt}= J_{ANT}- J_{F1F0}$,	(4)

$\frac{{d[ADP]}_{c}}{dt}= J_{HYD}- \delta J_{ANT}$,	(5)

$\frac{d\Delta\psi}{dt}=\frac{1}{c_{mito}}(b_{1}J_{O}-b_{2}J_{F1F0}- J_{ANT}-J_{H,LEAK}-J_{NCX}-{2J}_{MCU}-{2J}_{X}-J_{AGC})$.	(6)

Where Ca^2+^ flux through mitochondrial Ca^2+^ uniporter (MCU) and Na^+^/Ca^2+^ exchangers (NCX) are given as:

$J_{MCU}= v_{MCU}\frac{\frac{{[{Ca}^{2+}]}_{c}}{K_{1}}\left( 1+ \frac{{[{Ca}^{2+}]}_{c}}{K_{1}} \right)^{3}}{\left( 1+ \frac{{[{Ca}^{2+}]}_{c}}{K_{1}} \right)^{4}+ \frac{L}{\left( 1+ \frac{{[{Ca}^{2+}]}_{c}}{K_{2}} \right)^{2.3}}}e^{p_{1}\Delta\psi}$,	(7)

$J_{NCX}= v_{NCX}\left( \frac{{[{Ca}^{2+}]}_{m}}{{[{Ca}^{2+}]}_{c}} \right)e^{p_{2}\Delta\psi}$.	(8)

The additional Ca^2+^ flux (J_x_) was added to capture the many experimental results, and particularly the observations that mitochondrial Ca^2+^ does not drop in MCU knocked-down cells.^8,9^

$J_{x}= k_{x}\left( {[{Ca}^{2+}]}_{c}- {[{Ca}^{2+}]}_{m} \right)e^{p_{3}\Delta\psi}$.	(9)

The effects of Pyruvate Dehydrogenase (PDH)-catalyzed reaction, glycolytic pathway (k_GLY_), and the TCA cycle (reduction of [NAD+]_m_ into NADH) is formulated as:

$J_{PDH}= k_{GLY}\frac{1}{q_{1}+ \frac{{[NADH]}_{m}}{{[{NAD}^{+}]}_{m}}}\frac{{[{Ca}^{2+}]}_{m}}{q_{2}+ {[{Ca}^{2+}]}_{m}}$,	(10)

The Aspartate-Glutamate Carrier (AGC) is part of MAS NADH shuttle system defined as

$J_{AGC}= v_{AGC}\frac{{[{Ca}^{2+}]}_{c}}{K_{AGC}+ {[{Ca}^{2+}]}_{c}}\frac{q_{2}}{q_{2}+{[{Ca}^{2+}]}_{m}}e^{p_{4}\Delta\psi}$.	(11)

The rate at which NADH is oxidized in the Electron Transport Chain (ETC) and the rate at which protons are extruded from the mitochondria are combined into one equation as

$J_{O}= k_{O}\frac{{[NADH]}_{m}}{q_{3}+{[NADH]}_{m}}\left( 1+ e^{\frac{\Delta\psi- q_{4}}{q_{5}}} \right)^{-1}.$	(12)

The activity of Adenine Nucleotide Translocator (ANT) in electrogenic exchange of ATP or ADP across the inner mitochondrial membrane is given by

$J_{ANT}= v_{ANT}\frac{1- \left( \frac{\propto_{c}}{\propto_{m}} \right)\left( \frac{[{ATP]}_{c}}{{[ADP]}_{c}} \right)\left( \frac{{[ADP]}_{m}}{{[ATP]}_{m}} \right)}{\left( 1+ \propto_{c}\frac{[{ATP]}_{c}}{{[ADP]}_{m}}e^{-0.5\frac{F\Delta\psi}{RT}} \right)\left( 1+ \frac{[{ADP]}_{m}}{{\propto_{m}[ATP]}_{m}} \right)}e^{-\frac{F\Delta\psi}{RT}}$, (13)

where $\propto_{c}$ and $\propto_{m}$ denote the fact that only a fraction of nucleotides has access to the transporter. The dependence of the membrane potential is due to the negatively charged ADP and ATP. The rate of ATP synthesis by F_1_F_0_-ATPase is modeled as

$J_{F1F0}= v_{F1F0}\left( \frac{q_{6}}{q_{6}+ [{ATP]}_{m}} \right)\left( 1+ e^{\frac{q_{7}- \Delta\psi}{q_{8}}} \right)^{-1}$.	(14)

J_HYD_ represents the rate of ATP consumption (ATP hydrolysis) in the cytosol. The first term encodes the link between Ca^2+^ activity and ATP consumption in the cytosol whereas the second term captures ATP-consuming processes in the cytosol.

$J_{HYD}= \frac{V_{SERCA}}{2}+ k_{HYD}\frac{[{ATP]}_{c}}{K_{h}+ [{ATP]}_{c}}$.	(15)

The activity of SERCA and its dependence on ATP is formulated as

$V_{SERCA}= v_{P}\frac{{{[Ca}^{2+}]}_{c}^{2}}{K_{P}^{2}+ {{[Ca}^{2+}]}_{c}^{2}}\frac{{[ATP]}_{c}}{K_{e}+ {[ATP]}_{c}}$.	(16)

The Ohmic mitochondrial proton leak is given as

$J_{H,LEAK}= q_{9}\Delta\psi+ q_{10}$.	(17)

Finally, the total NADH, ADP, and ATP in the mitochondria and cytosol are given by conservation equations. That is,

${[NADH]}_{m}+ {[{NAD}^{+}]}_{m}= {[NAD]}_{m}^{TOT}$,	(18)

${[ADP]}_{m}+ {[ATP]}_{m}= {[A]}_{m}^{TOT}$,	(19)

${[ADP]}_{c}+ {[ATP]}_{c}= {[A]}_{c}^{TOT}$.	(20)

Various parameters used in the model are defined and listed in **Table S7**.

**Human blood-brain barrier (BBB) model**

Control iPSCs (CTRL_8) and LS iPSCs (ATP6_2) were differentiated into brain capillary endothelial cells (BCECs) following previously described protocols.^10,11^ Single cells were isolated using Accutase (Thermo Fisher Scientific) and seeded onto Matrigel-coated 6-well plates (Nunc™, Thermo Fisher Scientific) in 2 ml/well mTeSR™ Plus supplemented with 10 µM Y-27632 (STEMCELL Technologies). The starting cell number was optimized for both cell lines respectively. After 3 days, medium was changed to 2 ml/well unconditioned medium (UM) when the optimal cell density of 2 - 4 x 10^3^ cells/cm^2^ was reached (referred to as day 0). UM was composed of 78.5% DMEM/F12 (ThermoFisher Scientific), 20% KnockOutTM serum replacement (ThermoFisher Scientific), 1% MEM NEAA (Thermo Fisher Scientific), 0.5% L-glutamine (Capricorn), and 0.1 mM β-mercaptoethanol (Thermo Fisher Scientific). UM was changed daily for the following five days to initiate co-differentiation of BCECs and neuronal cells. On day 6, medium was changed to 4 ml/well endothelial cell (EC) medium, composed of Human Endothelial-SFM (ThermoFisher Scientific) and 0.5 % B27 Supplement (Thermo Fisher Scientific), supplemented with 20 ng/ml hFGF and 10 µM retinoic acid (RA) for BCEC expansion. On day 8, BCECs were dissociated using Accutase for 30 min and seeded at a cell density of 1 x 10^6^ cell/cm^2^ onto collagen IV/fibronectin-coated transwell membranes (0.4 µm pore size, 24-well format, Greiner) in EC medium supplemented with 20 ng/mL hFGF and 10 µM RA. BCECs were adapted to EC medium without hFGF and RA at day 9 for 24 h. We measured transendothelial electrical resistance (TEER) values using an electrode to evaluate the integrity of the in vitro BBB. Only BBBs with TEER values ≥ 1000 Ω*cm^2^ at day 10 were included. To monitor monolayer integrity for both iPSC-derived BBB models, 10 µM sodium fluorescein (Sigma-Aldrich) was added to the apical compartment of two reference inserts for each BCEC differentiation. The intensity of the fluorescent tracer molecule was measured in pooled samples of both compartments respectively using a fluorescent plate reader (Infinite M1000 Pro, TECAN, excitation: 490 nm, emission: 525 nm). We applied 10 µM sildenafil, sildenafil citrate, or reference compounds in EC medium to the apical compartment of two inserts each and incubated the cells for 1 h at 37 °C, 5 % CO_2_ on an orbital shaker (300 rpm). Diazepam and atenolol were used as internal controls given their known high and low BBB permeability, as previously demonstrated.^12,13^ Media from both the apical (A) and basolateral (B) compartments were collected, pooled and stored at -80 °C until quantification. Sildenafil and reference compounds were quantified using mass spectrometry. Peak areas were used to calculate the apparent permeability (P_app_) of each substance using equation:

$P_{app}$*=*$\frac{\Delta Q}{\Delta t\times A\left( \frac{C_{D}+C_{0}}{2} \right)\times{10}^{6}}$

$$\Delta Q=Conc. receiver \Delta t$$

$$\Delta t=incubation time$$

$$A=filter surface area [\frac{{cm}^{2}}{well}]$$

$$C_{D}=Conc. donor \Delta t$$

$$C_{0}=Conc. donor t_{0}$$

Quantification of sildenafil, diazepam and atenolol in apical and basolateral media samples was performed using LC-MS/MS. The same instrumentation that was used to determine the Sildenafil blood concentration was used for this purpose. The settings were maintained except for minor adjustments. Chromatographic separation of atenolol (t_R_ = 3.5 min), sildenafil (t_R_ = 4.3 min) and diazepam (t_R_ = 5.5 min) was achieved with gradient elution within 10 min of total run-time. After positive electrospray ionization, the MS/MS detector recorded three mass transitions per drug compound (fragmentor voltage [FV] and collision energies [CE] in parentheses): Sildenafil: *m/z* 475.2 → 58.0 (FV: 220 V, CE: 68 eV, *quantifier*), *m/z* 475.2 → 100.0 (FV: 220 V, CE: 28 eV), *m/z* 475.2 → 283.1 (FV: 220 V, CE: 44 eV); Diazepam: *m/z* 285.1 → 154.1 (FV: 144 V, CE: 28 eV), *m/z* 285.1 → 193.1 (FV: 144 V, CE: 36 eV, *quantifier*), *m/z* 285.1 → 222.1 (FV: 144 V, CE: 28 eV) and Atenolol: *m/z* 267.2 → 56.0 (FV: 128 V, CE: 32 eV), *m/z* 267.2 → 74.1 (FV: 128 V, CE: 24 eV), *m/z* 267.2 → 144.9 (FV: 128 V, CE: 28 eV, *quantifier*). Peak areas were determined with MassHunter Quantitative Analysis software (version 10.1, Agilent Technologies) and the three drugs were quantified using matrix-matched external calibration in the concentration range of 0.001 to 10 µM.

**Supplementary Figures**

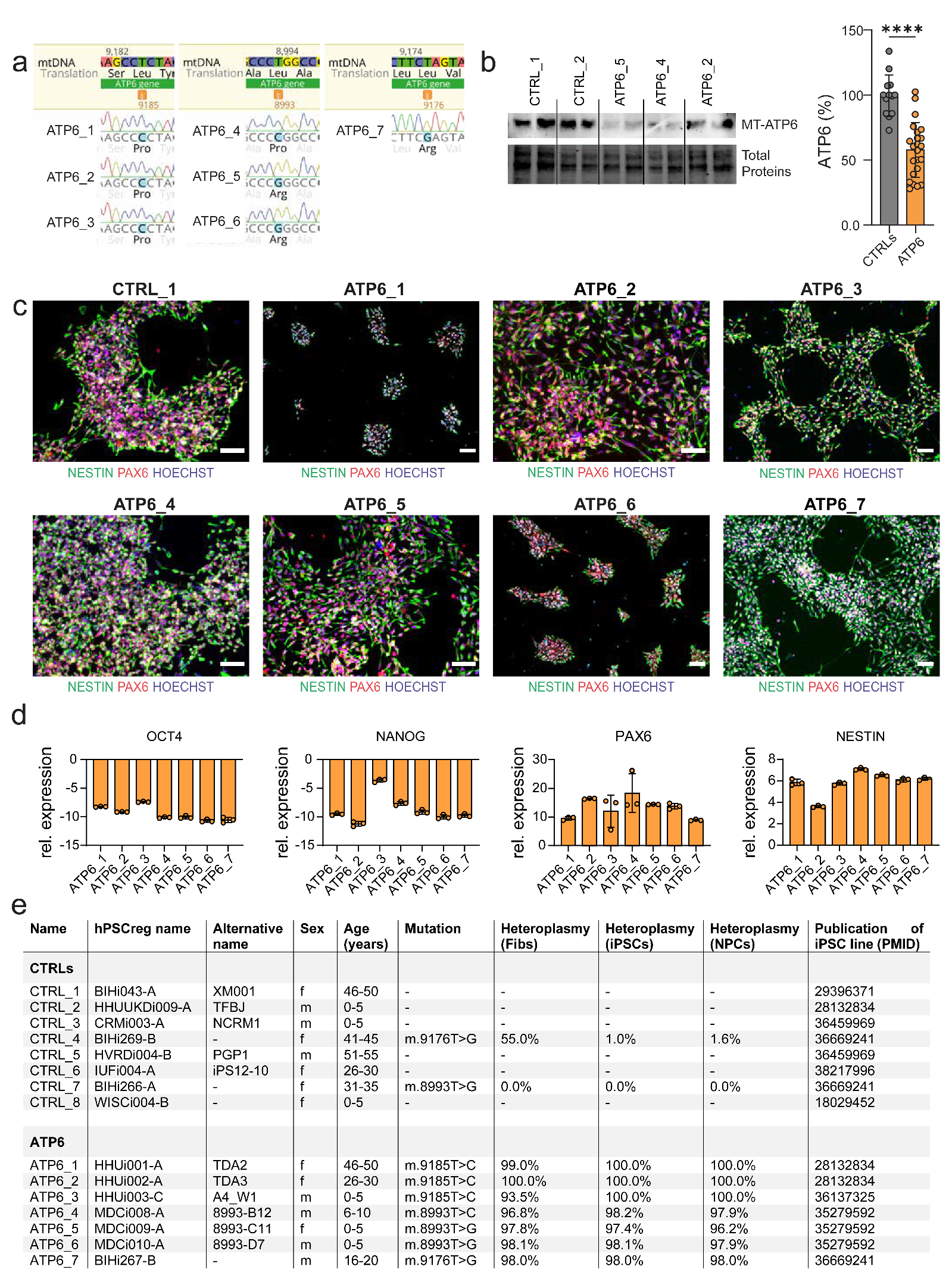

**Figure S1. Generation of LS NPCs (related to Figure 1). a.** Genotyping of LS NPCs by Sanger sequencing confirmed the presence of *MT-ATP6* mutations: m.9176T>C (ATP6_1, ATP6_2, ATP6_3), m.8993T>C (ATP6_4), m.8993T>G (ATP6_5, ATP6_6), and m.9176T>G (ATP6_7). **b.** Representative immunoblot analysis repeated in n=3 independent experiments showing ATP6 protein expression in control NPCs (CTRL_1 and CTRL_2) and LS NPCs representing the four *MT-ATP6* mutations: ATP6_2 for m.9176T>C, ATP6_4 for m.8993T>C, ATP6_5 for m.8993T>G, and ATP6_7 for m.9176T>G. **c.** Representative immunostaining of LS NPCs showing the presence of NPC markers NESTIN and PAX6 similarly to control NPCs. Scale bar: 50 µm. **d.** Gene expression analysis by qPCR showing downregulation of iPSC markers *OCT4* and *NANOG*, and upregulation of NPC markers *NESTIN* and *PAX6* in LS NPCs compared to their respective LS iPSC lines. Relative transcript levels were calculated based on the 2-ΔΔCT method. Data were normalized to the housekeeping gene ornithine decarboxylase antizyme 1 (OAZ1) and presented as mean Log2 ratios (mean +/- SD) in relation to control iPSCs (CTRL_1). **e.** Details of the lines used in this study from controls and from patients with LS carrying *MT-ATP6* mutations including the mutation load (heteroplasmy) in the original fibroblasts (Fibs) and related iPSCs and NPCs.

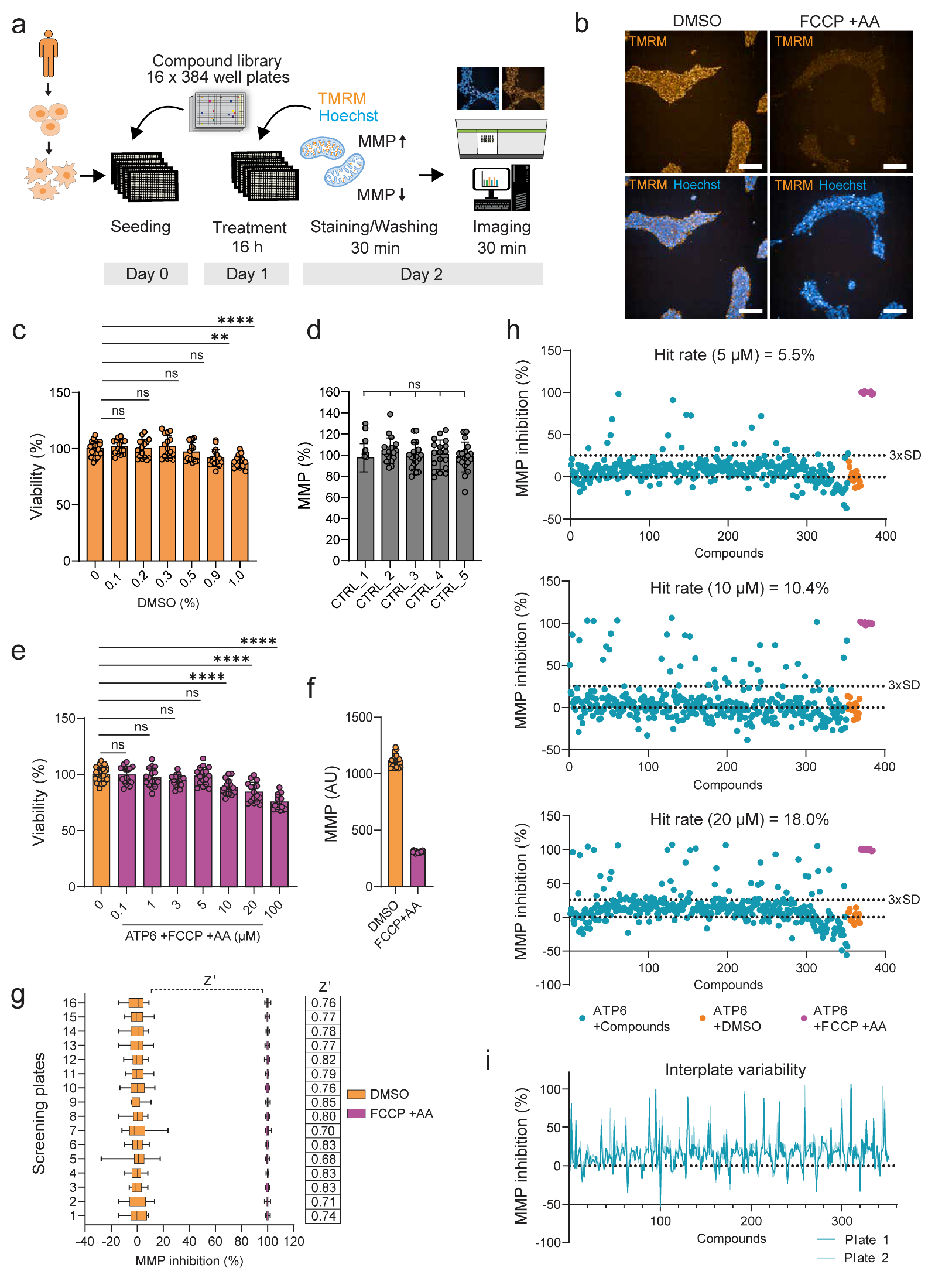
**Figure S2. Compound screening in LS NPCs (related to Figure 1). a.** Schematic of compound screening workflow in LS NPCs using Operetta CLS high-content analysis (HCA) system. **b.** Representative images of LS NPCs (ATP6_2) treated with either DMSO or 5 µM FCCP and 5 µM antimycin A (AA) to uncouple MMP and live-cell stained with the dye TMRM and Hoechst. Images obtained with Operetta CLS microscope using 20 x water objective. Scale bar: 100 µm. **c.** Quantification of cell viability with Cell Titer-Glo assay in LS NPCs (ATP6_2). Dots represent mean values per well. ***p<0.001, ****p<0.0001, ns (not significant); ordinary one-way ANOVA with Dunnett’s multiple comparison. **d.** HCA-based MMP quantification with TMRM in different control NPCs. Dots represent mean values per well out of n=3 independent experiments. ns (not significant); ordinary one-way ANOVA with Dunnett’s multiple comparison. **e.** HCA-based quantification of cell viability using Cell Titer-Glo assay in LS NPCs (ATP6_2) after 16 h treatment with increasing FCCP+AA concentrations normalized to DMSO concentration. Dots represent mean values per well. ****p<0.0001, ns (not significant); ordinary one-way ANOVA with Dunnett’s multiple comparison. **f.** HCA-based MMP quantification with TMRM in LS NPCs (ATP6_2) treated for 16 h with either 0.05 % DMSO or 5 µM FCCP+AA. Dots represent mean values per well. **g.** Assay window for the HCA-based MMP screen in LS NPCs (ATP6_2) in independent screening plates. Z’ factor was calculated as described previously^14^ and remained between 0.68-0.85 across screening plates indicating an excellent assay window. **h.** Screening tests with 16 h treatment of 352 compounds using the HCA-based MMP quantification with TMRM and Hoechst in LS NPCs (ATP6_2). Blue dots: LS NPCs with the compounds dissolved in DMSO; orange dots: LS NPCs treated with only DMSO; purple dots: LS NPCs treated with 5 µM FCCP+AA as positive controls. Upper dotted line marks 3 x DMSO standard deviation (SD) and was set as the threshold for MMP depolarization to calculate and compare hit rates using different compound concentrations. **i.** HCA-based MMP quantification with TMRM and Hoechst in LS NPCs (ATP6_2) with 352 compounds (10 µM after 16 h) demonstrating good reproducibility of independent screening plates.

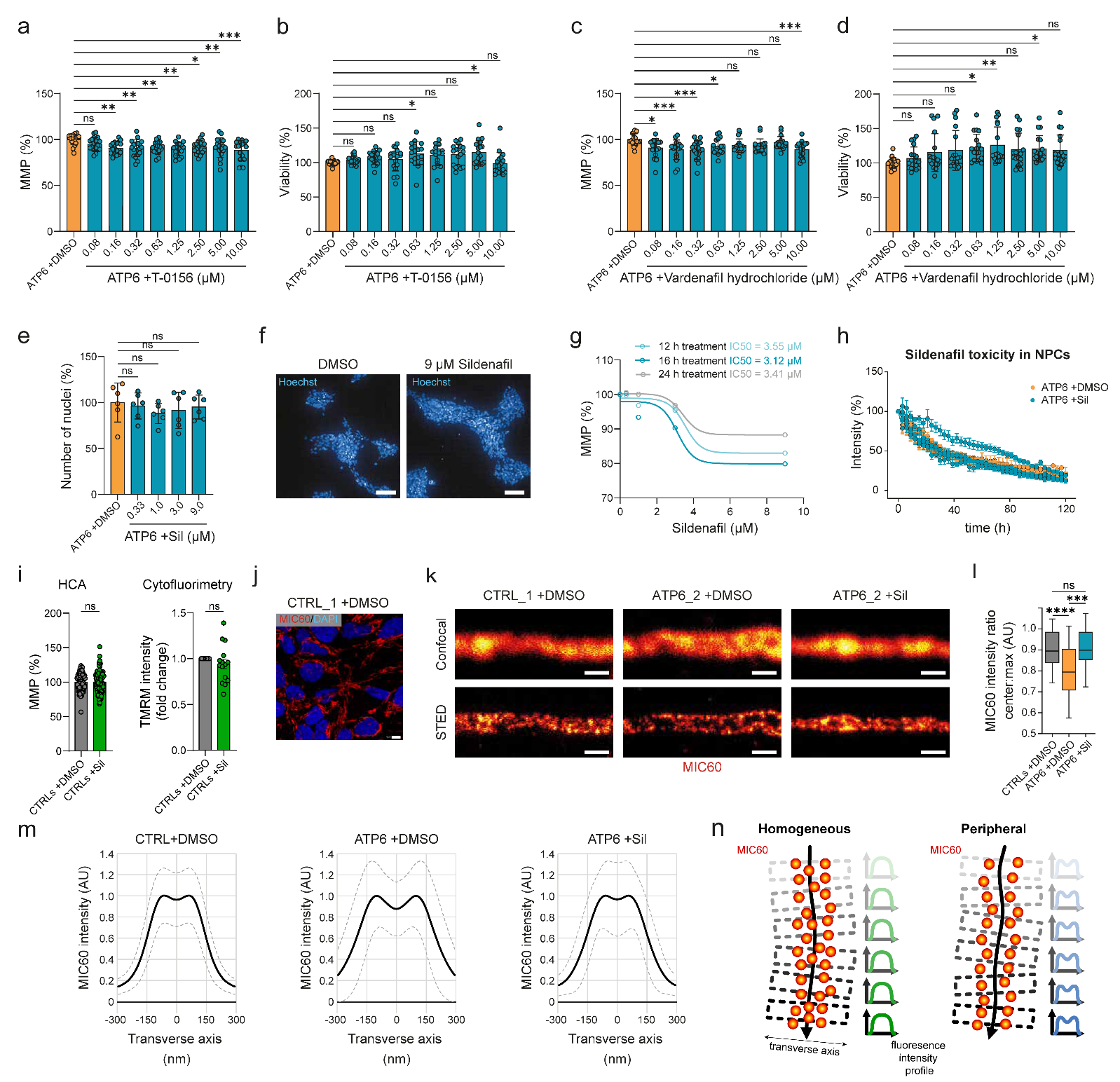
**Figure S3. PDE5 inhibitors as leads in normalizing MMP in LS NPCs (related to Figure 1). a-d.** HCA-based MMP quantification with TMRM and cell viability using Cell Titer-Glo assay in LS NPCs (ATP6_2) treated with increasing concentrations of PDE5i T-0156 or vardenafil hydrochloride compared to DMSO only. Dots represent mean values per well out of n=3 independent experiments. *p<0.05, **p<0.01, ***p<0.001, ns (not significant); ordinary one-way ANOVA with Dunnett’s multiple comparison. **e.** HCA-based cell viability with Hoechst in LS NPCs (ATP6_2) treated with increasing concentrations of PDE5i sildenafil. Dots represent mean values per well. ns (not significant); ordinary one-way ANOVA with Dunnett’s multiple comparison. **f.** Representative images of LS NPCs (ATP6_2) treated with either 0.1 % DMSO or 9 µM sildenafil (in a final concentration of 0.1 % DMSO) for 16 h stained with Hoechst. Images obtained with Operetta CLS microscope using 20 x objective. Scale bar: 100 µm. **g.** Half-maximal inhibitory concentration (IC50) values calculated in LS NPCs (ATP6_2) treated with 0 (0.1 % DMSO), 0.33, 3 or 9 µM sildenafil for 12 h (light blue), 16 h (black), or 24 h (dark blue). Data represents mean values of 6 wells each. **h.** Toxicity assessment of sildenafil in LS NPCs (ATP6_2, ATP6_4, ATP6_5 and ATP6_7) either treated with 0.1 % DMSO (orange) or 10 µM sildenafil in 0.1 % DMSO (cyan) using Cellcyte live-cell analyzer. Dots and bars (SD) represent individual signal for each line of n=3 independent experiments. **i.** MMP quantification by either HCA or cytofluorimetry in control NPCs (CTRL_1) treated with 10 µM sildenafil for 16 h. **j.** Representative image of control NPCs (CTRL_1) stained with MIC60 and DAPI. Scale bar: 5 µm. **k.** Representative images of MIC60 staining taken by confocal or stimulated emission depletion microscopy (STED) in control NPCs (CTRL_1) and LS NPCs (ATP6_2) treated with either DMSO or 10 µM sildenafil for 16 h. Scale bar: 500 nm. **l.** Ratio of MIC60 central fluorescence intensity to MICO60 maximal fluorescence intensity recorded with STED microscopy for the transversal profiles shown in m. ***p<0.001, ****p<0.0001, ns; ordinary one-way ANOVA with Dunnett’s multiple comparison. **m.** MIC60 average fluorescence intensity profiles across the transverse axis of mitochondria recorded with STED microscopy in control NPCs (CTRL_1) and LS NPCs (ATP6_2) treated with either DMSO or 1 µM sildenafil for 24 h. The mean transversal profile is shown in black; dashed lines indicate SD. **n.** Cartoon depicting the distribution pattern of MIC60 in mitochondrial cristae. Left, homogeneous distribution resulting in a fluorescence intensity profile with a broad plateau at the central maximum along the transversal mitochondrial axis was seen in control NPCs and in LS NPCs treated with sildenafil. Right, peripheral distribution of MIC60 was seen in untreated LS NPCs.

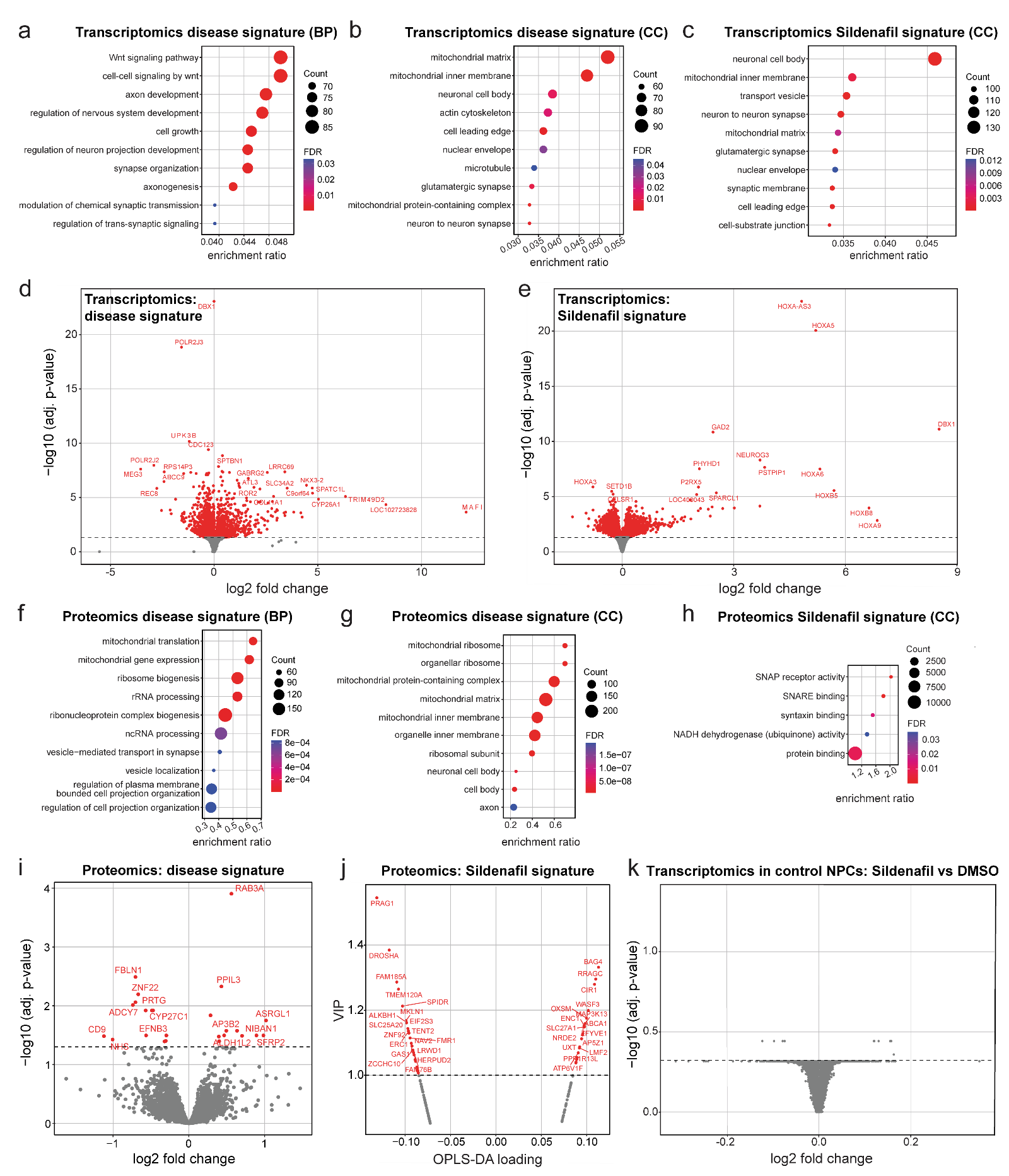
**Figure S4. Disease signature and sildenafil signature unveiled by multi-omics (related to Figure 2). a-b.** Dot plot highlighting top ten enriched Gene Ontology (GO) biological processes (BP) and GO cellular components (CC) in LS NPCs (ATP6_2, ATP_4, ATP6_5, ATP6_7) compared to control NPCs (CTRL_1, CTRL_2, CTRL_3, CTRL_4) based on transcriptomics. **c.** Dot plot highlighting top ten enriched GO CC in ATP6 NPCs (ATP6_2, ATP_4, ATP6_5, ATP6_7) treated with sildenafil for 16 h compared to DMSO based on transcriptomics. **d-e.** Volcano plots of bulk transcriptomics depicting genes differentially regulated (red dots) in LS NPCs (ATP6_2, ATP_4, ATP6_5, ATP6_7) compared to control NPCs (CTRL_1, CTRL_2, CTRL_3, CTRL_4) or in LS NPCs (ATP6_2, ATP_4, ATP6_5, ATP6_7) treated with sildenafil compared to DMSO. **f-g.** Dot plot highlighting top ten enriched GO BP and GO CC in LS NPCs (ATP6_2, ATP_4, ATP6_5, ATP6_7) compared to control NPCs (CTRL_1, CTRL_2, CTRL_3, CTRL_4) based on proteomics. **h.** Dot plot highlighting top ten enriched GO CC in LS NPCs (ATP6_2, ATP_4, ATP6_5, ATP6_7) treated with sildenafil for 16 h compared to DMSO based on proteomics. **i-j.** Volcano plots of proteomics depicting proteins differentially regulated (red dots) in LS NPCs (ATP6_2, ATP_4, ATP6_5, ATP6_7) compared to control NPCs (CTRL_1, CTRL_2, CTRL_3, CTRL_4) or in LS NPCs (ATP6_2, ATP_4, ATP6_5, ATP6_7) treated with sildenafil compared to DMSO. **k.** Volcano plot of bulk transcriptomics of control NPCs (CTRL_1, CTRL_2, CTRL_3, CTRL_4) treated with sildenafil compared to DMSO.

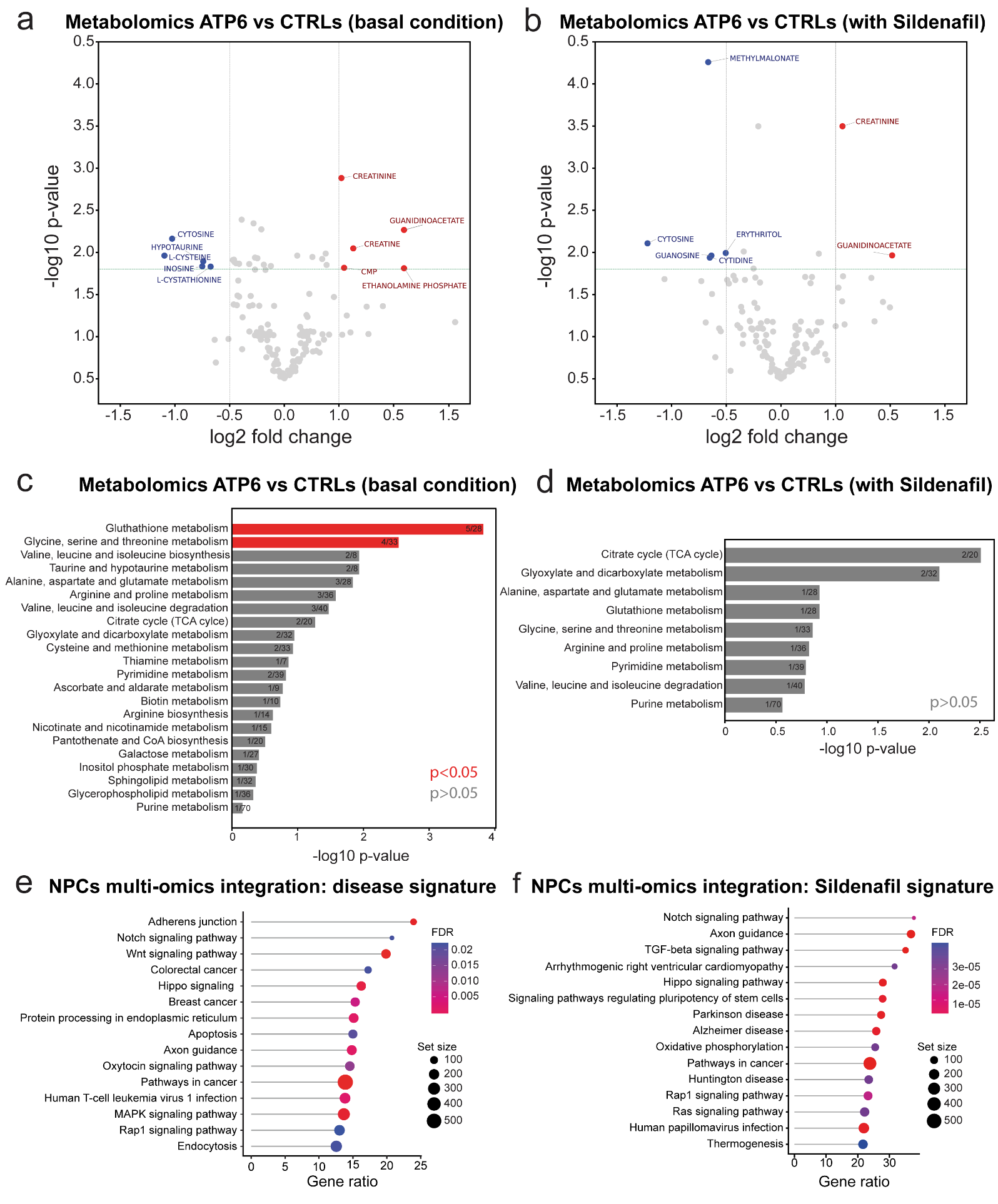
**Figure S5. Metabolic signature of LS NPCs and multi-omics integration (related to Figure 2). a-b.** Volcano plots of metabolomics depicting upregulated metabolites (red) and downregulated metabolites (blue) in LS NPCs (ATP6_2, ATP_4, ATP6_5, ATP6_7) compared to control NPCs (CTRL_1, CTRL_2, CTRL_3, CTRL_4) either in basal DMSO conditions or after 16 h treatment with 10 µM sildenafil. **c-d.** Enrichment analysis of altered metabolic pathways (red) and identified metabolic pathways (gray) in LS NPCs (ATP6_2, ATP_4, ATP6_5, ATP6_7) compared to control NPCs (CTRL_1, CTRL_2, CTRL_3, CTRL_4) either in basal DMSO conditions or after 16 h treatment with 10 µM sildenafil. **e-f.** Multi-omics integration highlighting KEGG pathways modified in LS NPCs compared to control NPCs (disease signature) or LS NPCs treated with sildenafil compared to DMSO (sildenafil signature).

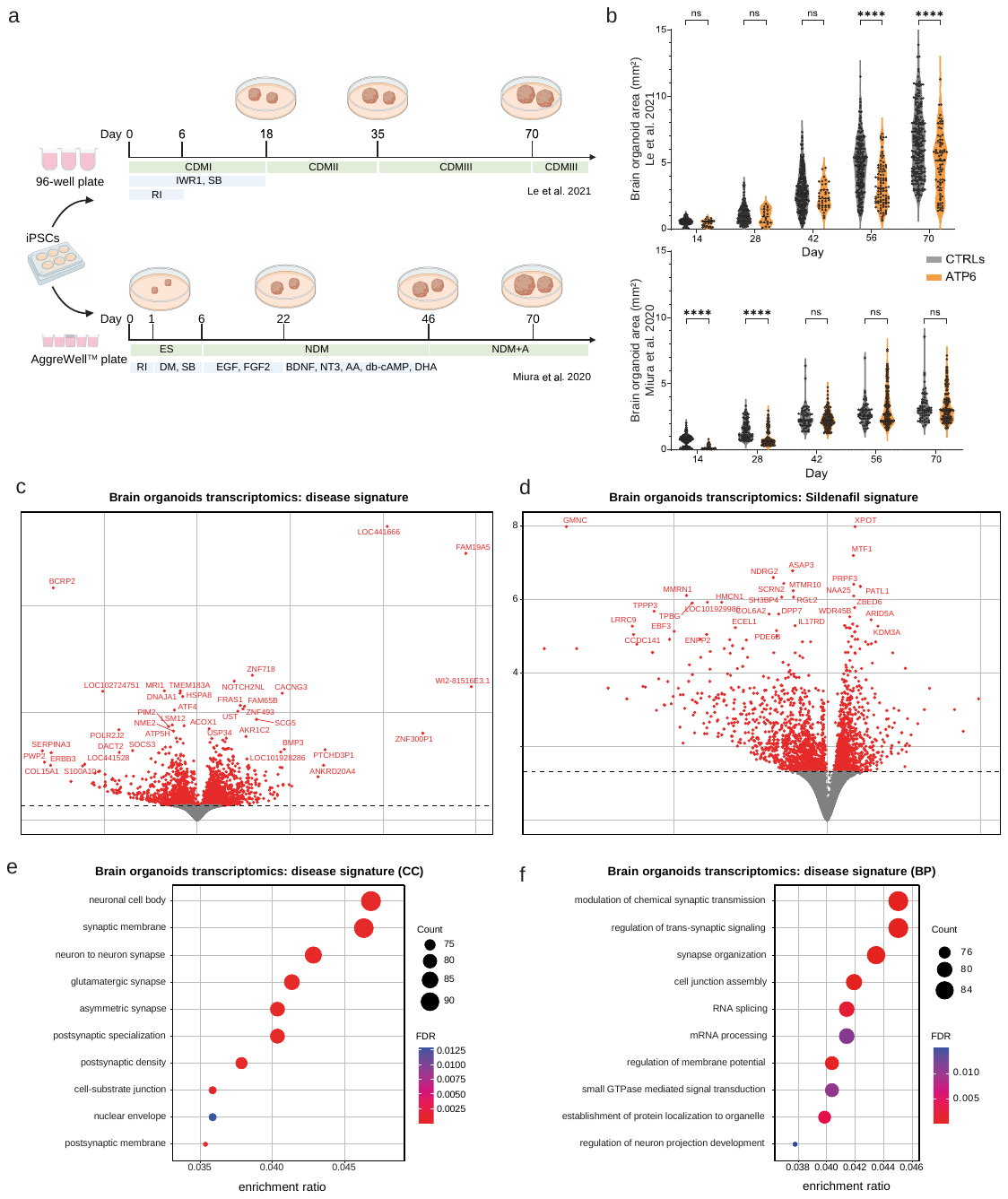
**Figure S6. Brain organoid models of LS (related to Figure 2). a.** Schematic of the two protocols employed for the generation of cortical brain organoids: Le et al with 96-well plates^15^ or Miura et al with AggreWells plates^16^. **b.** Quantification of the size of individual brain organoids grown following the two protocols until day 70. ****p<0.0001, ns (not significant); two-tailed Mann-Whitney U test. **c-d.** Volcano plots of bulk transcriptomics depicting differentially expressed genes (red dots) in day 70 LS brain organoids (ATP6_2, ATP6_7) compared to control brain organoids (CTRL_1, CTRL_2) or in day 70 LS brain organoids (ATP6_2, ATP6_7) treated with 10 µM sildenafil for 24 h compared to DMSO. Fold change (FC). **e-f.** Dot plots highlighting top ten enriched GO BP and GO CC in day 70 LS brain organoids (ATP6_2, ATP6_7) compared to control brain organoids (CTRL_1, CTRL_2) based on bulk transcriptomics.

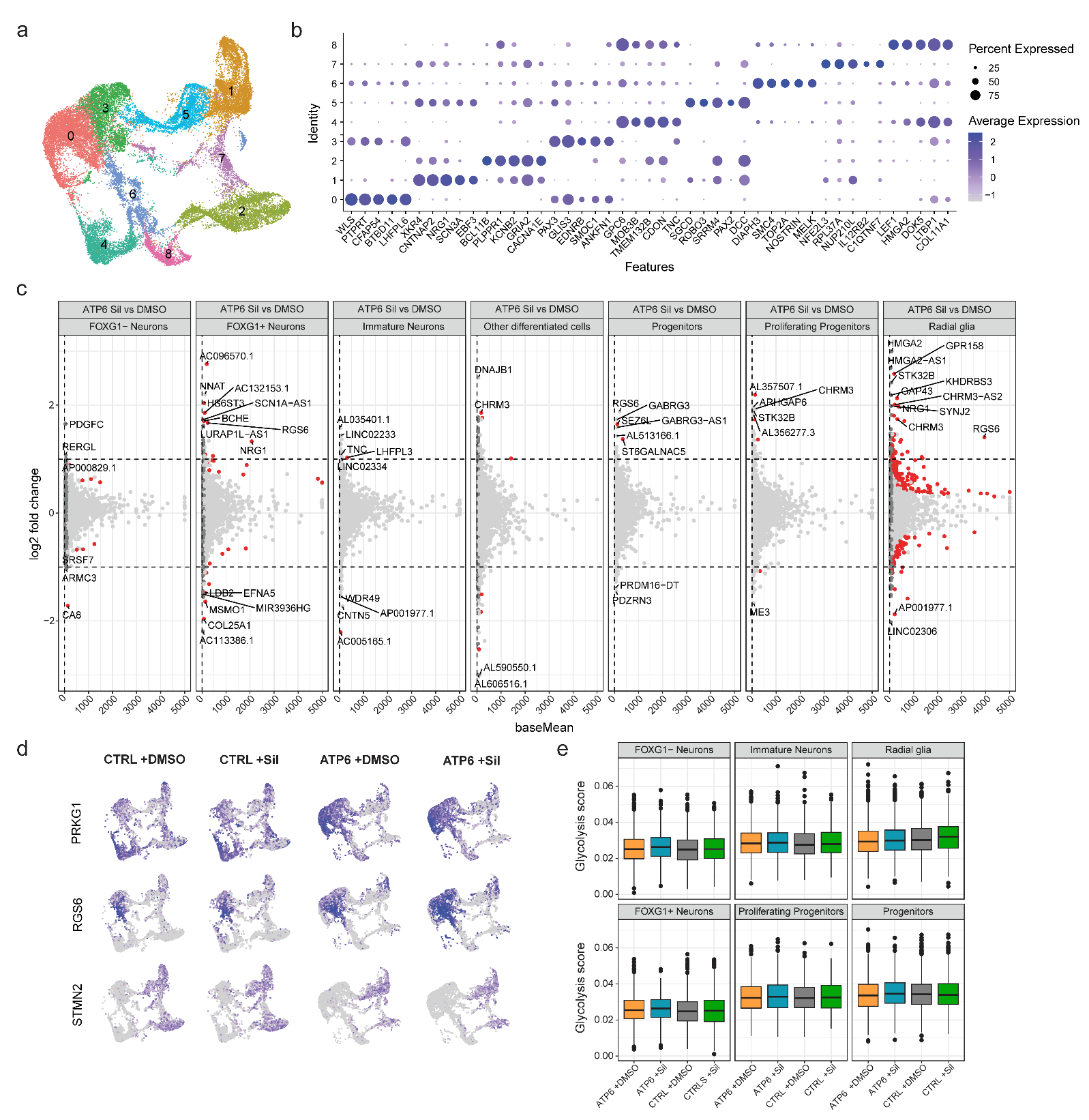
**Figure S7. Single-nucleus RNA sequencing (snRNAseq) of LS brain organoids (related to Figure 3). a.** UMAP representation of 9 clusters within day 70 cortical brain organoids. **b.** Dot plot featuring five representative genes for each of the 9 clusters present within day 70 cortical brain organoids **(Table S5)**. **c.** Pseudo-bulk analysis of day 70 LS brain organoids (ATP6_7) treated with either 10 µM sildenafil or DMSO for 45 days for each of the 7 populations composing day 70 brain organoids. Volcano plots with red dots indicating differentially expressed genes. **d.** UMAP representation of the gene expression distribution of *PRKG1*, *RGS6* and *STMN2* within day 70 brain organoids from controls (CTRL_1) and LS (ATP6_7) treated with either sildenafil or DMSO for 45 days. **e.** Molecular signatures database hallmark glycolysis score across the 7 populations of day 70 brain organoids.

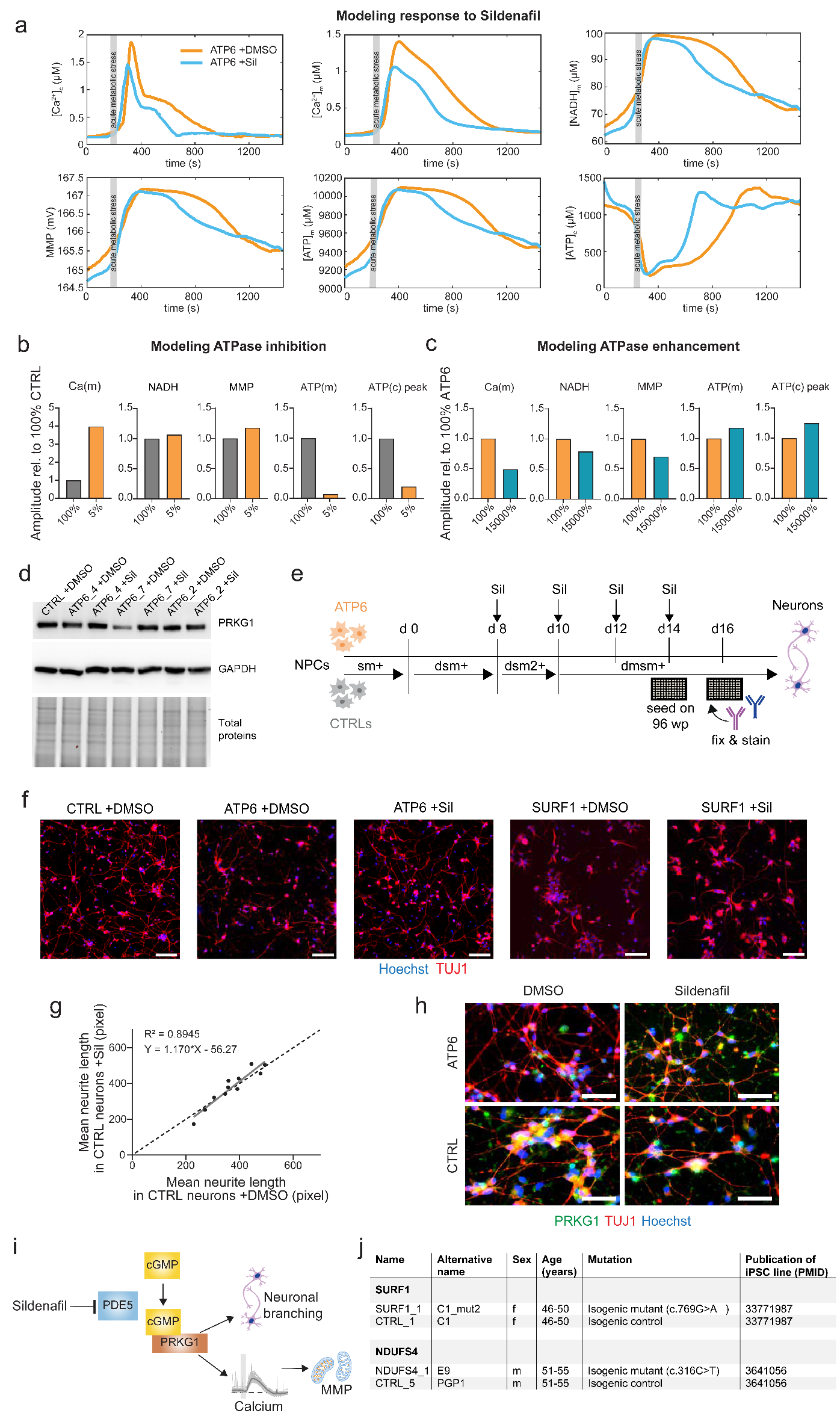

**Figure S8. Functional validations in human neuronal models of LS (related to Figure 4). a.** Computational model of mitochondrial metabolism based on calcium traces measured in cortical brain organoid slices (cBOS) **(Figure 4b)** describing the effect of sildenafil treatment on mitochondrial parameters in LS cBOS. **b.** Computational model of ATPase enhancement (orange bars) of control cBOS (gray bars) based on calcium traces of cBOS. The model showed that decreasing complex V activity in control cells leads to effects on cellular bioenergetics similar to those we have experimentally measured in LS NPCs. **c.** Computational model of ATPase enhancement (cyan bars) of ATP6-mutant cBOS (orange bars) based on calcium traces of cBOS. The model suggests that increasing complex V activity of ATP6-mutant cells leads to effects on cellular bioenergetics similar to those we have experimentally measured in LS NPCs treated with sildenafil. Hence, the calcium response elicited by sildenafil in mutant cells may potentially represent a primary mechanism of action of sildenafil that would in turn lead to beneficial bioenergetic consequences. **d.** Representative immunoblot of PRKG1 in control NPCs and LS NPCs treated with 10 µM sildenafil or DMSO for 16 h. The image refers to the quantification reported in **Figure 4h**. **e.** Schematic of the differentiation into dopaminergic-enriched neuronal cultures to assess the effects of sildenafil treatment. **f.** Representative images of day 16 dopaminergic-enriched neurons from controls (CTRL_1), MT-APT6 mutants (ATP6_7), and SURF1 mutants treated with 10 µM sildenafil or DMSO for 8 days. Scale bar: 100 µm. **g.** Linear regression of the mean neurite length (mean over median) in control neurons (CTRL_1, n=4; CTRL_2, n=3; CTRL_3, n=1; CTRL_5, n=3; n=number of independent experiments) treated with sildenafil for 8 days over control neurons exposed to only DMSO (grey line). Black dotted line shows the hypothetical linear regression in which no effect is seen due to sildenafil treatment (y=x). Dots represent individual values per experiment out of n=10 independent experiments. **h.** Exemplary immunostaining for PRKG1 and TUJ1 in day 16 control neurons (CTRL_2) and LS neurons (ATP6_4) treated for 8 days with sildenafil or DMSO. Scale bar: 50 µm. **i.** Cartoon of suggested mode of action of sildenafil in LS neural cells. Following PDE5 inhibition and accumulation of cGMP, the activation of PRKG1 could lead on the one hand to modulate calcium levels with consequent impact on cellular bioenergetics and on the other to enhance neuronal branching. **j.** Details of the additional LS iPSC lines used in this study carrying mutations in the gene *SURF1* or *NDUFS4* and their respective isogenic controls.

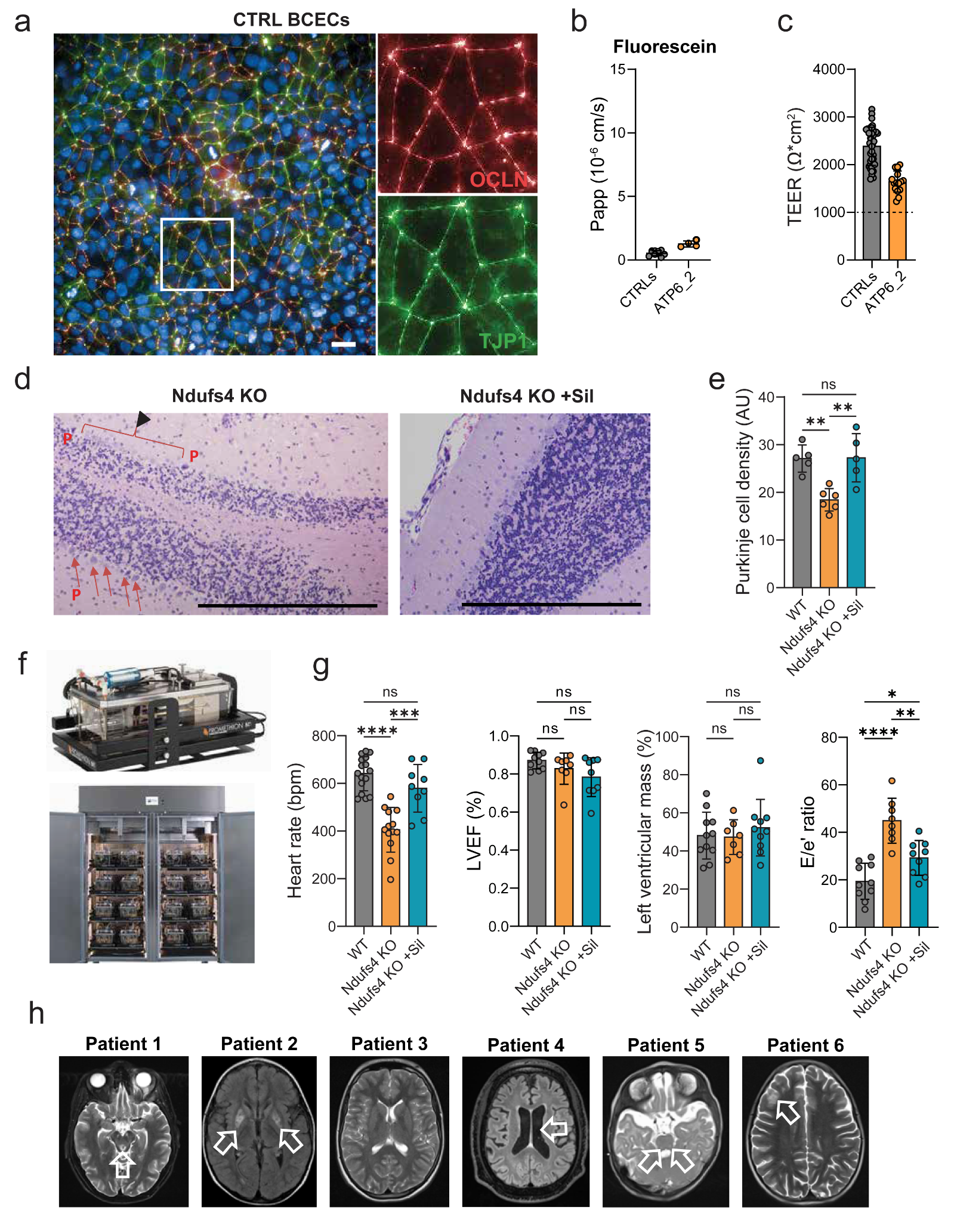

**Figure S9. Sildenafil impact on human BBB model, *Ndufs4* KO mice, and LS patients (related to Figure 4 and 5). a.** Representative immunostaining of brain capillary endothelial cells (BCECs) derived from control iPSCs (CTRL_8) showing expression and co-localization of occludin (OCLN) and tight junction protein 1 (TJP1). Scale bar: 20 µm. **b.** Apparent permeability coefficient (P_app_ [10^-6^ * cm/s]) from apical (A) to basolateral (B) compartment of fluorescein in control BCECs (CTRL_8) and LS BCECs (ATP6_2) in n=3-5 independent experiments. **c.** Transendothelial electrical resistance (TEER) measurement in BCECs derived from control iPSCs (CTRL_8) and LS iPSCs (ATP6_2) in n=3-5 independent experiments on day 10 of differentiation. **d-e.** Representative immunohistochemistry and quantification for Purkinje cells (P) density in 42-45-day-old *Ndufs4* KO mice (n=5-6). Arrows indicate loss of Purkinje cells. **p<0.01, ns (not significant); ANOVA post-hoc analysis. **f.** Promethion metabolic chamber used to assess the metabolic fitness of *Ndufs4* KO mice **(Figure 5c) g.** Cardiac effects of sildenafil treatment in 42-45-day-old *Ndufs4* KO mice (n=8-10): heart rate (bpm) and diastolic dysfunction measured by tissue Doppler imaging, left ventricular mass (%), left ventricular ejection fraction (LVEF, %), and ratio of the peak early mitral inflow velocity (E) over the early diastolic mitral annular velocity (e′) measured by echocardiography. *p<0.05, **p<0.01, ***p<0.001, ****p<0.001, ns (not significant); two-tailed Mann-Whitney U test. **h.** Cranial magnetic resonance imaging (cMRI) of the six patients undergoing off-label compassionate treatment with sildenafil. Arrows indicate lesions in the basal ganglia lesions and brainstem characteristic of LS (see supplementary methods for details).

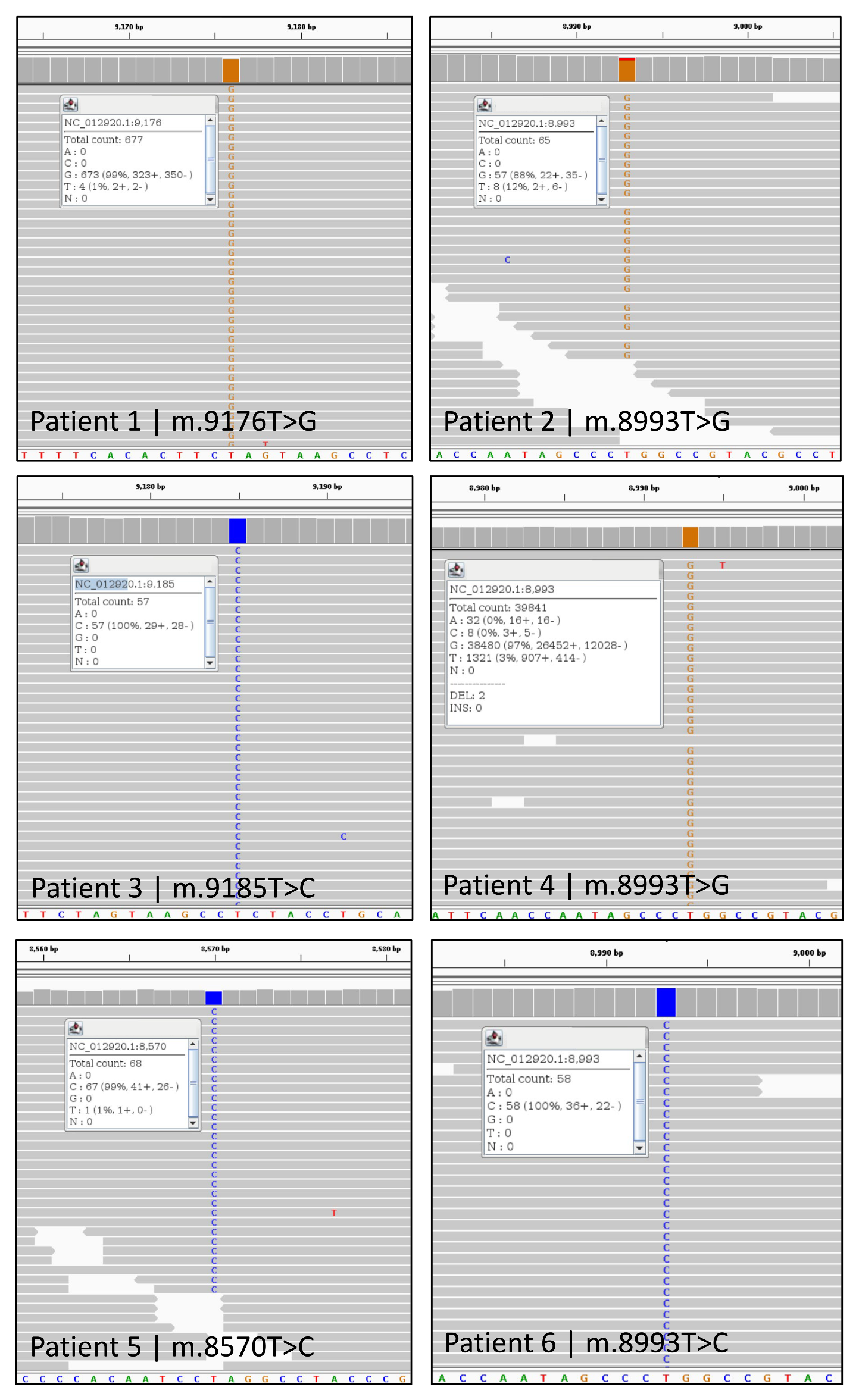
**Figure S10. Genetic diagnosis of the six patients with LS treated with sildenafil (related to Figure 5).** *MT-ATP6* mutations and degrees of heteroplasmy in the Whole Exome Sequencing datasets of the 6 patients with LS treated with sildenafil. Data are visualized with the IGV viewer (v2.16).

**Supplementary Tables**

**Table S1.** Bulk transcriptomics of LS NPCs and control NPCs showing differentially expressed genes (DEG).

**Table S2.** Proteomics of LS NPCs and control NPCs showing differentially expressed proteins (DEP).

**Table S3.** Metabolomics of LS NPCs and control NPCs showing differential metabolites (DM).

**Table S4.** Bulk transcriptomics of cortical brain organoids from LS patients and controls and acute sildenafil treatment showing differentially expressed genes (DEG).

**Table S5.** Clusters identified by single-nucleus RNA sequencing (snRNAseq) of cortical brain organoids from LS patients and controls.

**Table S6.** Pseudo-bulk analysis of snRNAseq datasets of cortical brain organoids from LS patients and controls and chronic sildenafil treatment showing differentially expressed genes (DEG) within the different brain organoid populations.

**Table S7.** List of parameters employed for the computational mitochondrial model based on calcium responses of cortical brain organoid slices (cBOS).

**Table S8.** Clinical outcomes in six LS patients carrying *MT-ATP6* mutations upon treatment with sildenafil.

**Table S9.** Questionnaire on side effects of continuous sildenafil use in six LS patients treated with sildenafil.
