## Supplementary material for "Pluripotent stem cell-based drug discovery uncovers sildenafil as a treatment for mitochondrial disease": Table S7

**Table S7:** List of parameters for the computational mitochondrial model (equations 2 – 20 in the Methods)

| **Parameter** | **Description** | **Value (Unit)** | **References** |
| --- | --- | --- | --- |
| c_mito_ | Mitochondrial inner membrane divided by Faraday constant | 1.8 (µM.mV ^-1^) | ^1^ |
| f_m_ | Fraction of free over buffer-bound [Ca^2+^] in mitochondria | 0.00025 | This work |
| L | Allosteric equilibrium constant for uniporter conformations | 50 | ^2^ |
| α_c_ | Cytosolic ADP and ATP buffering coefficient | 0.111 | ^3^ |
| α_m_ | Mitochondrial ADP and ATP buffering coefficient | 0.139 | ^3^ |
| v_AGC_ | Maximum rate constant of NADH production via malate-aspartate shuttle | 25 (µMs^-1^) | ^4^ |
| v_MCU_ | Maximum rate constant of the MCU | 0.00001 (µMs^-1^) | ^4^ |
| v_NCX_ | Maximum rate constant of the NCX | 0.00035 (µMs^-1^) | ^4^ |
| v_F1F0_ | Maximum rate constant of the F1F0 ATPase | Varies (µMs^-1^) | This work |
| v_ANT_ | Maximum rate constant of the Adenine Nucleotide Translocator | 5000 (µMs^-1^) | ^1,5^ |
| v_p_ | Maximum rate constant of the SERCA pumps | 120 (µMs^-1^) | ^6^ |
| $A_{c}^{TOT}$ | Total concentration of cytosolic adenine nucleotides | 4000 (µM) | This work |
| $A_{m}^{TOT}$ | Total concentration of mitochondrial adenine nucleotides | 15000 (µM) | ^1^ |
| ${NAD}_{m}^{TOT}$ | Total concentration of mitochondrial pyridine nucleotides | 250 (µM) | ^4^ |
| K_1_ | Dissociation constant for [Ca^2+^] translocation by MCU | 6 (µM) | ^4^ |
| K_2_ | Dissociation constant for MCU activation by [Ca^2+^] | 0.38 (µM) | ^2^ |
| K_p_ | Dissociation constant of [Ca^2+^] from SERCA | 0.35 (µM) | ^6^ |
| K_h_ | Michaelis-Menten constant for ATP hydrolysis | 1000 (µM) | ^4^ |
| k_AGC_ | Dissociation constant of [Ca^2+^] from AGC | 0.14 (µM) | ^7^ |
| k_e_ | Dissociation constant of ATP from SERCA pumps | 0.05 (µM) | ^4,8^ |
| k_o_ | Rate constant of NADH oxidation by ETC | Varies (µMs^-1^) | This work |
| k_GLY_ | Velocity of glycolysis (empirical) | Varies (µMs^-1^) | This work |
| k_HYD_ | Maximum rate of ATP hydrolysis | 100 (µMs^-1^) | ^4^ |
| k_x_ | Maximum rate constant of bidirectional [Ca^2+^] leak from mitochondria | 0.008 (s^-1^) | ^4^ |
| b_1_ | Scaling factor between NADH consumption and change in membrane voltage | Varies | This work |
| b_2_ | Scaling factor between ATP production by ATPase and change in membrane voltage | 3.43 | ^1^ |
| δ | Ratio between mitochondrial volume to cytosolic volume | 0.0733 | ^1^ |
| p_1_ | Voltage dependence coefficient of MCU activity | 0.1 (mV ^-1^) | ^4^ |
| p_2_ | Voltage dependence coefficient of NCX activity | 0.016 (mV^-1^) | ^4^ |
| p_3_ | Voltage dependence coefficient of [Ca^2+^] leak | 0.05 (mV^-1^) | ^4^ |
| p_4_ | Voltage dependence coefficient of AGC activity | 0.01 (mV^-1^) | ^4^ |
| F | Faraday’s constant | 96480 (Cmol^-1^) |  |
| T | Temperature | 310.16 (K) |  |
| R | Ideal gas constant | 8315 (mJ.mol^-1^ K^-1^) |  |
| q_1_ | Michaelis-Menten-like constant for NAD^+^ consumption by the TCA cycle | 1 (µM) | ^1^ |
| q_2_ | Half-maximal activating of the TCA cycle by mitochondrial [Ca^2+^] | 0.1 (µM) | ^4^ |
|  | Half-maximal activating for indirect inhibition of the AGC by cytosolic [Ca^2+^] | 0.1 (µM) | ^4^ |
| q_3_ | Michaelis-Menten constant for NADH consumption by the ETC | 100 (µM) | ^1^ |
| q_4_ | Voltage dependence coefficient 1 of ETC activity | 177 (mV) | ^1^ |
| q_5_ | Voltage dependence coefficient 2 of ETC activity | 5 (mV) | ^1^ |
| q_6_ | Inhibition constant of ATPase activity by ATP | 10000 (µM) | ^1^ |
| q_7_ | Voltage dependence coefficient of ATPase activity | 190 (mV) | ^1^ |
| q_8_ | Voltage dependence coefficient of ATPase activity | 8.5 (mV) | ^1^ |
| q_9_ | Voltage dependence of the proton leak | 2 (µMs^-1^mV^-1^) | ^1^ |
| q_10_ | Rate constant of voltage-independent proton leak | -30 (µMs^-1^) | ^1^ |
