## Supplementary material for "Pluripotent stem cell-based drug discovery uncovers sildenafil as a treatment for mitochondrial disease": Table S8

**Table S8.** Clinical outcomes in six LS patients carrying *MT-ATP6* mutations upon treatment with sildenafil.

| **Patient 1** | |
| --- | --- |
| Age range, sex | 16-20 years, male |
| Mutation | *MT-ATP6* m.9176T>G (99.4 % heteroplasmy) |
| Past medical history | Mildly globally delayed early childhood development, tendency to walk insecurely (ataxia), delayed speech development and dysarthria. At 10-15 years of age, diagnosis of a combined axonal and demyelinating neuropathy and increasing lower limb weakness. |
| Symptoms | At 16-20 years of age, a metabolic crisis developed with encephalopathy, cerebral seizures, severe motor-sensory neuropathy predominantly in the legs, mechanical ventilation due to respiratory insufficiency, cardiomyopathy (LVEF around 45 %), nutrition only possible via PEG feeding tube. The motor-sensory neuropathy had been preexistent and had impeded his walking ability, which was, however, possible with gait ataxia. |
| Sildenafil Doses | 3 x 40 mg (2 mg/kg body weight (BW)/d in 3 single doses) |
| **Treatment progress:** treatment started in December 2018 and is ongoing | |
| After 1 month | Extubation possible, can breathe independently. |
| After 3 months | Sitting stably, movements against gravity possible, stable respiration, ECHO cardiography left ventricle ejection fraction (LVEF) increase to 65 %. |
| After 5 months | Tracheostomy closure and percutaneous gastrostomy (PEG) feeding tube removal, EEG normal, discontinuation of antiepileptic drugs. |
| After 18 months | Independently moves while sitting in the wheelchair, even manages to climb curbstones with the wheelchair and maneuver on ascending terrain. |
| After 30 months | Lifts himself into a wheelchair, walks at times with a walker with forearm supports, medication was very well tolerated throughout the period, echocardiographic investigations have been normal the entire time, cerebral seizures did not recur. Peripheral neuropathy did not change. |
| After 3 years | The patient and his parents report that he had been completely unable to move his legs for half a day, which later gradually improved without intervention (there was no trigger, infection, or fever). These episodes had occurred frequently since 5-10 years of age. Before the start of the sildenafil therapy these episodes had occurred every 2 months and they did not present anymore after the initiation of sildenafil therapy. As the patient had increased in body weight, the dosage was adapted to 2 mg/kg BW/d. |
| Last assessment after 6 years | The patient has remained stable with no further metabolic crises despite two COVID19 infections. He is able to move independently in his wheelchair and he can use a walker with arm support indoors. He can eat a normal diet. He complained about palpitations, but echocardiography and ECG-monitoring over 24 hours was normal in December 2024. Neuropathy and dysarthria have not improved. |
| **Patient 2** | |
| Age range, sex | 10-15 years, male |
| Mutation | *MT-ATP6* m.8993T>G (87.7 % heteroplasmy) |
| Past medical history | The motor development of the patient was delayed. He was always ataxic and fell frequently. Heart, eye, and hearing examinations were always normal. |
| Symptoms | Parents report that about twice a year there were situations in which he acutely decompensated during febrile illnesses. This was accompanied by severe vomiting and deterioration of speech with slurred articulation (transient aphasia). |
| Sildenafil Doses | 3 x 20 mg (1.2 mg/kg BW/d in 3 single doses) |
| **Treatment progress:** treatment started in October 2019 and is ongoing | |
| After 2 months | Increase in free walking distance from 500 m to > 2,000 m, significantly less exhaustible, increase in speed and safety in the Nine-Hole-Peg Test. Medication is well tolerated, normal cardiac examination findings |
| Last assessment after 5 years | No more metabolic crisis recorded to date. Parents describe their child as having considerably more energy over the day in comparison to his state before initiation of sildenafil treatment. He can now stay up longer and does not become sleepy over the day as before. His ataxia has improved and his tendency to stumble and fall down is greatly diminished. He tolerates the medication well without unwanted side effects. He can now walk up to 5 km without longer pausing. Normal cardiologic function as assessed by echocardiography and ECG monitoring in June 2024. His SARA score improved from 15.5 (year 2018) to 11 (year 2024). |
| **Patient 3** | |
| Age range, sex | 21-25 years, female |
| Mutation | *MT-ATP6* m.9185T>C (100 % homoplasmic) |
| Past medical history | Mildly delayed motor development, first metabolic crisis at the 0-5 years of age during a viral infection with loss of ambulation. These incidences increased in frequency and lasted from 3 hours to 3 days. Later on, she developed axonal neuropathy and progressive dependence from a wheelchair. |
| Symptoms | Relapsing episodes of muscle weakness and paralysis (4-5 per week), severe demyelinating neuropathy, urinary incontinence, chronic metabolic acidosis, exercise insufficiency. |
| Sildenafil Doses | 3 x 25 mg (1.4 mg/kg BW/d in 3 single doses) slow increase up to 2.9 mg/kg BW/d |
| **Treatment progress:** treatment started in May 2019 and has been discontinued in November 2020 | |
| After 6 months | Significant decrease in paralytic episodes and absence of muscle cramps. The medication is well tolerated. The motor neuropathy did not improve. |
| After 12 months | The patient has experienced an itchy drug rash predominantly in the face and trunk as an allergic reaction towards sildenafil and it was decided to discontinue the sildenafil medication. However, the symptoms of muscle cramps and paralytic episodes returned. |
| Last assessment after 18 months | The sildenafil medication had been reinitiated and the symptoms of the intermittent paraplegia improved. However, also the drug rash reappeared. Thus, the sildenafil medication was discontinued. |
| **Patient 4** | |
| Age range, sex | 35-40 years, male |
| Mutation | *MT-ATP6* m.8993T>G (96.7 % heteroplasmy) |
| Past medical history | Psychomotor developmental delay, progressive ataxia with loss of independent walking at 10-5 years of age, and loss of head control and capability to sit independently at 30-35 years of age. |
| Symptoms | Chronic condition with progressive worsening of motor function. |
| Sildenafil Doses | May 2022, starting with 3 x 10 mg/d, increased dosage to actual 3 x 30 mg/d (1.5 mg/kg BW/d) |
| **Treatment progress:** treatment started in May 2022 and is ongoing | |
| After 8 months | He experienced improvement after 2 days at the lower dosage (3 x 10 mg/day) in terms of muscle strength (he was able to move and raise the legs from the bed and also the head control improved) and of dysphagia. This initial improvement was maintained until now, but no further improvements were noticed after he reached the high dose (3 x 30 mg/day) from June 2022. He tolerated well the drug (no hypotension, only occasional priapism). |
| Last assessment after 2.5 years | After few days with dosages of 3 x 30 mg, he started moving the legs again. These improvements are maintained until now, and he keeps making small progresses. The parents found a significant improvement in speaking, eating, and in the movement of the arms. He can now even almost stay in the sitting position by himself, which was not possible before the therapy. Parents noted that when he had to discontinue the sildenafil medication due to unrelated surgery, he immediately became more flaccid and could not hold his head anymore. This improved again as soon as sildenafil therapy could be re-initiated after the surgery. |
| **Patient 5** | |
| Age range, sex | 0-5 years, male |
| Mutation | *MT-ATP6* m.8570T>C (98.5 % heteroplasmy) |
| Past medical history | The patient was diagnosed as floppy infant for severe muscle hypotonia. He had to be fed with a gastroenteral tube. He had to been admitted to hospital several times due to metabolic crises associated with febrile infections. |
| Symptoms | Due to the severe state of health, palliative care had been initiated shortly before the start of the sildenafil therapy. |
| Sildenafil Doses | 3 x 4 mg (1.5 mg/kg BW/d in 3 single doses) => has since been adapted to the increasing body weight. |
| **Treatment progress:** treatment started in September 2019 and is ongoing | |
| After 12 months | No more palliative care required. No metabolic crisis observed since starting the sildenafil treatment. The child has progressed in his motor development: can turn, can swallow soft foods, can sit stably at table, head control exists, has gained weight well. The medication is well tolerated. |
| Last assessment after 5 years | No further metabolic crises despite several febrile infections. In terms of his development, he has made significant progress. He understands and responds to speech, laughs, reaches for objects that interest him. No seizures. Normal night sleep. No unwanted side effects of sildenafil have been observed. |
| **Patient 6** | |
| Age range, sex | 6-10 years, female |
| Mutation | *MT-ATP6* m.8993T>C (100 % homoplasmy) |
| Past medical history | Normal psychomotor development, reaching all developmental milestones until the 6-10 years of age. |
| Symptoms | At 6-10 years of age, during febrile infection with vomiting she experienced a metabolic crisis with muscular hypotonia, loss of head control, declining cognitive capabilities, loss of speech, and dysphagia. Subsequently, she developed progressive distal spastic paraplegia and dystonia of upper libs and face. |
| Sildenafil Doses | 1.8 mg/kg BW/d in 3 single doses |
| **Treatment progress:** treatment started in June 2024 and is ongoing. | |
| After 3 months | The patient showed improvements in both motor and cognitive abilities. She regained full head control, was able to sit unsupported, and could stand with assistance. Her facial expressions improved, and her consciousness normalized. While she remained non-verbal, her vocalizations and emotional expression showed improvement. |
| After 8 months | Verbal communication capabilities improved, she gained muscle strength, the ataxia subsided, however, she suffers from progressive dystonia. Intentional motor movements are possible. |
