## Supplementary material for "Pluripotent stem cell-based drug discovery uncovers sildenafil as a treatment for mitochondrial disease": Table S9

**Table S9.** Questionnaire on side effects of continuous sildenafil use in LS patients treated with sildenafil for longer than 6 months.

Queries refer to whether **(i)** the symptom is absent (**N**), **(ii)** the severity of the symptom (**mild 1 to severe 5**), **(iii)** no information provided (**-**).

|  | **patient 1** | **patient 2** | **patient 3** | **patient 4** | **patient 5** | **patient 6** |
| --- | --- | --- | --- | --- | --- | --- |
| **Cardiovascular system** |  |  |  |  |  |  |
| arterial hypotension | N | N | N | N | N | N |
| postural syncope (dizziness when getting up) | N | N | N | - | - | N |
| worsening of existing heart failure | N | N | N | N | N | N |
| palpitations | **3^a^** | N | N | N | N | N |
| **Chest** |  |  |  |  |  |  |
| chest pain | N | N | N | - | - | N |
| difficulties breathing | N | N | **1** | N | N | N |
| burning feeling in the chest | N | N | N | - | - | N |
| tightness of the chest | N | N | - | - | - | N |
| **Gastrointestinal system** |  |  |  |  |  |  |
| difficulty in swallowing | N | N | N | N | N | **4^d^** |
| excessive hunger | **5^b^** | N | N | N | - | N |
| nausea | N | N | N | N | - | N |
| vomiting | N | N | N | N | - | N |
| stomach upset | N | N | N | N | - | N |
| burning feeling in the stomach | **3** | N | N | - | - | N |
| diarrhea | N | N | **1** | N | N | N |
| indigestion | N | N | **4** | N | - | N |
| **Genitourinary system** |  |  |  |  |  |  |
| priapism (prolonged, painful erection of penis) | N | N | - | N | N | - |
| bladder pain (increased urinary frequency) | N | N | N | N | - | N |
| pain on urination | N | N | N | N | - | N |
| urinary urgency | N | N | **3** | N | N | N |
| **Brain, head, and sensory system** |  |  |  |  |  |  |
| headaches, migraine | N | N | N | N | - | N |
| convulsions (seizures) | N | N | - | N | N | N |
| slurred speech | N | N | N | N | - | **?^e^** |
| trembling and shaking | N | N | **1** | N | N | N |
| lack of coordination | N | N | N | N | - | **?^f^** |
| nosebleeds | N | N | N | N | N | N |
| facial flushing | N | N | **5^c^** | N | - | N |
| transient visual disturbances | N | N | N | N | - | N |
| double vision | N | N | N | N | - | N |
| eye pain | N | N | N | N | - | N |
| deafness or hearing loss | N | N | N | N | N | N |
| tinnitus | N | N | N | N | - | N |
| **Psychiatric symptoms** |  |  |  |  |  |  |
| anxiety | N | N | N | N | N | N |
| confusion | N | N | N | N | N | N |
| unusual tiredness or weakness | N | N | N | N | N | N |
| difficulty in concentrating | N | N | N | N | N | N |
| abnormal dreams | N | N | N | - | - | N |
| sleepiness | N | N | N | N | N | N |
| sleeplessness | N | N | N | N | - | N |
| mental depression | N | N | N | N | - | N |

**Notes: (a)** The palpitations were investigated by a cardiologist, but no arrhythmia was detected. **(b)** The hyperphagia was related to a transition period from school to vocational training (3 years after the start of sildenafil treatment), had since improved. **(c)** The patient suffered from an itchy drug rash and had to discontinue the drug, whereupon the symptoms recurred. **(d)** The dysphagia is due to the Leigh syndrome (was present before sildenafil therapy). Cannot be evaluated due to **(e)** dysarthria and **(f)** incoordination due to Leigh syndrome (was present before sildenafil therapy).
