## Supplementary material for "Pluripotent stem cell-based drug discovery uncovers sildenafil as a treatment for mitochondrial disease": Resource Table

### **RESOURCES TABLE**

| **REAGENT or RESOURCE** | **SOURCE** | **IDENTIFIER** |
| --- | --- | --- |
| **Antibodies** | | |
| Purified anti-Pax-6 antibody, Poly19013 | BioLegend | 901301; RRID: AB_2565003 |
| Anti-β-Tubulin III Antibody, mouse monoclonal, 2G10 | Sigma-Aldrich | T8578 |
| PRKG1 polyclonal antibody | Proteintech | 21646-1-AP; RRID: AB_2878897 |
| Anti-Phosphorylated Vimentin (Ser55) mAb | MBL Life Science | D076-3 |
| Anti-Nestin monoclonal antibody, clone 10C2 (mouse) | Millipore | MAB5326; RRID: AB_2251134 |
| Occludin Monoclonal Antibody (OC-3F10) | Invitrogen | 33-1500; RRID: AB_2533101 |
| ZO-1 Polyclonal antibody | Proteintech | 21773-1-AP; RRID: AB_10733242 |
| Cleaved Caspase-3 (Asp175) Antibody | Cell Signaling Technology | 9661 |
| Anti-Iba1 antibody [EPR16589] - Mouse IgG1 (Chimeric) | Abcam | Ab283319; RRID: AB_2924797 |
| ATP6 polyclonal antibody | Immunological Science | AB-83828 |
| Anti-GAPDH antibody [6C5] | Abcam | Ab8245; RRID: AB_2107448 |
| Goat anti-Mouse IgG (H+L) Highly Cross-Adsorbed Secondary Antibody, Alexa Fluor 568 | Invitrogen | A-11031; RRID: AB_144696 |
| Donkey anti-Rabbit IgG (H+L) ReadyProbes™ Secondary Antibody, Alexa Fluor™ 488 | Invitrogen | R37118; RRID: AB_2556546 |
| Donkey anti-Mouse IgG (H+L) Highly Cross-Adsorbed Secondary Antibody, Alexa Fluor™ 488 | Invitrogen | A-21202; RRID: AB_141607 |
| Donkey anti-Rabbit IgG (H+L) Highly Cross-Adsorbed Secondary Antibody, Alexa Fluor™ Plus 647 | Invitrogen | A32795; RRID: AB_2762835 |
| Donkey anti-Rabbit IgG (H+L) Highly Cross-Adsorbed Secondary Antibody, Alexa Fluor™ 488 | Invitrogen | A-21206; RRID: AB_2535792 |
| Goat anti-Mouse IgG (H+L) Cross-Adsorbed Secondary Antibody, Alexa Fluor™ 633 | Invitrogen | A-21050 |
| Goat Anti-Mouse IgG (H + L)-HRP Conjugate | BioRad | 1706516; RRID: AB_2921252 |
| Anti-Rabbit IgG (H+L), HRP Conjugate | Promega | W4018 |
| Anti-Mitofilin antibody [EPR8749] | abcam | Ab137057; RRID: AB_3676556 |
| **Chemicals, peptides, and recombinant proteins** | | |
| ROCK inhibitor Y-27632 | Enzy Life Sciences | ALX-270-333-M005 |
| Non-essential amino acids (MEM-NEAA) 100X | Gibco | 11140-050 |
| Sodium Pyruvate | Gibco | 11360070 |
| B-27 with Vitamin A (50×) | Gibco | 17504044 |
| B-27 without Vitamin A (50×) | Gibco | 12587010 |
| N2 Supplement (100×) | Gibco | 17502-048 |
| GDNF | R & D System | 212-GD-010 |
| StemPro Accutase | Thermo Fisher Scientific | A1110501 |
| Neurobasal | Gibco | 21103-049 |
| DMEM/F/12 | Gibco | 31330038 |
| DMEM glucose-free | Gibco | 11966025 |
| Neurobasal A-Medium glucose-free | Gibco | A2477501 |
| Db-cAMP | StemCell Technologies | 73886 |
| GlutaMAX | Gibco | 35050061 |
| FBS | Gibco | 10270106 |
| Heparin | Merck | 375095 |
| MycoZap | Lonza | VZA-2012 |
| Pen/Strep | Gibco | 15140122 |
| 2-mercaptoethanol | Gibco | 31350010 |
| L-glutamine | Gibco | 25030081 |
| Recombinant Human NT-3 | Peprotech / Biozol | 450-03 |
| BDNF | MACS Miltenyi | 130-096-811 |
| CHIR99021 | Sigma Aldrich | SML1046 |
| Laminin | Sigma Aldrich | L2020 |
| FGF8-a | R & D systems | 4745-F8-050 |
| TGFbeta 3 | StemCell Technologies | 78156 |
| cis-4,7,10,13,16,19-Docosahexaensäure (DHA) | Sigma-Aldrich | D2534 |
| Human Recombinant Activin A | StemCell Technologies | 78001.1 |
| Doxycycline hydrochloride (DOX) | Sigma Aldrich | D3072 |
| WNT antagonist IWR1 | EMD Millipore Corp | 681669 |
| 16% Paraformaldehyde (PFA) | Thermo Fisher Scientific | 28906 |
| Hoechst 33342 | Invitrogen | H3570 |
| Matrigel, Growth Factor reduced | Corning | 356231 |
| (+)-sodium L-ascorbate (Vitamin C) | Sigma Aldrich | A4034 |
| TGF-b3 | StemCell Technologies | 78156 |
| Purmorphamine | Sigma Aldrich | 540220 |
| Dorsomorphine | Sigma-Aldrich | P5499 |
| StemMACS™ SB431542 | MACS Miltenyi | 130-105-336 |
| Donkey serum | Merck Millipore | S30 |
| Triton-X-100 | Sigma-Aldrich | 93443 |
| Tween 20 | Sigma-Aldrich | P1504 |
| abberior STAR 635 (STAR RED) | Abberior | ST635-1002 |
| DAPI | Sigma-Aldrich | D9542 |
| MitoProbe™ TMRM Assay Kit for Flow Cytometry | Invitrogen | M20036 |
| Incucyte dye red | Sartorius | 4717 |
| Sildenafil-d_3_ | CDN isotopes | D-6366 |
| HpaII | NEB | R0171 |
| StuI | NEB | R0187 |
| XbaI | NEB | R0145 |
| Geltrex™ Reduced-Growth Factor Basement-Membrane Matrix, LDEV-free, stem-cell qualified | Gibco | A1413302 |
| FCCP | Biozol | SEL-S8276 |
| Antimycin A | Sigma-Aldrich | A8674 |
| Sildenafil citrate | Sigma | SML3033 |
| Sildenafil | Selleckchem | S468402 and S468403 |
| Mowiol with 0.1 % 1,4-Diazabicyclo[2.2.2]octan (DABCO) | Carl Roth | 0713.1 |
| SuperFrost Plus glass slides | VWR | 631-0447 |
| Pro-Long Glass Antifade Mountant | Invitrogen | P36984 |
| oligo d(T)_18_ primers | Thermo Fisher Scientific | SO132 |
| dNTP mix | Thermo Fisher Scientific | 10319879 |
| mTeSR™ Plus | STEMCELL Technologies | 100-0276 |
| KnockOut^TM^ serum replacement | Gibco | 10828-028 |
| Human Endothelial-SFM | Gibco | 11111044 |
| Hank’s Balanced Salt Solution (HBSS) | Sigma | H9394 |
| Oregon Green 488 BAPTA-1 AM | Invitrogen | O6807 |
| Pluronic |  |  |
| Sodium azide | Honeywell | 16466349 |
| Deoxy-D-glucose | Apollo Scientific | OR3900T |
| **Critical commercial assays** | | |
| Lactate-Assay kit | Sigma Aldrich | MAK064 |
| CellTiter-Glo® Luminescent Cell Viability assay | Promega | G7571 |
| Rnase-Free Dnase Set (50) (250) | Qiagen | 79254 |
| RNeasy Mini Kit (50) | Qiagen | 74104 |
| First Strand cDNA Synthesis Kit \| Thermo Scientific \| K1612 | Thermo Scientific | K1612 |
| PDS - Papain Dissociation System \| Cell Systems \| LK003150 | Cell Systems | LK003150 |
| Nucleo-Spin Tissue kit | Macherey-Nagel | 740952.50 |
| SYBR^TM^ Green PCR Master Mix | Applied Biosystems | 4364344 |
| Bicinchoninic Acid (BCA) protein assay kit | Thermo Scientific | 23225 |
| NucleoSpin RNA Plus kit | Macherey-Nagel | 740984.50 |
| RNA Cleanup XP beads | Agencourt |  |
| TURBO DNase rigorous treatment | Invitrogen | AM1907 |
| TruSeq Stranded Total LT Sample Prep Kit | Illumina | N/A |
| Chromium Single Cell 3' (vNext) Reagent Kit | 10X Genomics | N/A |
| ViaLight^TM^ Plus Kit | Lonza | LT07-221 |
| CyQUANT Cell Proliferation Assay | Invitrogen | C7026 |
| jetPRIME transfection | Polyplus | 101000027 |
| **Experimental models: Cell lines** | | |
| Human iPSC: BIHi043-A | [Helmholtz Zentrum München (HMGU)](https://hpscreg.eu/browse/provider/690) | PMID 29396371 |
| Human iPSC: HHUUKDi009-A | Heinrich-Heine-Universität Düsseldorf (HHUUKD) | PMID 28132834 |
| Human iPSC: CRMi003-A | RUCRD Infinite Biologics | PMID 36459969 |
| Human iPSC: BIHi269-B | Berlin Institute of Health (BIH) | PMID 36669241 |
| Human iPSC: HVRDi004-B | Synthego | PMID 36459969 |
| Human iPSC: IUFi004-A | Cell Applications | PMID 38217996 |
| Human iPSC: BIHi266-A | Berlin Institute of Health (BIH) | PMID 36669241 |
| Human iPSC: WISCi004-B | WiCell | PMID 18029452 |
| Human iPSC: HHUi001-A | Universitätsklinikum Düsseldorf (HHU) | PMID 28132834 |
| Human iPSC: HHUi002-A | Universitätsklinikum Düsseldorf (HHU) | PMID 28132834 |
| Human iPSC: HHUi003-C | Universitätsklinikum Düsseldorf (HHU) | PMID 36137325 |
| Human iPSC: MDCi008-A | Max Delbrück Center Berlin Buch (MDC) | PMID 35279592 |
| Human iPSC: MDCi009-A | Max Delbrück Center Berlin Buch (MDC) | PMID 35279592 |
| Human iPSC: MDCi010-A | Max Delbrück Center Berlin Buch (MDC) | PMID 35279592 |
| Human iPSC: BIHi267-B | Berlin Institute of Health (BIH) | PMID 36669241 |
| Human iPSC: C1_mut2 | Max Delbrück Center Berlin Buch (MDC) | PMID 33771987 |
| Human iPSC: E9 | Max Delbrück Center Berlin Buch (MDC) | PMID 33771987 |
| **Experimental models: Organisms/strains** | | |
| Germline *Ndufs4* KO, C57/BL6/J background | Supplementary reference Kruse et al. ^5^ |  |
| **Oligonucleotides** | | |
| ATP6: Forward: CAACCGACTAATCACCACCC, Reverse: GTTGAGCCGTAGATGCCGTC | IDT | N/A |
| ATP6_2: Forward: AACCAATAGCCCTGGCCGTA, Reverse: AGGGCTCATGGTAGGGGTAAA | IDT | N/A |
| GAPDH: Forward: CTGGTAAAGTGGATATTGTTGCCAT, Reverse: TGGAATCATATTGGAACATGTAAACC | IDT | N/A |
| OAZ1: Forward: GGATCCTCAATAGCCACTGC, Reverse: TACAGCAGTGGAGGGAGACC | IDT | N/A |
| NESTIN: Forward: TTCCCTCAGCTTTCAGGAC, Reverse: GAGCAAAGATCCAAGACGC | IDT | N/A |
| PAX6: Forward: CCAGGGCAATCGGTGGTAGT Reverse: ACGGGCACTCCCGCTTATAC | IDT | N/A |
| CTIP: Forward: TGGGTGCCTGCTATGACAAG, Reverse: GATGCCTTTCGTGGGTGAGA | IDT | N/A |
| SOX2: Forward: GTATCAGGAGTTGTCAAGGCAGAG, Reverse: TCCTAGTCTTAAAGAGGCAGCAAAC | IDT | N/A |
| PRKG1: Forward: ACAACTGTACCCGGACAGCGA, Reverse:  TCCTCTTGCACCCTGCCTGAT | IDT | N/A |
| OCT4: Forward: GTGGAGGAAGCTGACAACAA, Reverse: ATTCTCCAGGTTGCCTCTCA | IDT | N/A |
| NANOG: Forward: CCTGTGATTTGTGGGCCTG, Reverse: GACAGTCTCCGTGTGAGGCAT | IDT | N/A |
| Myco-f1: Forward: CGCCTGAGTAGTACGTTCGC | IDT | N/A |
| Myco-f2: Forward: GCGGTGTGTACAAACCCCGA | IDT | N/A |
| Myco-f3: Forward: TGCCTGAGTAGTCACTTCGC | IDT | N/A |
| Myco-f4: Forward: CGCCTGGGTAGTACATTCGC | IDT | N/A |
| Myco-f5: Forward: CGCCTGAGTAGTAGTCTCGC | IDT | N/A |
| Myco-f6: Forward: TGCCTGGGTAGTACATTCGC | IDT | N/A |
| Myco-r1: Reverse: GCGGTGTGTACAAGACCCGA | IDT | N/A |
| PRKG1 siRNA: Forward: GGAUAGAGGUUCGUUUGAATT, Reverse:  UUCAAACGAACCUCUAUCCCT | Ambion | 4390843 |
| **Software and algorithms** | | |
| CellProfiler 4.2.5. | Carpenter et al., 2006 | https://cellprofiler.org/ |
| GraphPad Prism 5.01 | GraphPad Software | N/A |
| Illustrator | Adobe |  |
| Cell Ranger (v.7.10) | 10x Genomics |  |
| Ultivo triple-quadrupole mass spectrometer | Agilent Technologies |  |
| MassHunter Software | Agilent Technologies |  |
| MToolBox v.1 |  | https://github.com/mitoNGS/MToolBox |
| ImageJ | Schindelin et al., 2012 | [Fiji](https://imagej.net/software/fiji/) |
| IGV viewer v2.163 | IGV | [https://igv.org](https://igv.org/) |
| Genetic Analyzer | Applied Biosystems | RRID:SCR_021901 |
| CELLCYTE Studio | Cytena | N/A |
| ZEN Microscopy Software | Zeiss | N/A |
| NanoDrop 2000 | Thermo Fisher Scientific | N/A |
| Nanodrop Spectrophotometer ND1000 | peQlab | N/A |
| CFX96 software | Bio-Rad | N/A |
| Image Lab software, 6.1 | Bio-Rad Laboratories | N/A |
| GeneMapper ID v.3.2.1 | Applied Biosystems | N/A |
| ND-1000 software (V3.8.1) | Thermo Fisher Scientific | N/A |
| Xcalibur software | Thermo Fischer Scientific | N/A |
| TraceFinder 5.1 software | Thermo Fischer Scientific | N/A |
| Columbus software (v 2.9.0) | Revvity | N/A |
| FlowJo software (version 7.6) | FlowJo | N/A |
| OriginPro Software | OriginLab Corporation | N/A |
| MassHunter Software | Agilent Technologies |  |
| **Other** | | |
| CytoSmart Cell Counter | Greiner Bio-One | 6749 |
| Operetta® CLS™ | Revvity | HH16000020 |
| 96-well Cell Culture Microplate, PS, F-Bottom, black TC, µCLEAR, 96-well black, clear bottom | Greiner Bio-One | 655090 |
| BIOFLOAT 96 well plate 4PCS | FaCellitate GmbH | F202003 |
| µ-Plate 96 Well Square | Ibidi | 89626 |
| ZEISS Axio Observer Apotome 3 | Zeiss | [ZEISS Apotome 3: Optical sectioning in widefield fluorescence microscopy](https://www.zeiss.com/microscopy/en/products/light-microscopes/widefield-microscopes/apotome-3.html?utm_source=google&utm_medium=search-ad&utm_campaign=C-00011305&utm_content=global&gad_source=1&gclid=CjwKCAjwwe2_BhBEEiwAM1I7sQHogKcYWh3auOR02OLSze5whUYNBdNDtw-6QEokXWe8TENhMAtNWxoCJoEQAvD_BwE) |
| Vibratome Microm HM 650 V | Thermo Fisher Scientific | 920120 |
| Ultivo triple-quadrupole mass spectrometer | Agilent Technologies | G6465BA |
| EnSight multimode plate reader | Revvity | N/A |
| Cellcyte X | Cytena | [CELLCYTE X™ - Live Cell Imager And Analyzer \| CYTENA](https://www.cytena.com/products/live-cell-imaging/cellcyte-x/) |
| INFINITY platform | Abberior Instruments | [INFINITY - @abberior.rocks](https://abberior.rocks/superresolution-confocal-systems/infinity/) |
| Vevo 2100 VisualSonics System | FUJIFILM VisualSonics | N/A |
| Promethion | Sable Systems International | [Promethion Core Metabolic and Behavioral Phenotyping Systems](https://www.sablesys.com/products/promethion-core-line/) |
| Infinite M1000 Pro | TECAN | [Tecan \| Thermo Fisher Scientific - DE](https://www.thermofisher.com/de/en/home/industrial/pharma-biopharma/drug-discovery-development/dd-misc/instrument-compatibility-portal/tecan.html) |
| Confocal laser scanning microscope C1 | Nikon Mikroskope Solutions | [Nikon's Digital Eclipse C1 Microscope System Delivers High Resolution Confocal Images at Sensible Price \| News \| Nikon Instruments Inc.](https://www.microscope.healthcare.nikon.com/about/news/nikons-digital-eclipse-c1-microscope-system-delivers-high-resolution-confocal-images-at-sensible-price) |
| Mastercycler X50s | Eppendorf | 6311000010 |
| CFX96™ Real-Time System qPCR machine | Bio-Rad | [CFX96 Touch Real-Time PCR Detection System \| Bio-Rad](https://www.bio-rad.com/de-de/product/cfx96-touch-real-time-pcr-detection-system?ID=LJB1YU15&s_kwcid=AL%2118120%213%21695727856508%21%21%21g%21%21%2115996395757%21137889175132&WT_mc_id=241223044753&WT_srch=1&WT_knsh_id=_kenshoo_clickid_&gad_source=1&gclid=CjwKCAjwwe2_BhBEEiwAM1I7sZZZPMCy4fFLJPdwCbv3G8eDks587nHjl4syI5kWt055BVBhxFgQDBoC4lsQAvD_BwE) |
| ChemiDoc MP Imaging system | Bio-Rad | [ChemiDoc MP Imaging System \| Bio-Rad](https://www.bio-rad.com/de-de/product/chemidoc-mp-imaging-system?ID=NINJ8ZE8Z) |
| low-attachment U-bottom 96-well plates | Corning | 3474 |
| AggreWell | STEMCELL Technologies | 34815 |
| NovaSeq 6000 system | Illumina | [NovaSeq 6000 System \| Powerful sequencing with scalable throughput](https://www.illumina.com/systems/sequencing-platforms/novaseq.html) |
| Dionex Ultimate 3000 | Thermo Scientific | [UltiMate 3000 HPLC and UHPLC Systems \| Thermo Fisher Scientific - DE](https://www.thermofisher.com/de/en/home/industrial/chromatography/liquid-chromatography-lc/hplc-uhplc-systems/ultimate-3000-hplc-uhplc-systems.html) |
| timsTOF SCP mass spectrometer | Bruker Daltonics | [timsTOF SCP \| Bruker](https://www.bruker.com/en/products-and-solutions/mass-spectrometry/timstof/timstof-scp.html) |
| SeQuant ZIC-pHILIC | Merck | 1.50462 |
| LSR-Fortessa X-20 | Becton Dickinson | [LSRFortessa™ X-20 \| Benchtop Flow Cytometer](https://www.bdbiosciences.com/en-us/products/instruments/flow-cytometers/research-cell-analyzers/bd-lsrfortessa-x-20) |
| 384-well black-wall, clear-bottom plates | Revvity | 6007460 |
| Janus MDT | Revvity | YJLM001 |
| Victor Multiple Plate Reader Spectrophotometer | Revvity | HH35000500 |
| Vibratome Microm HM 650 V | Thermo Fischer Scientific | 920120 |
| Millicell-CM inserts | Millipore | [Millicell® Cell Culture Inserts - Zellkulturplatten-Einsätze](https://www.merckmillipore.com/DE/de/product/Millicell-Cell-Culture-Inserts,MM_NF-C10504" \o "https://www.merckmillipore.com/DE/de/product/Millicell-Cell-Culture-Inserts,MM_NF-C10504) |
| Eclipse FN-1 | Nikon | [ECLIPSE FN1 \| Upright Microscopes \| Microscope Products \| Nikon Instruments Inc.](https://www.microscope.healthcare.nikon.com/products/upright-microscopes/eclipse-fn1) |
